# Mechanistic model for booster doses effectiveness in healthy, cancer and immunosuppressed patients infected with SARS-CoV-2

**DOI:** 10.1101/2022.06.30.22277076

**Authors:** Chrysovalantis Voutouri, C. Corey Hardin, Vivek Naranbhai, Mohammad R. Nikmaneshi, Melin J. Khandekar, Justin F Gainor, Triantafyllos Stylianopoulos, Lance L. Munn, Rakesh K. Jain

## Abstract

SARS-CoV-2 vaccines are effective at limiting disease severity, but effectiveness is lower among patients with cancer or immunosuppression. Effectiveness wanes with time and varies by vaccine type. Moreover, vaccines are based on the ancestral SARS-CoV-2 spike-protein that emerging variants may evade. Here, we describe a mechanistic mathematical model for vaccination-induced immunity, validate it with available clinical data, and predict vaccine effectiveness for varied vaccine platforms in the setting of variants with ability to escape immunity, increased virulence, or enhanced transmissibility. We further account for concurrent cancer or underlying immunosuppression. The model confirms enhanced immunogenicity following booster vaccination in immunosuppressed patients but predicts at least one more booster dose is required for these individuals to maintain protection. We further studied the impact of variants on immunosuppressed individuals as a function of the interval between multiple booster doses. Our model is useful for planning future vaccinations, and tailoring strategies to risk groups.

**Significance Statement:** Current SARS-CoV-2 vaccines are effective at preventing COVID-19 or limiting disease severity in healthy individuals, but effectiveness is lower among patients with cancer or immunosuppression. Here, we address the need for predictions of vaccine effectiveness over time by building on our mathematical framework to account for vaccination-induced immunity. A booster dose of both mRNA vaccines can induce a robust enhancement of both antibody levels and numbers of pertinent types of adaptive immune cells, which is predicted to provide sufficient protection for more than one year in healthy patients. However, our model suggests that for immunosuppressed people or patients with cancer receiving an immunosuppressive treatment, the booster effect may wane, and perhaps could be considered on a more frequent basis.

## Introduction

COVID-19 has created unprecedented challenges for healthcare systems and society. Despite the effectiveness and widespread availability of vaccines in many countries, the pandemic persists. The large unvaccinated population, incomplete vaccine response in immunosuppressed patients, and waning immunity following vaccination all contribute to the persistence of the crisis. The large unvaccinated and immunosuppressed populations, moreover, can allow for continued viral spread and emergence of new SARS-CoV-2 variants (1, 2). Understanding the limits of vaccine-induced immunity is vital to ongoing efforts to address the COVID-19 pandemic. Indeed, numerous clinical studies are taking place worldwide to evaluate the protective capacity of mRNA and vector vaccines over time against the ancestral and emerging variants of the SARS-CoV-2 virus, for those vaccinated with or without prior infection as well as for patients who may have a suboptimal vaccine response due to underlying disease (e.g., cancer) or due to treatment with immunosuppressive therapies (e.g., chemotherapy) (3-14). Based on these studies, it is now accepted that booster doses are required to maintain sufficient levels of immunity, particularly in patients with cancer who receive chemotherapy or other immunosuppressive treatment or for other individuals receiving immunomodulatory medications (4, 7, 9, 13-18). These findings have been rapidly incorporated into public health recommendations, but it remains uncertain how long booster doses ensure immunity, how effective they are against emerging variants or how to tailor vaccine regimens in high-risk populations. There is currently no proven framework for prospective prediction of optimal vaccine schedules in a dynamically evolving pandemic, especially for the high-risk patient populations most likely to suffer from deleterious consequences of waning immunity.

Computational modeling can be used to explore the biological mechanisms of COVID-19 to identify better treatment strategies (19-26). We recently developed a mechanistic model of COVID-19 that includes lung infection by the SARS-CoV-2 virus, the innate and adaptive immune responses, local and systemic thrombosis and a pharmacokinetic/pharmacodynamic (PK/PD) model of the dissemination of virus, cytokines and micro-thrombi throughout the body (27). This mechanistic model(28) consists of a series of differential equations which describe the dynamics of the above processes and enable the testing of hypotheses about the interactions between viral infection and various components of the immune response to that infection. As such, it should be contrasted with data-driven models(29, 30) which seek to develop predictors of individual patients’ courses though the analysis of large clinical data sets. Indeed, the large number of parameters in our model would be difficult to precisely validate against clinical data. Here we seek to develop hypotheses about possible consequences for the immune response to viral infection with impairments in specific arms of the immune system. Such hypotheses should, of course, be subject to verification by experimental data. Given the uncertainty in parameter values, we conduct extensive sensitivity analyses in order to determine how robust our hypotheses are to changes in key parameters. Here, we address the need for predictions of vaccine effectiveness over time by building on our mathematical framework to account for vaccination-induced immunity. We include the separate mechanisms of mRNA and viral-vector vaccines in healthy, immunosuppressed, and patients with cancer. The model accounts for translation of viral antigens, the production of antigen and its presentation by dendritic cells, the subsequent activation of T cells and B cells to create CD4^+^ and CD8^+^ effector and memory T cells as well as short-lived and long-lived plasma (antibody-secreting) B cells. We also consider various anti-cancer therapies, including chemotherapy and PD-L1/PD-1 immune checkpoint blockade (Fig 1).

**Figure 1.**
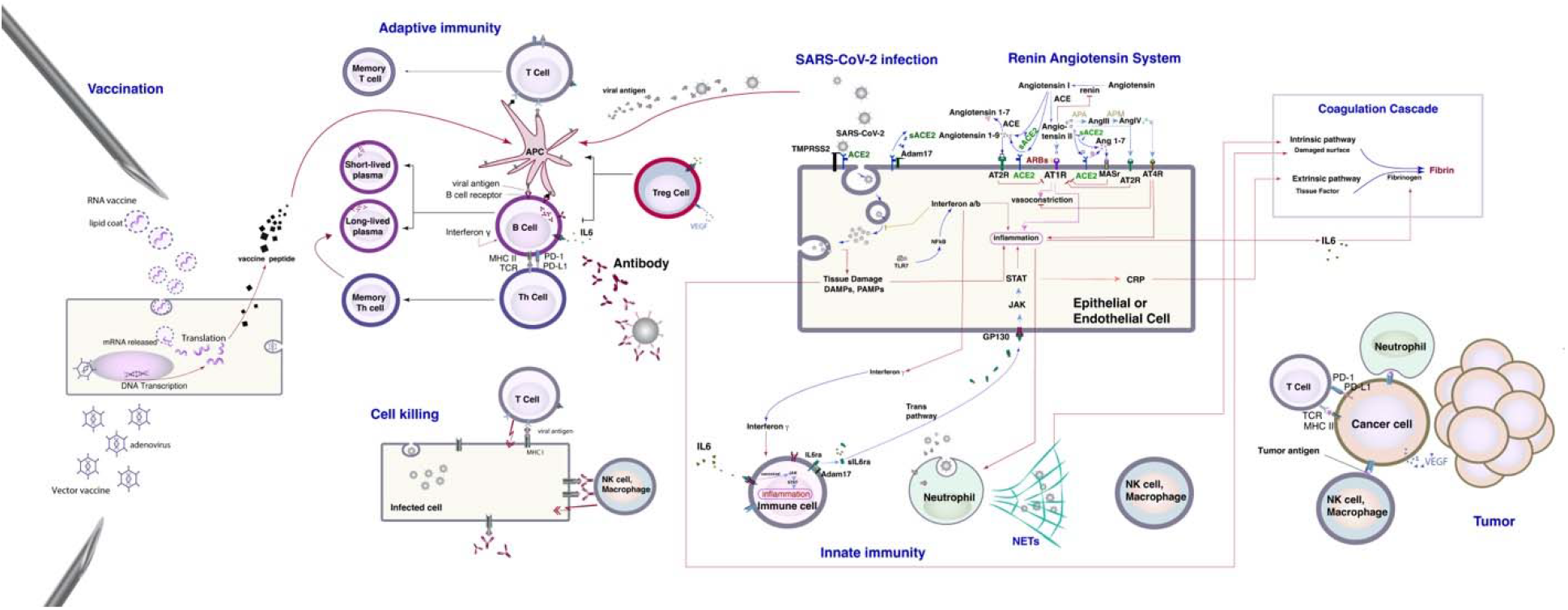
Schematic representation of the mathematical model. The basic components of the model are: i) a detailed model of lung infection by SARS-CoV-2 that includes innate and adaptive immune responses, known mechanisms of the renin-angiotensin system (RAS) and the coagulation cascade. Intracellular virus initiates inflammatory pathways through toll-like receptors and NFκB, which produces interferons and other inflammatory cytokines. The viral antigens, along with inflammatory cytokines, facilitate activation of naïve B and T cells, creating virus-specific effector cells. Activation of naïve immune cells is controlled by viral antigen strength and the status of immune checkpoint inhibition (specifically PD-L1/PD-1). ii) A Pharmacokinetic/Pharmacodynamic model of dissemination of viral particles, cytokines, micro-thrombi and antibodies in the major organs (lung, heart, liver, brain, spleen, gastro-intestinal, upper body, lower body, torso, cardiac vessels and the tumor; see Fig. S1). iii) All steps of vaccination-induced immunity for mRNA and vector vaccines, including the translation of viral antigens, the production of antigen presenting cells by dendritic cells, the subsequent activation of T cells and B cells to create CD4^+^ and CD8^+^ effector and memory T cells as well as short-lived and long-lived plasma (antibody-secreting) B cells. iv) Tumor cells and interactions with the immune system. Proliferation of tumor cells depends on oxygen levels in the tissue, and their death rate on the interaction of cancer cells with immune cells (effector CD8^+^ T cells, natural killer cells, type 1 macrophages and neutrophils) as well as on the effect of cancer therapy. We assume no tumor cell-virus interactions.

We first validate our model with clinical data for vaccinated healthy individuals and patients with cancer, and then we compare model predictions of temporal changes in antibody levels directed against the SARS-CoV-2 spike protein to clinical data to demonstrate the predictive capabilities of the model. Subsequently, we use the model to make predictions regarding the anticipated need for booster vaccine doses among healthy individuals and those with cancer and/or receiving immunosuppressive therapies. Finally, we use the model to explore how specific characteristics of potential viral variants would influence immune escape and infectivity. We find that for healthy individuals vaccinated and boosted with mRNA-1273 or BNT-162b2a, robust immunogenicity against the ancestral and delta variant extends beyond a year. Immunogenicity is also significantly enhanced following booster vaccination in patients with cancer on various anti-cancer therapies and for patients without cancer on immunosuppressive agents, such as B cell depleting therapy, long term corticosteroids or TNF blockers. However, our model predicts that one or more additional booster doses will be required for these individuals to maintain long term immunity. Similar results were seen with Ad26.COV2.S, although antibody levels were less than for the mRNA vaccines.

## Results

### Validation of model predictions for SARS-CoV-2 spike protein with available clinical data for healthy individuals and cancer patients

A description of the model is provided in the caption of Fig. 1. The values of the model parameters related to viral infection and the pharmacokinetics/ pharmacodynamics of COVID-19 were defined and validated in previous work (27) (and are presented in the Table S3). Here, we incorporated new equations and model parameters related to vaccination-induced immunity, summarized in Table S1. These parameters include the affinity for and uptake rate of vaccine particles by cells, the rate of DNA transcription to mRNA, the production rate of viral antigens and the degradation rates of the vaccine and the viral antigen. To calculate the baseline values of these parameters, we fitted model predictions to clinical data of average values of anti-spike antibody levels directed against the SARS-CoV-2 spike protein in healthy individuals who received either two doses of the mRNA vaccines (BNT-162b2a or mRNA-1273) or a single dose of the Ad26.COV2.S vaccine presented in our previous work (Naranbhai V et al. (12)). Note that these initial fits were not intended to yield parameters that precisely predict the response of any individual patient but rather baseline values for our further analysis. To quantify the quality of the fit, we provide in the figure caption the χ^2^ values, calculated for the model predictions versus the clinical data. These values are: χ^2^= 0.0081 for the BNT-162b2 vaccine, χ^2^=0.0484 for the mRNA-1273 vaccine and χ^2^=0.1139 for the Ad26.COV2.S vaccine. To account for inherent clinical heterogeneity (which may not be accounted for by any single set of parameter choices) and assess the robustness of our predictions to variations from the best-fit parameter values, we repeated simulations using a range of values within an order of magnitude around the baseline values. Simulations were repeated for all possible combinations among parameters taking 100 different values for each parameter. The results are presented in the figures as standard error bars from the baseline values.

Subsequently, we compared model predictions with additional data from seven independent clinical studies on booster doses (i.e., a third dose for BNT-162b2a and mRNA-1273 and a second dose for Ad26.COV2.S) for healthy individuals as well as with clinical data of patients living with cancer who had received chemotherapy or PD-L1/PD-1 immune checkpoint blockers and had been vaccinated (8-12, 31, 32). Modifications made to specific model parameters in order to fit various cancer treatments are shown in Table S1. The model was able to provide accurate predictions of antibody levels for all sets of clinical data considered (Fig 2, and the corresponding χ^2^ values in the figure caption).

**Figure 2.**
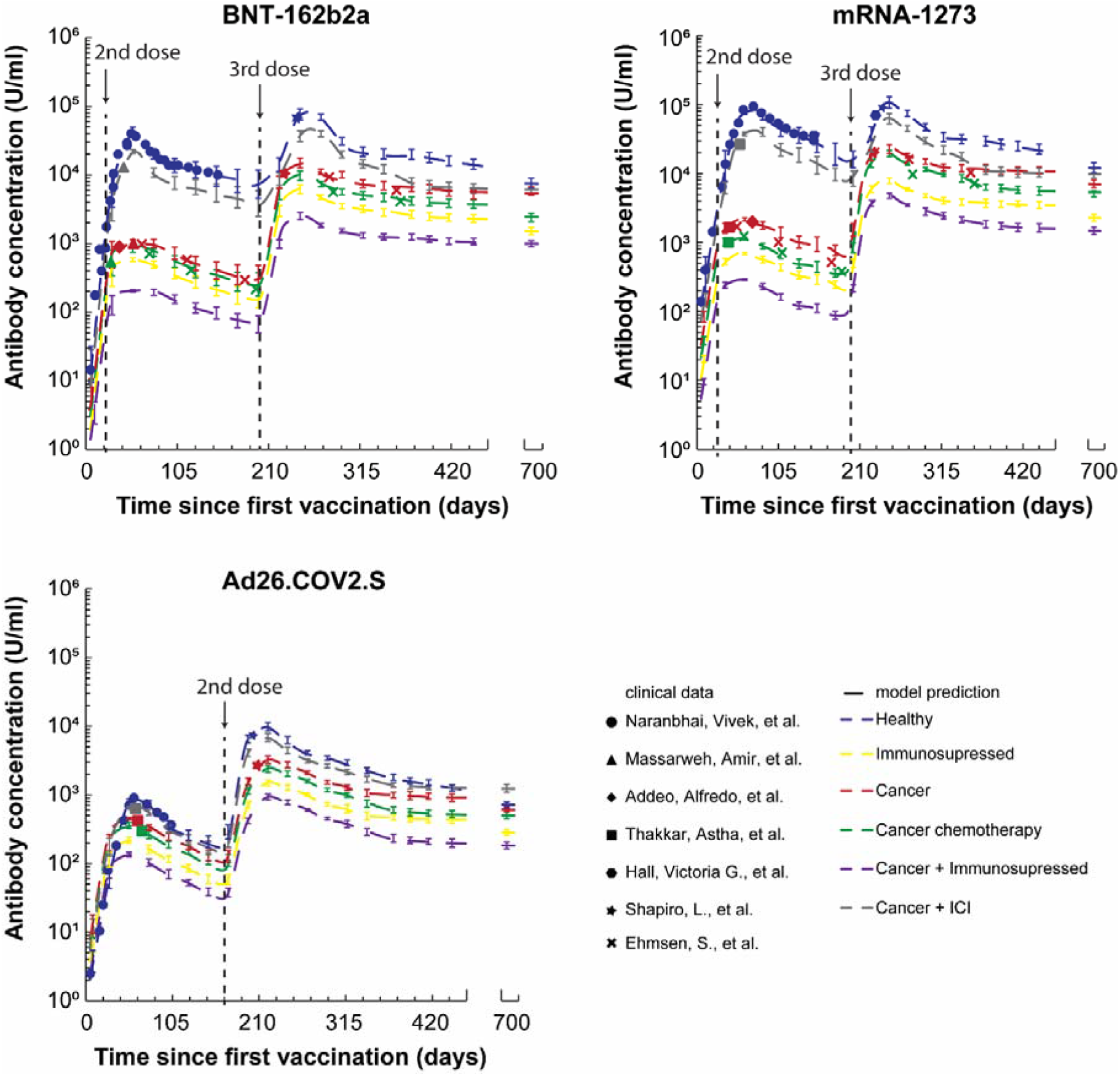
Model predictions and validation with clinical data of anti-spike antibody levels from vaccinated healthy people and lung cancer patients as a training set. The scatter symbols correspond to clinical data (8-12, 31, 32) and the dash lines to model predictions. The sum of the squared error (i.e., χ^2^ value) between the clinical data for each of the 6 studies and the corresponding model predictions for the baseline values of model parameters were calculated. For the temporal variation of antibody levels in healthy: χ^2^= 0.0081 for the BNT-162b2 vaccine, χ^2^=0.0484 for the mRNA-1273 vaccine and χ^2^=0.1139 for the Ad26.COV2.S vaccine. For the single time point data of patients with cancer: Cancer: χ^2^=0.00013 (BNT-162b2), χ2=0.0045(mRNA-1273), χ^2^=0.0083 (Ad26.COV2.S). Cancer+ICI: χ^2^=0.00032(BNT-162b2), χ^2^=0.00214(mRNA-1273), χ2=0.00074 (Ad26.COV2.S). Cancer + chemotherapy: χ^2^=0.00061 (BNT-162b2), χ^2^=0.0053 (mRNA-1273), χ^2^=0.0072 (Ad26.COV2.S). Error bars present the standard error of antibody concentration for all values of model parameters considered.

This ability of our modelling approach to accurately predict antibody dynamics is critical given that antibody levels are directly linked to immune protection from SARS-CoV-2 infection (33). Mathematical modelling of the relationship between antibody concentration and neutralizing titres (20), also justify the use of antibody concentrations as a correlate of protection.

### Robustness of spike-specific antibody levels in healthy individuals, patients with cancer and immunosuppressed individuals

We first assessed the long-term robustness of spike-specific antibody levels by simulating the effects of vaccination over a period of 100 weeks (i.e., 700 days), including a 3^rd^ dose for the mRNA vaccines and a 2^nd^ dose of the vector vaccine. Figure 2 presents the model predictions of anti-spike antibody levels as a function of time. In agreement with clinical studies, the model predicts a significant reduction in the antibody levels in all types of vaccines within 6 months and a rapid and robust increase in antibody levels following a booster dose. Interestingly, for healthy individuals, the antibody levels following a booster dose are predicted to stay above 10,000 U/ml for the mRNA vaccines and 1,000 U/ml for the vector vaccine for the entire simulated period (i.e.,78-80 weeks from booster dose) but reach lower values for patients with cancer and particularly for immunosuppressed patients with or without cancer. Antibody levels above 1000 U/ml(14) have been suggested to correlate with protective immunity. The only exception are patients with cancer that receive PD-L1/PD-1 inhibition; they exhibit the same antibody levels as healthy individuals per our model predictions. Even though such long-term data from a booster dose are still not available, model predictions agree with the limited data of antibody levels and T cell immune responses available as of now (34-38) (Fig 2) as well as with predictions made by extrapolating clinical data after a 3^rd^ mRNA vaccination for patients with cancer on chemotherapy, anti-CD20 therapy or on BTK inhibitors (32).

### Cell mediated immunity following vaccination

We next examined the effects of vaccination on the levels of immune cells that influence the severity of viral infection (Fig 3 for BNT-162b2a, Fig S1 and S2 for mRNA-1273 and AD26.COV2.S respectively). Within a period of 6 months from the second dose of the mRNA vaccines, B cell, CD4^+^, CD8^+^ T cell and antigen presenting cell (APC) levels are predicted to drop up to 50%, mirroring the corresponding decrease in antibody levels. For the vector vaccine, the decrease in the 6-month period is less profound, but the peak levels achieved are also significantly lower compared to the mRNA vaccines. Following a booster dose, there is an enhancement of all immune cell levels against the virus. Healthy individuals and patients with cancer receiving PD-L1/PD-1 inhibition therapy benefitted the most, and at the end of the 100-weeks simulation, the levels of all immune cells were up to twice as high as before the booster dose. In contrast, immunosuppressed individuals with or without cancer benefit the least, with a less than 50% increase in APCs and memory B cells, CD4^+^ and CD8^+^ T cells.

**Figure 3.**
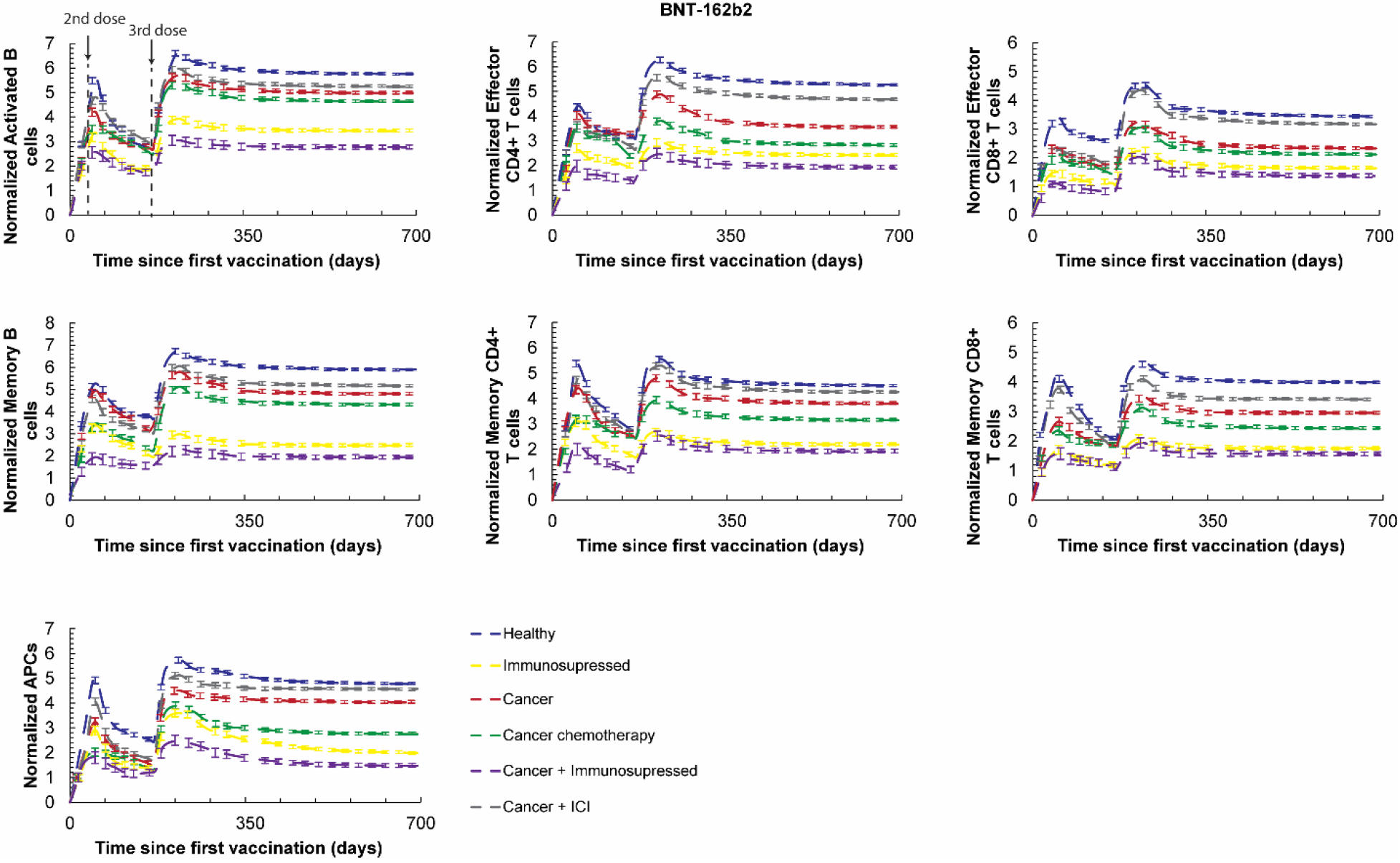
Vaccination induced immunity following the BNT-162b2a vaccine in different patient groups. Levels of B cell, CD4^+^, CD8^+^ T cell and antigen presenting cells (APCs) are depicted for a period of 100 weeks following initial vaccination and a booster dose 6 months later. Values are normalized to the initial value of the corresponding naïve cell type. Error bars present the standard error for all values of model parameters considered accounting for interpatient heterogeneity. The initial values of each naïve cell type are: immature dendritic cells 10^3^ [cells], Naïve CD4+ T cells 10^3^ [cells], Naïve CD8+ T cells 10^3^ [cells], Naïve B cells 10^3^ [cells].

### Vaccine induced protection against severe disease caused by viral variants

Next, we investigated the effects of new variants of the virus in patients on immunosuppression who had received a booster dose. Given that initial model validation was performed when the delta variant was dominant, we consider baseline values of virus infection (Table S2) to be related to the delta variant, which was treated as the control. Our data on infection after vaccination are consistent with the reduced efficacy of a 2 dose regimen for Omicron sub variants BA.1, BA.2, BA.4/5, BA.2 L452Q and BA.2 S704L. However, addition of a booster dose enhances the antibody response against these variants to similar levels seen with the 2 dose regimen with a delta infection. We modulated several parameters in the model to mimic variants that might differ in viral fitness and immune evasion. These include: the affinity of virus for ACE2 (i.e., binding to and detachment from ACE2), the rate of virus internalization into host cells, the replication rate of the internalized virus, the release rate of the virus from infected cells, the clearance rate of viral particles by antibodies, and the activation of immune cells by virus (antigenicity). In our analysis, we accounted separately for variations in each of these parameters either by changing the pertinent rates by 10-fold (mild mutation) or by 100-fold (severe mutation). Exposure to virus was considered to take place 6 months after the booster dose.

To examine the clinical course following a booster dose and then infection with a variant virus, we compared the predicted values for viral load, coagulation/microthrombi formation in the lungs, oxygen saturation levels and the concentration of memory CD4^+^, CD8^+^ and B cells following a booster dose. Results for the BNT-162b2a vaccine are shown in Fig 4. Fig. S3 and S4 present the corresponding results for the mRNA-1273 and Ad26.COV.S vaccines. We find that variants capable of escaping vaccine induced immunity can lead to more severe infections characterized by increased viral load, more microthrombi formation and lower arterial oxygen saturation. This becomes more evident for severe mutations and for patients who received a vector vaccine (Fig 4B and Fig. S3, S4). Furthermore, model predictions indicate a decrease in CD4^+^ and CD8^+^ T cells following infection by a variant in agreement with recent clinical data for the Omicron variant(39). The variants that produced the worst clinical metrics were those with reduced immunogenicity, increased ACE2 binding, and decreased antibody-virus binding. Parameters associated with virus replication, internalization and exocytosis had relatively less effect on clinical trajectories. These data are consistent with recent reports that even after a booster dose, infection with novel omicron subvariants BA.4 and BA.5 result in lower levels of neutralizing antibodies compared to earlier variants(40).

**Figure 4.**
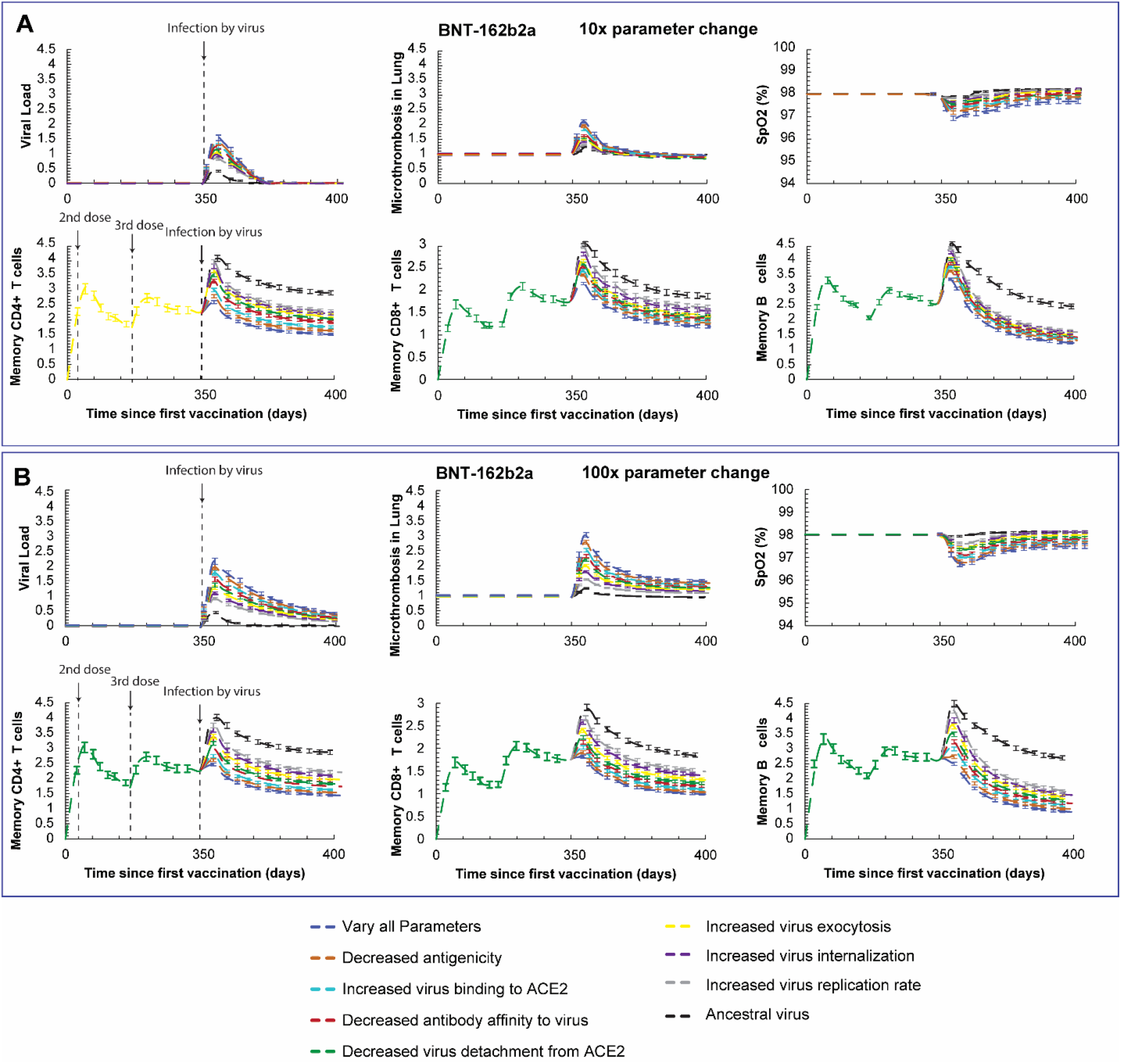
Vaccine induced protection against severe disease caused by viral variants with mild (A) or severe (B) mutations. Predictions of the viral load, coagulation/microthrombi formation in the lungs, oxygen saturation (SpO2) and the concentration of the memory CD4^+^, CD8^+^ and B cells for breakthrough infections after BNT-162b2a vaccination and booster dose. Virus infection was assumed to take place 6 months after the booster dose. The legend lists the parameters related to viral variants that have been varied. Normalized cell values are calculated by division with the initial value of the corresponding naïve cell type. The initial values of each naïve cell type are: immature dendritic cells 10^3^ [cells], Naïve CD4+ T cells 10^3^ [cells], Naïve CD8+ T cells 10^3^ [cells], Naïve B cells 10^3^ [cells]. The simulations were performed for mild mutations (10-fold change in parameters, A) and more severe mutations (100-fold changes in parameters, B).

We further extended our analysis to other phenotypes, including healthy and older individuals and repeated simulations for variants with increased ACE2 binding and increased replication rate (Fig 5&6 and Fig. S5-8). The decrease in memory T cells and in B cells following viral infection is found in all phenotypes but it is more prominent in immunosuppressed and older individuals compared to young healthy people. This is particularly evident for severe mutations affecting ACE2 binding affinity where the levels of memory T cells and B cells drop by more than 50% compared to the corresponding values of the healthy individuals. The decrease in the memory cells population following infection by a variant is attributed to the fact that the variant can more effectively infect endothelial and epithelial cells that causes an increase in IFNγ, which in turn decreases the concentration of T cells and B cells compared to the ancestral virus. Our results demonstrate that changes in viral parameters of fitness and immune escape may result in more severe infections not only on immunosuppressed patients but even for healthy individuals or older individuals not on immunosuppressive therapies.

**Figure 5.**
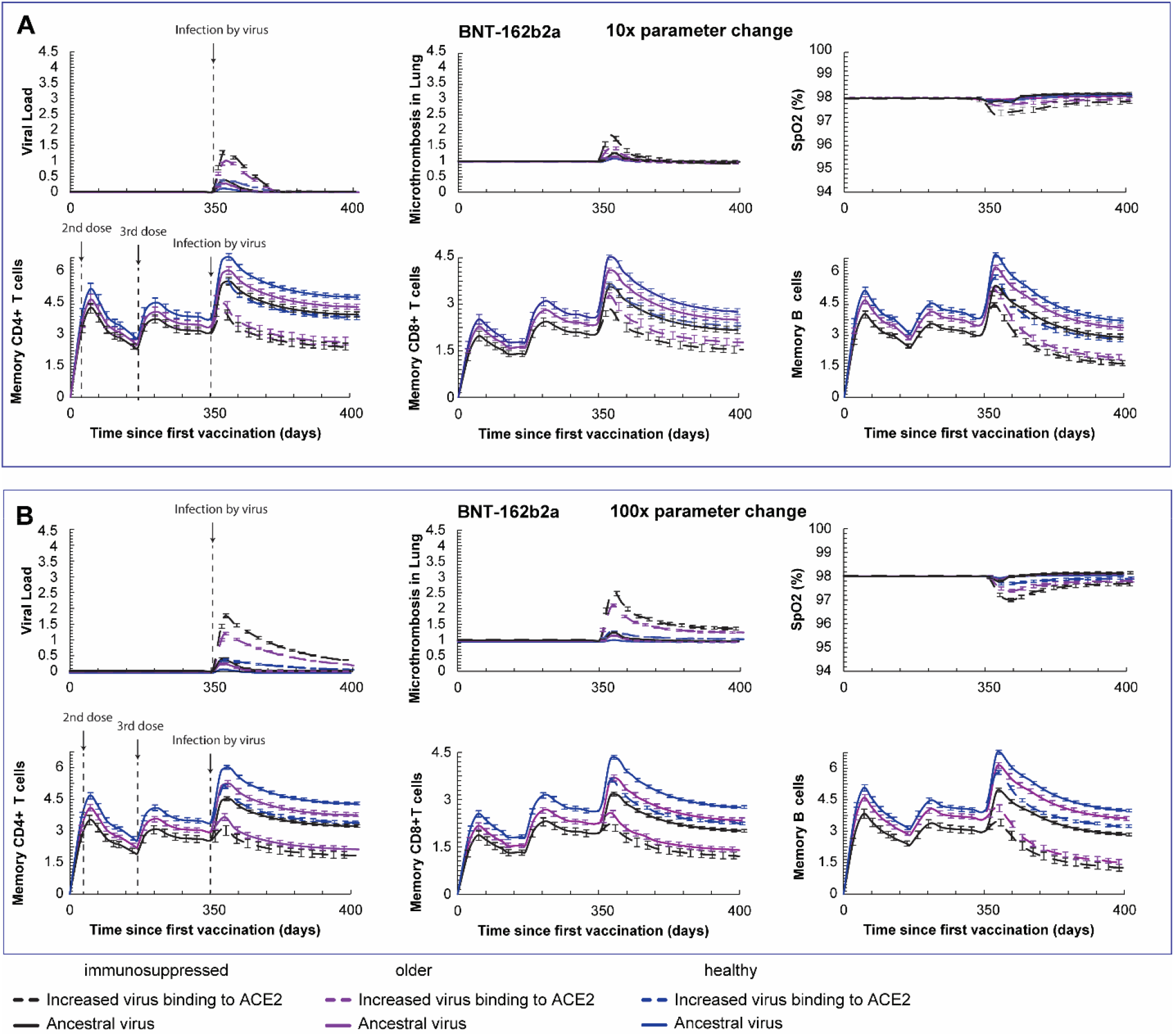
Vaccine (BNT-162b2a) induced protection against severe disease caused by viral variants with mild (A) or severe (B) mutations in ACE2 binding for healthy, older and immunosuppressed. Predictions of the viral load, coagulation/microthrombi formation in the lungs, oxygen saturation (SpO2) and the concentration of the memory CD4^+^, CD8^+^ and B cells for breakthrough infections after BNT-162b2a vaccination and booster dose. Virus infection was assumed to take place 6 months after the booster dose. Normalized cell values are calculated by division with the initial value of the corresponding naïve cell type. The initial values of each naïve cell type are: immature dendritic cells 10^3^ [cells], Naïve CD4+ T cells 10^3^ [cells], Naïve CD8+ T cells 10^3^ [cells], Naïve B cells 10^3^ [cells]. The simulations were performed for mild mutations (10-fold change in parameter, A) and more severe mutations (100-fold change in parameter, B).

**Figure 6.**
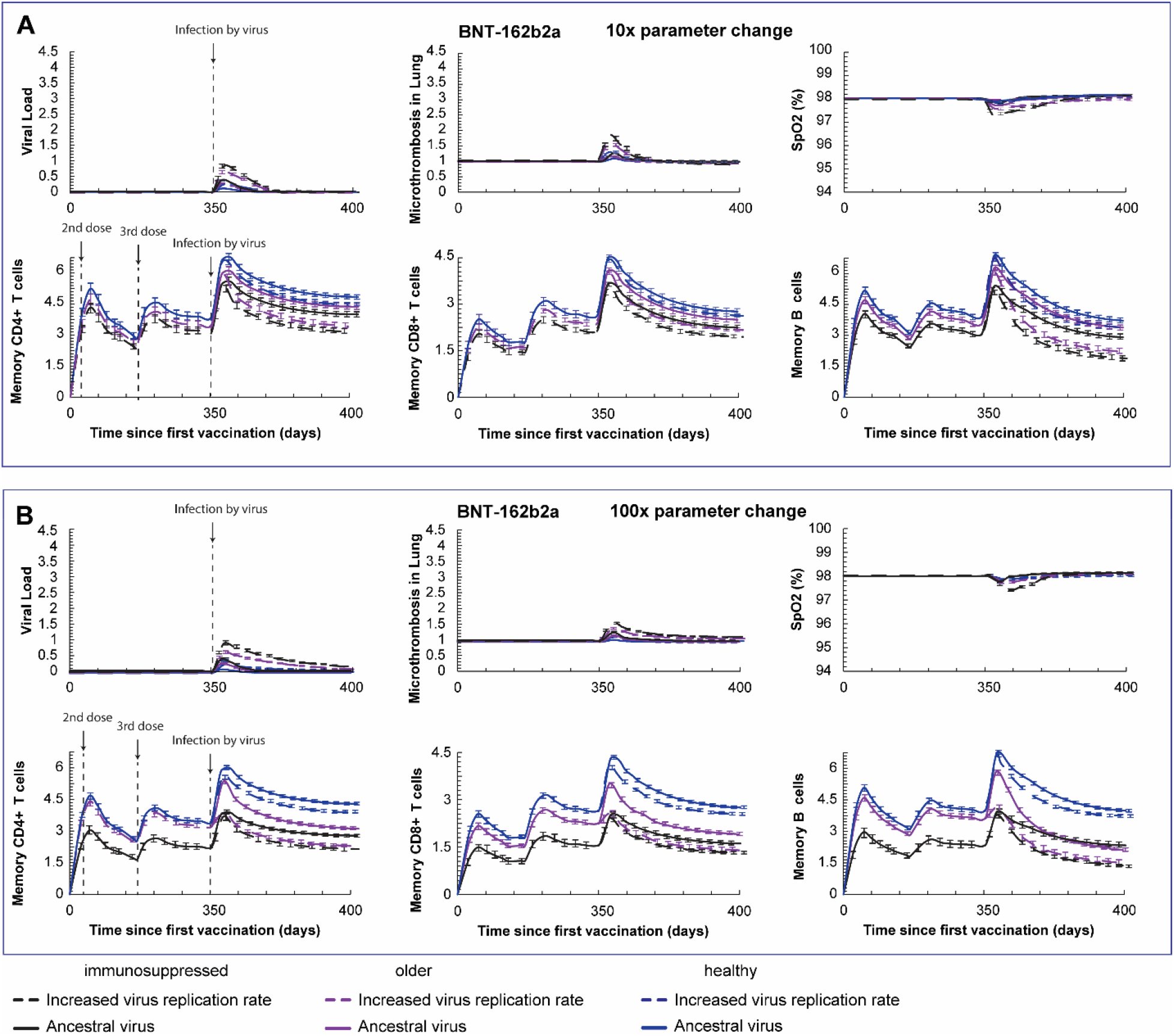
Vaccine (BNT-162b2a) induced protection against severe disease caused by viral variants with mild (A) or severe (B) mutations in virus replication ability for healthy, older and immunosuppressed. Predictions of the viral load, coagulation/microthrombi formation in the lungs, oxygen saturation (SpO2) and the concentration of the memory CD4^+^, CD8^+^ and B cells for breakthrough infections after BNT-162b2a vaccination and booster dose. Virus infection was assumed to take place 6 months after the booster dose. Normalized cell values are calculated by division with the initial value of the corresponding naïve cell type. The initial values of each naïve cell type are: immature dendritic cells 10^3^ [cells], Naïve CD4+ T cells 10^3^ [cells], Naïve CD8+ T cells 10^3^ [cells], Naïve B cells 10^3^ [cells]. The simulations were performed for mild mutations (10-fold change in parameter, A) and more severe mutations (100-fold change in parameter, B).

### Effect of intervals between booster doses on viral infection and immunity

We next simulated different time intervals between a first and a second booster dose for both mRNA and vector vaccines on viral infection and cell immunity. We first repeated the simulations of viral infection by a variant (with severe mutation) for patients on immunosuppression, varying the time interval of the first booster dose from 3 to 6 months. As before, exposure to virus was considered to take place 6 months after the booster dose. Model predictions are depicted in Fig 7 for the BNT-162b2a vaccine and in Fig. S9 and S10, for the mRNA-1273 and AD26.COV2.S vaccines, respectively. Interestingly, the model predicts a more than 60% reduction in viral load and microthrombi formation (for the mRNA vaccines) and higher levels of memory CD4^+^ and CD8^+^ T cells when the first booster is administered after a 6-month interval compared to a 3-month interval. In the mathematical model, the activation of CD4^+^/CD8^+^ T cells and B cells depends largely on the [PD1-PDL1] complex. Specifically, the lower the levels of [PD1-PDL1], the more the CD4^+^/CD8^+^ T cells and B cells get activated, resulting in higher concentrations of the corresponding memory cells. The temporal variation of [PD1-PDL1] varying the time to 1^st^ booster from 3 to 8 months is depicted in Fig. S13. The results suggest that for a 6-month time interval to 1^st^ booster dose, the concentration of [PD1-PDL1] has lower values, which results in increased production of all types of memory cells (Fig. 7). Recent publications have attributed the improvement in the antibody response with delayed booster dosing to greater affinity maturation (20), but our model does not account for this mechanism. A second booster vaccination can further increase immunity in patients on immunosuppression although the levels of memory CD4^+^ and CD8^+^ T cells and B cells are lower than those of healthy individuals after one booster vaccination (Fig 8 for BNT-162b2a, Fig S11 and S12 for mRNA-1273 and AD26.COV2.S, respectively). However, model predictions suggest that the time interval of the second booster dose does not strongly affect cell immunity or the severity of viral infection. The second booster vaccination seems to be protective for patients on immunosuppression regardless of when it is given in a period 3-6 months following the first booster. The same conclusion was reached when we repeated simulations for all possible combinations varying the time to 1^st^ and 2^nd^ booster from 3 to 8 months (Fig 8 and Fig S14). In Fig. 8 model predictions for the maximum values of viral load and lung microthrombi and for the minimum values of SpO2 are shown as a function of the times to the 1^st^ and 2^nd^ booster, whereas Fig S14 depicts the equilibrium values after viral injection for the memory CD4^+^ and CD8^+^ T cells and B cells. Our results show that the best interval to the first and to the second booster is 6 months for all vaccines.

**Figure 7.**
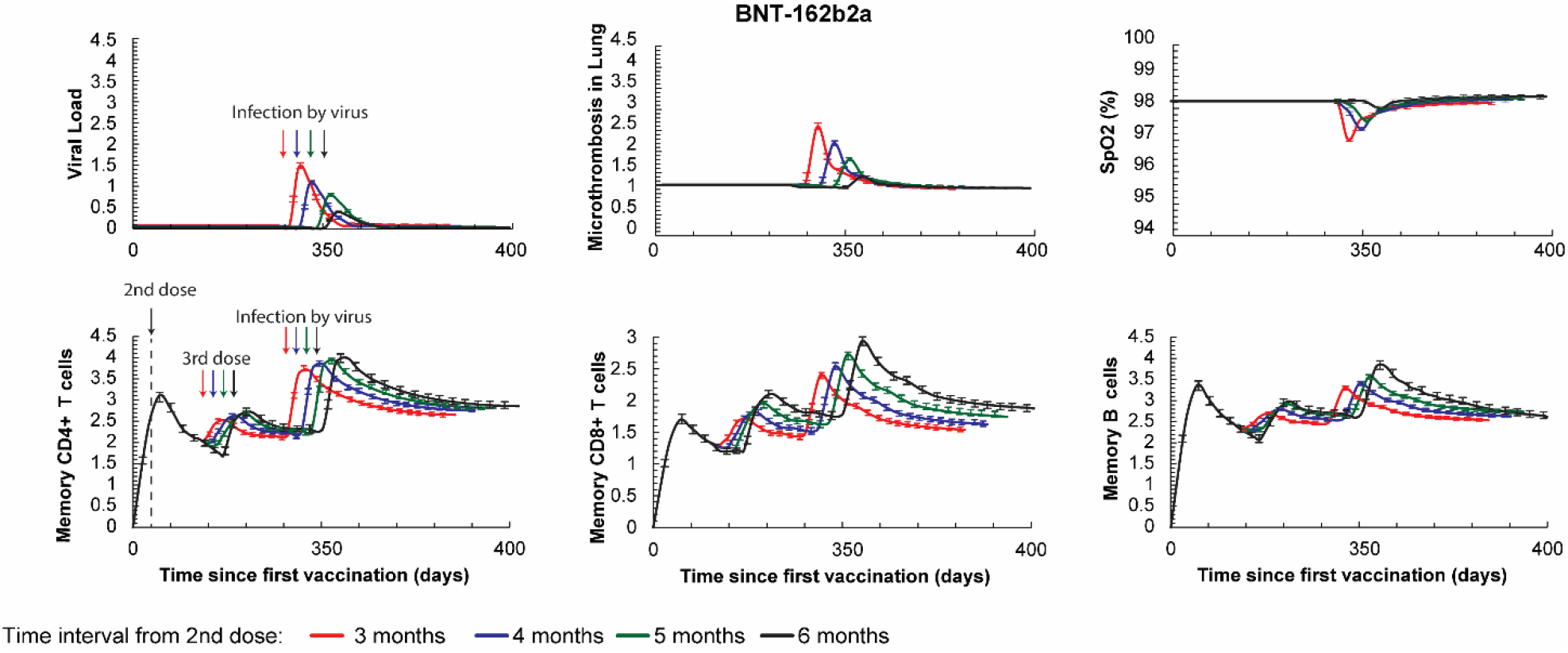
Model predictions for the effect of the timing of the first booster on vaccine induced protection. Predictions of the viral load, coagulation/microthrombi formation in the lungs, oxygen saturation (SpO2) and the concentration of the memory CD4+, CD8+ and B cells for breakthrough infections after BNT-162b2a vaccination for intervals between the 2^nd^ and 3^rd^ dose of 3, 4, 5 and 6 months. Virus infection was assumed to take place 6 months after the booster dose. Normalized cell values are calculated by division with the initial value of the corresponding naïve cell type. Error bars present the standard error for all values of model parameters considered in order to account for interpatient heterogeneity. The initial values of each naïve cell type are: immature dendritic cells 10^3^ [cells], Naïve CD4+ T cells 10^3^ [cells], Naïve CD8+ T cells 10^3^ [cells], Naïve B cells 10^3^ [cells].

**Figure 8.**
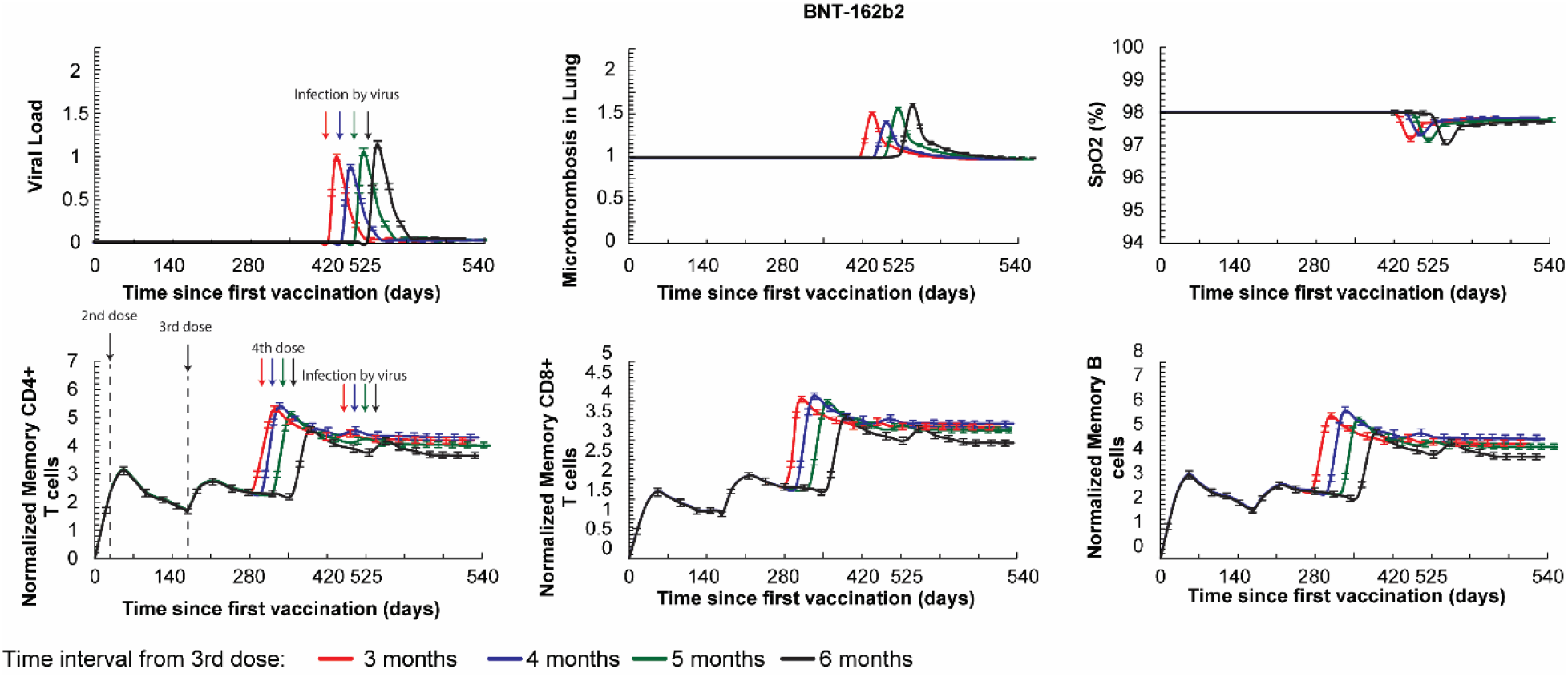
Model predictions for the effect of the timing of the second booster on vaccine induced protection. Predictions of the viral load, coagulation/microthrombi formation in the lungs, oxygen saturation (SpO2) and the concentration of the memory CD4+, CD8+ and B cells for breakthrough infections after BNT-162b2a vaccination for intervals between the 3^rd^ and 4^th^ dose of 3, 4, 5 and 6 months. Virus infection was assumed to take place 6 months after the booster dose. Normalized cell values are calculated by division with the initial value of the corresponding naïve cell type. Error bars present the standard error for all values of model parameters considered in order to account for interpatient heterogeneity. The initial values of each naïve cell type are: immature dendritic cells 10^3^ [cells], Naïve CD4+ T cells 10^3^ [cells], Naïve CD8+ T cells 10^3^ [cells], Naïve B cells 10^3^ [cells].

**Figure 9.**
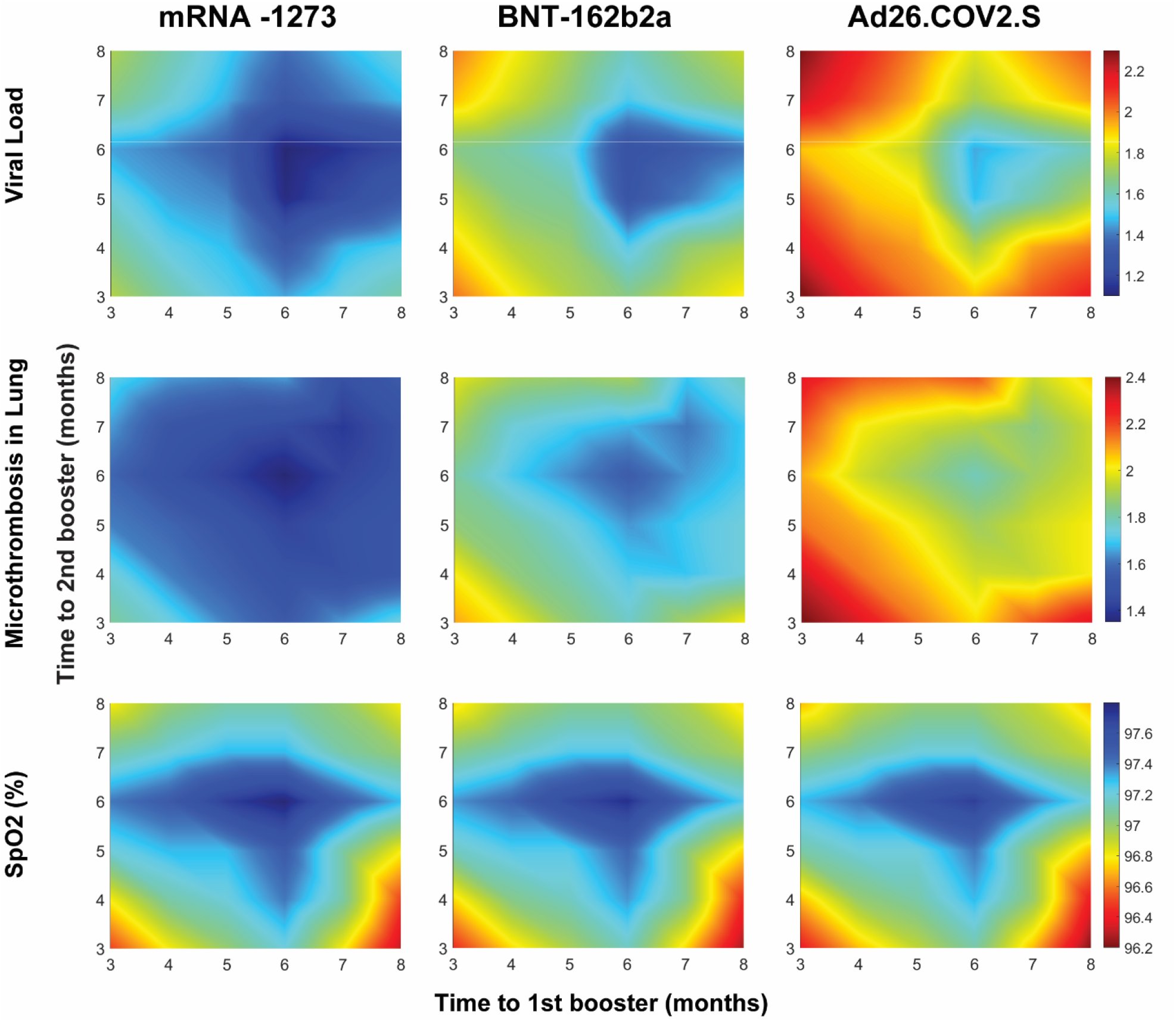
Phase diagrams of maximum viral load and microthrombosis in the lung and minimum oxygen saturation (SpO2) levels caused by viral infection as a function of the time to 1^st^ and 2^nd^ booster vaccinations for all types of vaccines and for immunosuppressed individuals. Virus infection was assumed to take place 6 months following 2^nd^ booster dose.

Finally, we sought to exploit the incorporation of both the humoral and the cellular immune responses in our model to explore the relative importance of these two mechanisms of vaccine-induced immunity. We repeated simulations of virus infection following a booster dose for older individuals having received the BNT-162b2a vaccine but with a zeroing out of the production of either T cell or the B cell. The results show that the effect of setting to zero the production of B cells results in higher viral load and microthrombi formation and lower SpO2 compared to the effect of setting to zero the production of T cells (Fig. S15). These data are consistent with a greater importance of the humoral response to vaccine induced immunity.

## Discussion

Vaccination is an effective measure against SARS-CoV-2 infection and the development of COVID-19, but its effectiveness may wane over time and can be limited by virus or patient specific factors. Here, we developed a mathematical framework incorporating the known mechanisms of vaccination-induced immunity, SARS-CoV-2 infection and COVID-19 pathophysiology to investigate the effect of booster doses on humoral and cell mediated immunity and severity of clinical course following infection with ancestral and variant viruses. Focusing only on homologous vaccinations (i.e., no mixing of different vaccine types), we confirmed the known advantages of mRNA vaccines over vector vaccines and replicated the decreased immune response 6 months after vaccination. A booster dose of both mRNA vaccines can induce a robust enhancement of both antibody levels and numbers of pertinent types of adaptive immune cells, which is predicted to provide sufficient protection for more than one year in healthy patients. However, our model suggests that for immunosuppressed people or patients with cancer receiving an immunosuppressive treatment (e.g., chemotherapy or B cell depletion treatment), the booster effect may wane, and perhaps could be considered on a more frequent basis.

Our analysis reinforces current CDC guidance that all individuals benefit from a booster dose after their vaccination with mRNA vaccines, and immunosuppressed and older individuals should be further considered for more frequent booster doses. For the Ad26.COV2.S vector vaccine, additional booster doses should be considered for all individuals.

Patients with cancer and/or immunosuppression are at higher risk of severe outcomes from COVID-19, and are notably at higher risk of failing to clear the virus. For this reason, it is highly important to ensure protection in this patient population. Our model simulations suggest that these groups warrant tailored approaches to vaccination, in particular, considering additional doses beyond a first booster dose, prioritization for next generation vaccine approaches and consideration of additional prophylactic measures. As shown in our simulations with the severe mutation variant (Fig 7, S9 and 10), the timing of the booster dose affects the induction of immunity. Therefore, it is also important to consider that different patients (with or without cancer) will need different timing of vaccination.

Our model is also able to make predictions on the impact of novel SARS-CoV-2 variants on COVID-19 infection. The simulations suggest that variants with enhanced target cell binding, reduced antibody binding or reduced antigenicity/immunogenicity will present the greatest clinical challenges, even resulting in severe infections in healthy vaccinated individuals. Such variants reduce the protection afforded by the current vaccines, requiring additional booster doses or new vaccine formulations. We anticipate that this model could be adapted to fit other variants with differing features of immune escape or viral fitness. While adaptations, such as enhanced infectivity and immune evasion, are similar to those reported for Omicron, we make no claim that we model any specific variant – rather we seek to model the types of viral changes which might alter vaccine response. When long-term data from the response of the vaccines to viral variants, including the Omicron variant, become available, our model will be able to be compared to such clinical data.

There are a number of limitations to the analysis presented here. It depends upon a large number of parameters whose values are not explicitly known for the SARS-CoV-2 virus. Here, and in our previous research, we have performed a robust sensitivity analysis of all major parameters of the model and have shown that model predictions might change quantitatively, but our conclusions are not affected qualitatively(27, 41). In our approach, parameters are altered according to mechanistic hypotheses on the interaction between model components and outcome, which allows for direct testing of a hypothesis but at the cost of oversimplification of patient diversity and comorbidities.

Our analysis is also limited in studying the effects of cancer and immunosuppressive agents. It is clear there is large variability in the effects of cancer types (e.g., hematologic cancers vs. solid tumors) on antibody production after vaccination (13). In our analysis, we did not distinguish solid from liquid tumors from the immunosuppression point of view. Similarly, different chemotherapy or immunotherapy strategies are likely to produce differential effects on immune cell populations and antibody response that may meaningfully affect antibody levels. It is also known that specific chemotherapies are immunostimulatory(42), whereas we have considered only those that are immunosuppressive. However, our clinical validations sets were largely pan-cancer studies without granular detail on specific therapies, and thus we developed a more general model that can be refined as more disease-specific data become available.

In conclusion, utilizing existing clinical data and mathematical modelling, we provide predictions for vaccine efficacy over time frames that greatly exceed available clinical data. Our models predict that variants that reduce immunogenicity, increase ACE2 binding, and decrease antibody-virus binding pose the greatest risk, but tailoring vaccine booster timing may optimize their deployment further. These results could help inform the timing of booster vaccinations in individuals of different phenotypes and comorbidities, as well as for novel viral variants. As we approach an endemic phase of SARS-CoV-2, a rational approach to vaccine booster utilization may help ensure equitable access to vaccines and help prevent further outbreaks and development of new variants.

### Model Description

A detailed description of model equations, the description and values of model parameters, and the solution strategy are provided in Supplementary Materials.

## Data Availability

Data availability. All data supporting the findings of this study are available in the paper and the Supplementary Information.
Code availability. The COMSOL code will become available in Zenodo upon acceptance of the manuscript.

## Data availability

All data supporting the findings of this study are available in the paper and the Supplementary Information.

## Code availability

The COMSOL code will become available in Zenodo upon acceptance of the manuscript.

## Acknowledgments

Rakesh Jain’s research is supported by R01-CA259253, R01-CA208205, R01-NS118929, U01-CA261842, and U01-CA 224348, Outstanding Investigator Award R35-CA197743 and grants from the National Foundation for Cancer Research, Jane’s Trust Foundation, American Medical Research Foundation and Harvard Ludwig Cancer Center. Lance Munn’s research is supported by R01-CA2044949. Triantafyllos Stylianopoulos’s research is supported by the European Research Council ERC-2019-CoG-863955. Chrysovalantis Voutouri is supported by Marie Skłodowska Curie Actions Individual Fellowship Global Horizon 2020 MSCA-IF-GF-2020-101028945.

## Notes

**Competing Interest Statement:** J.F.G. has served as a compensated consultant or received honoraria from Bristol-Myers Squibb, Genentech/Roche, Ariad/Takeda, Loxo/Lilly, Blueprint, Oncorus, Regeneron, Gilead, Moderna, Mirati, AstraZeneca, Pfizer, Novartis, iTeos, Nuvalent, Karyopharm, Beigene, Silverback Therapeutics, Merck, and GlydeBio; research support from Novartis, Genentech/Roche, and Ariad/Takeda; institutional research support from Bristol-Myers Squibb, Tesaro, Moderna, Blueprint, Jounce, Array Biopharma, Merck, Adaptimmune, Novartis, and Alexo; and has an immediate family member who is an employee with equity at Ironwood Pharmaceuticals. L.L.M. owns equity in Bayer AG and is a consultant for SimBiosys. R.K.J. received consultant fees from Elpis, Innocoll, SPARC, SynDevRx; owns equity in Accurius, Enlight, Ophthotech, SynDevRx; and serves on the Boards of Trustees of Tekla Healthcare Investors, Tekla Life Sciences Investors, Tekla Healthcare Opportunities Fund, Tekla World Healthcare Fund; and received a grant from Boehringer Ingelheim. Neither any reagent nor any funding from these organizations was used in this study. Other co-authors have no conflict of interests to declare.

### Competing Interest Statement

Competing Interest Statement: J.F.G. has served as a compensated consultant or received honoraria from Bristol-Myers Squibb, Genentech/Roche, Ariad/Takeda, Loxo/Lilly, Blueprint, Oncorus, Regeneron, Gilead, Moderna, Mirati, AstraZeneca, Pfizer, Novartis, iTeos, Nuvalent, Karyopharm, Beigene, Silverback Therapeutics, Merck, and GlydeBio; research support from Novartis, Genentech/Roche, and Ariad/Takeda; institutional research support from Bristol-Myers Squibb, Tesaro, Moderna, Blueprint, Jounce, Array Biopharma, Merck, Adaptimmune, Novartis, and Alexo; and has an immediate family member who is an employee with equity at Ironwood Pharmaceuticals. L.L.M. owns equity in Bayer AG and is a consultant for SimBiosys. R.K.J. received consultant fees from Elpis, Innocoll, SPARC, SynDevRx; owns equity in Accurius, Enlight, Ophthotech, SynDevRx; and serves on the Boards of Trustees of Tekla Healthcare Investors, Tekla Life Sciences Investors, Tekla Healthcare Opportunities Fund, Tekla World Healthcare Fund; and received a grant from Boehringer Ingelheim. Neither any reagent nor any funding from these organizations was used in this study. Other co-authors have no conflict of interests to declare.

### Funding Statement

Rakesh Jains research is supported by R01-CA259253 R01-CA208205 R01-NS118929 U01-CA261842 and U01-CA 224348 Outstanding Investigator Award R35-CA197743 and grants from the National Foundation for Cancer Research Jane's Trust Foundation American Medical Research Foundation and Harvard Ludwig Cancer Center. Lance Munn's research is supported by R01-CA2044949. Triantafyllos Stylianopoulos's research is supported by the European Research Council ERC-2019-CoG-863955. Chrysovalantis Voutouri is supported by Marie Skłodowska Curie Actions Individual Fellowship Global Horizon 2020 MSCA-IF-GF-2020-101028945.

### Author Declarations

Mass General Brigham Institutional Review Board of Massachusetts General Hospital gave ethical approval for this work

