## Supplementary material for "Mechanistic model for booster doses effectiveness in healthy, cancer and immunosuppressed patients infected with SARS-CoV-2": Online supplement

**This PDF file includes:**

Supplementary text

Figures S1 to S15 (not allowed for Brief Reports)

Tables S1 to S3 (not allowed for Brief Reports)

SI References

**Supplementary Figures and Tables**

**Mechanistic model for booster doses effectiveness in healthy, cancer and immunosuppressed patients infected with the ancestral or variant SARS-CoV-2**

**Fig. S1**. Vaccination induced immunity caused by the mRNA-1273 vaccine. Levels of B cell, CD4^+^/CD8^+^ T cell and antigen presented cells (APCs) are presented for a period of 100 weeks following initial vaccination and a booster dose 6 months later. Values are normalized to the initial value of the corresponding naïve cell type. The initial values of each naïve cell type are: immature dendritic cells 10^3^ [cells], Naïve CD4+ T cells 10^3^ [cells], Naïve CD8+ T cells 10^3^ [cells], Naïve B cells 10^3^ [cells].

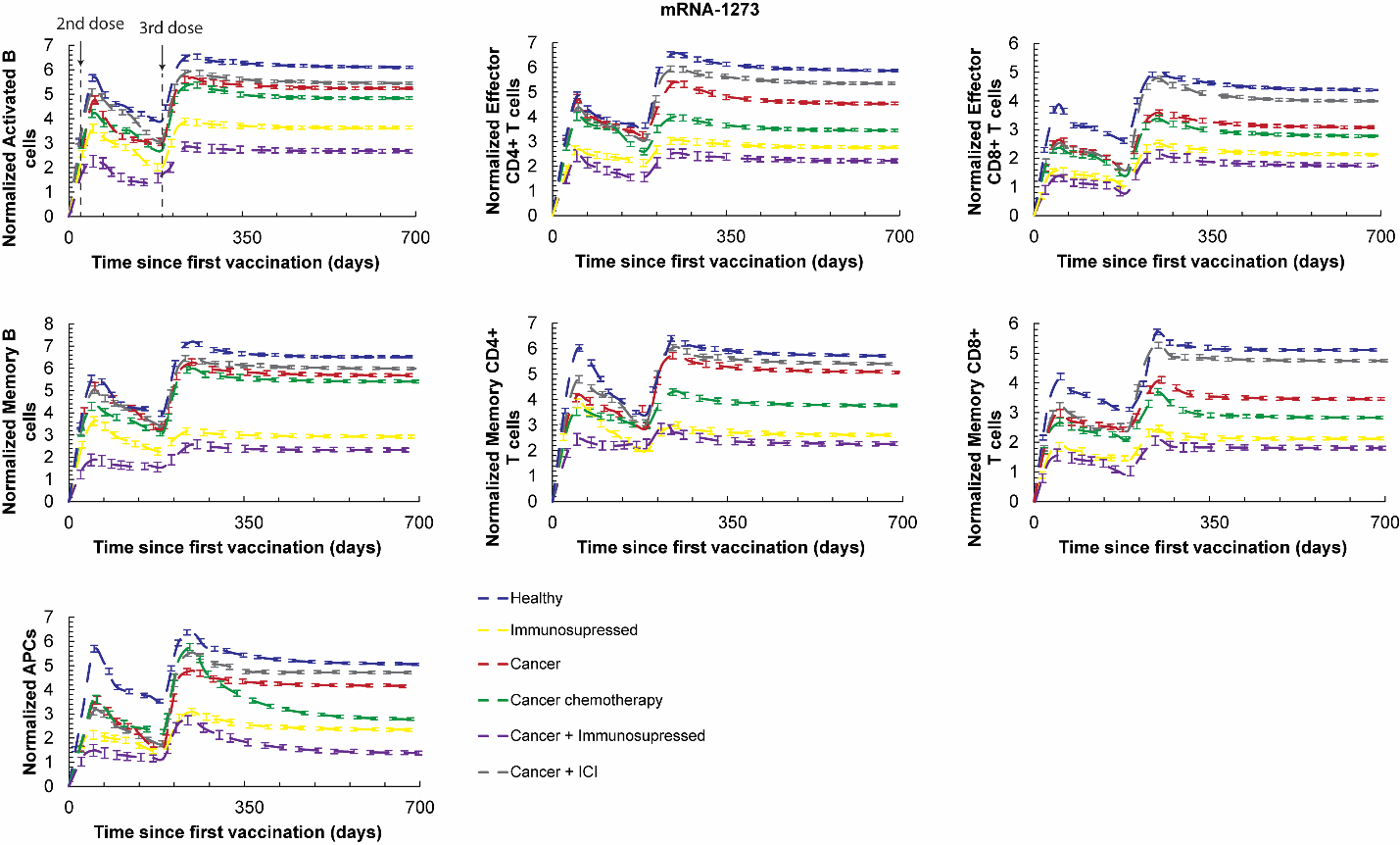

**Fig. S2**. Vaccination induced immunity caused by the Ad26.COV2.S vaccine. Levels of B cell, CD4^+^/CD8^+^ T cell and antigen presented cells (APCs) are presented for a period of 100 weeks following initial vaccination and a booster dose 6 months later. Values are normalized to the initial value of the corresponding naïve cells type. The initial values of each naïve cell type are: immature dendritic cells 10^3^ [cells], Naïve CD4+ T cells 10^3^ [cells], Naïve CD8+ T cells 10^3^ [cells], Naïve B cells 10^3^ [cells].

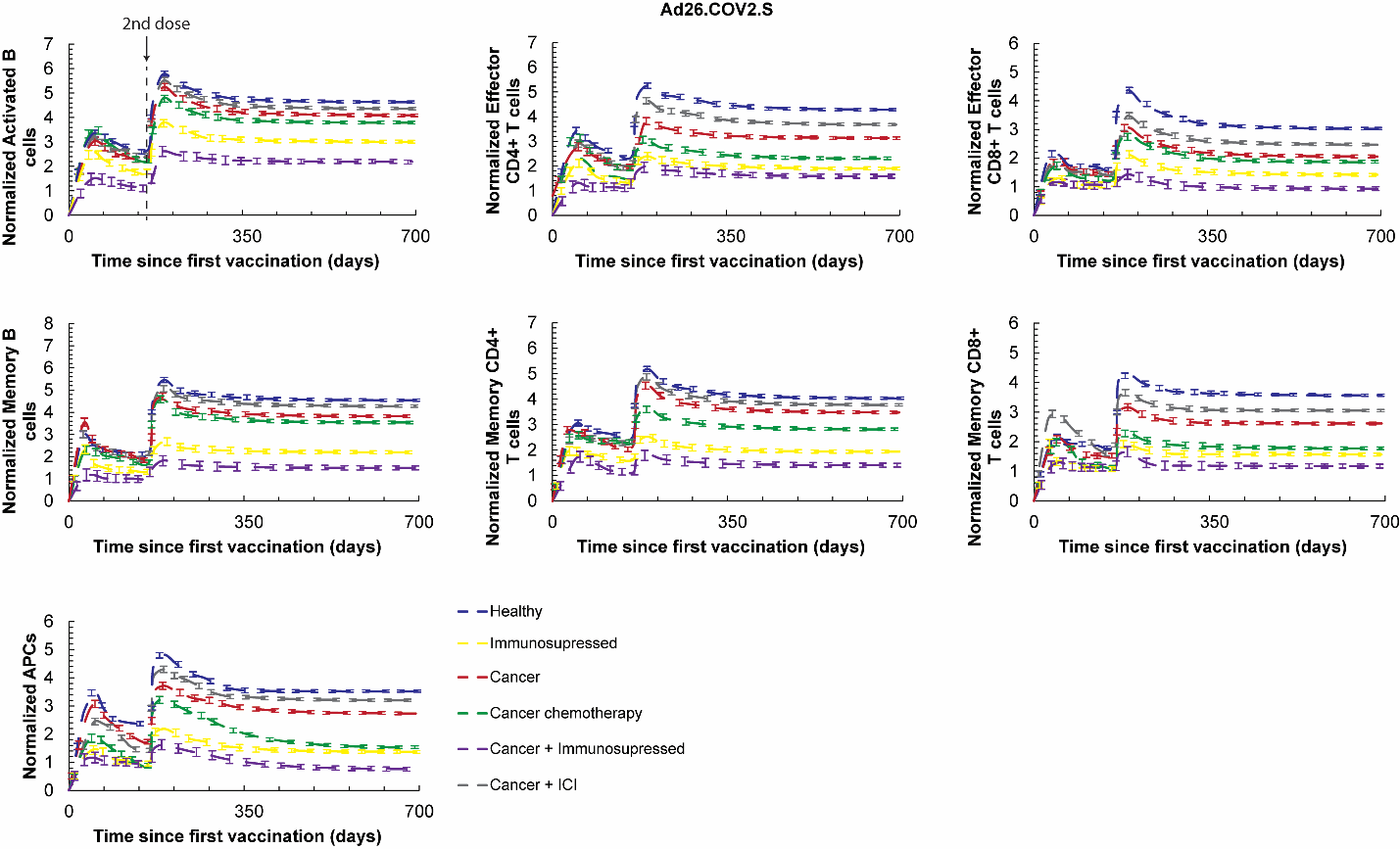

**Fig. S3**. **mRNA-1273 vaccine induced protection against severe disease caused by viral variants with mild (A) or severe (B) mutations.** Predictions of the viral load, coagulation/microthrombi formation in the lungs, oxygen saturation (SpO2) and the concentration of the memory CD4^+^, CD8^+^ and B cells for breakthrough infections after mRNA-1273 vaccination and booster dose. Virus infection was assumed to take place 6 months after the booster dose. The legend lists the parameters related to viral variants that have been varied. Normalized cell values are calculated by division with the initial value of the corresponding naïve cell type. The initial values of each naïve cell type are: immature dendritic cells 10^3^ [cells], Naïve CD4+ T cells 10^3^ [cells], Naïve CD8+ T cells 10^3^ [cells], Naïve B cells 10^3^ [cells]. The simulations were performed for mild mutations (10-fold change in parameters, A) and more severe mutations (100-fold changes in parameters, B).

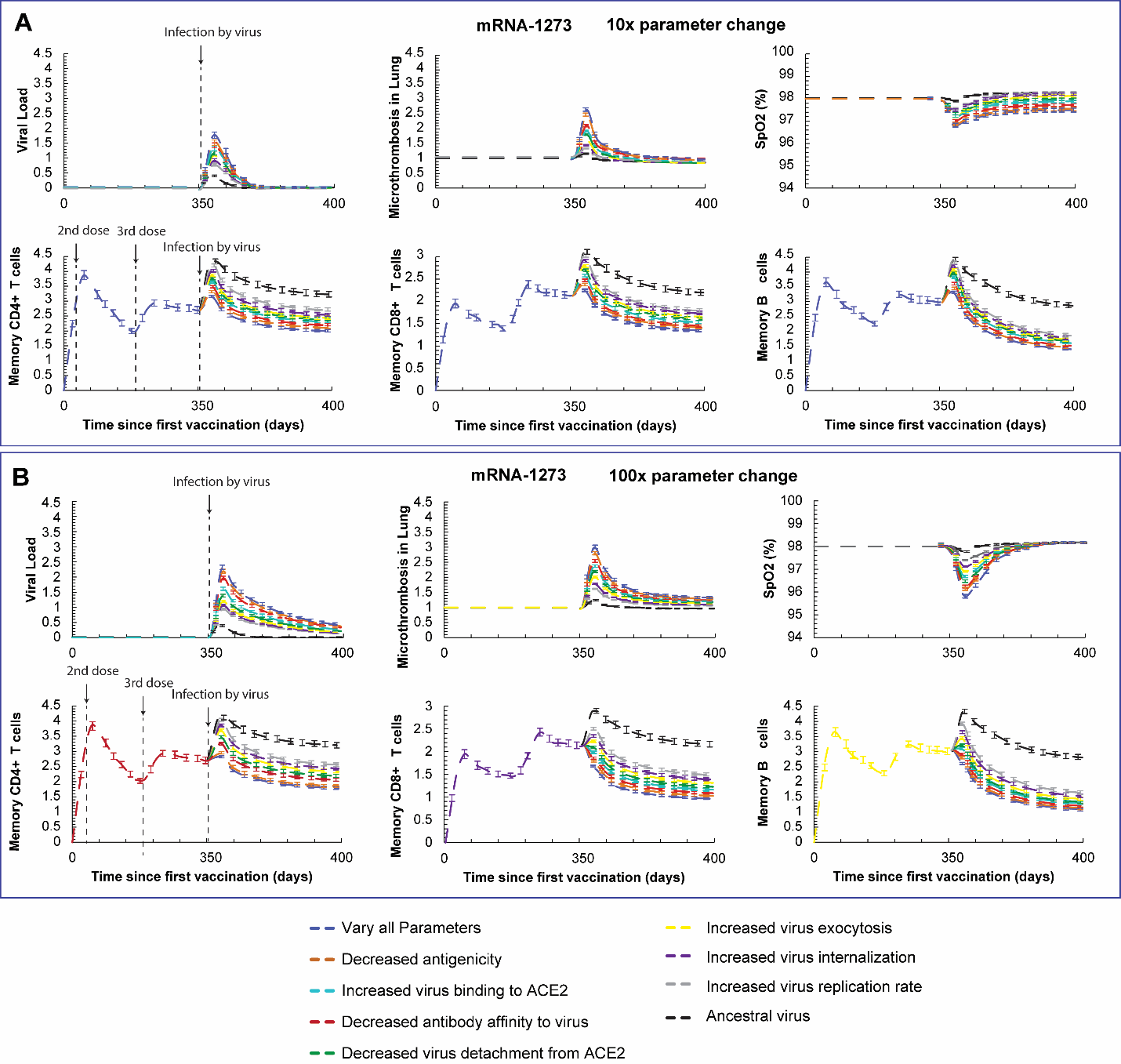

**Fig. S4**. **Ad26.COV2.S vaccine induced protection against severe disease caused by viral variants with mild (A) or severe (B) mutations.** Predictions of the viral load, coagulation/microthrombi formation in the lungs, oxygen saturation (SpO2) and the concentration of the memory CD4^+^, CD8^+^ and B cells for breakthrough infections after Ad26.COV2.S vaccination and booster dose. Virus infection was assumed to take place 6 months after the booster dose. The legend lists the parameters related to viral variants that have been varied. Normalized cell values are calculated by division with the initial value of the corresponding naïve cell type. The initial values of each naïve cell type are: immature dendritic cells 10^3^ [cells], Naïve CD4+ T cells 10^3^ [cells], Naïve CD8+ T cells 10^3^ [cells], Naïve B cells 10^3^ [cells]. The simulations were performed for mild mutations (10-fold change in parameters, A) and more severe mutations (100-fold changes in parameters, B).

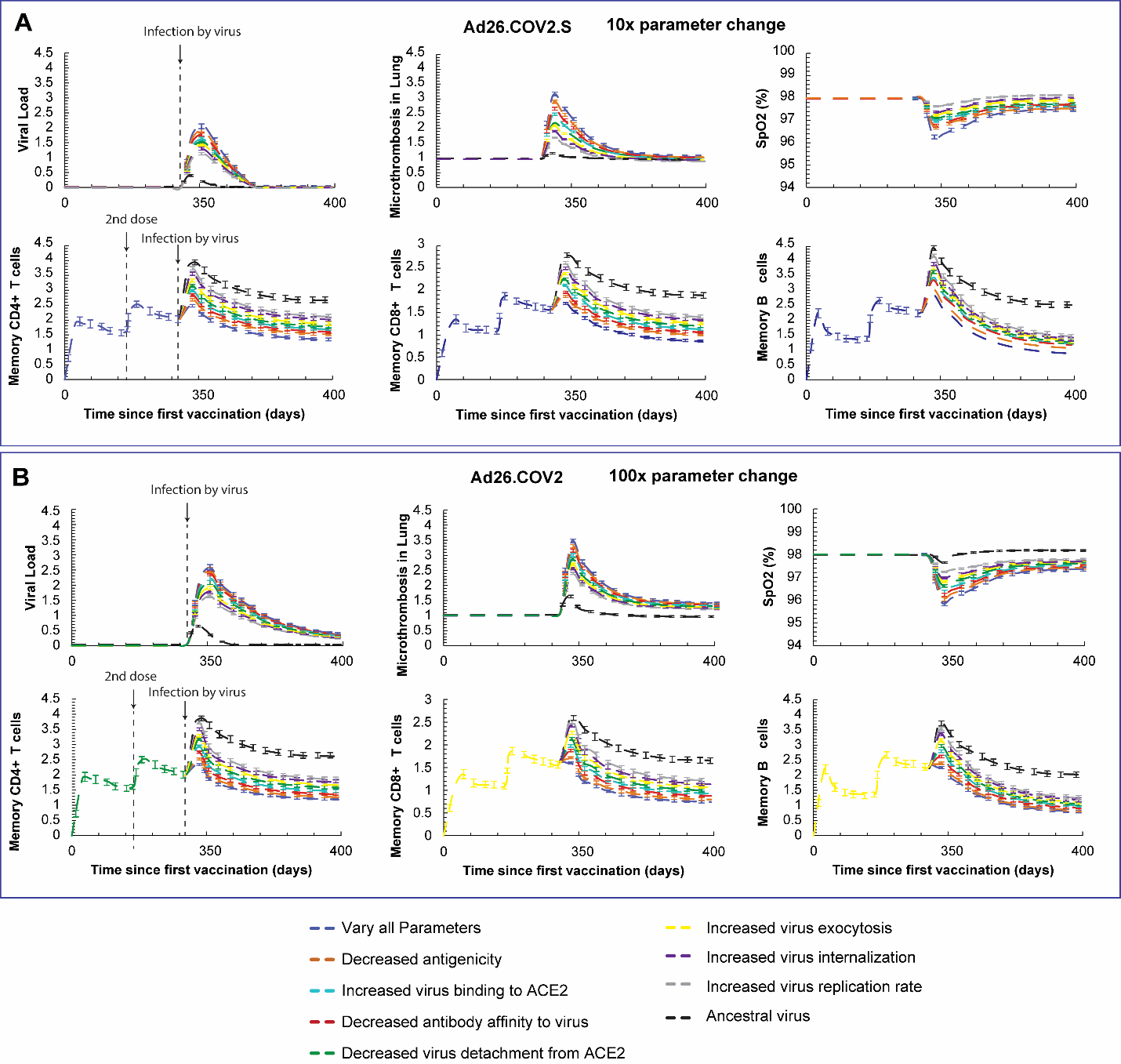

**Fig. S5. Vaccine (mRNA-1273) induced protection against severe disease caused by viral variants with mild (A) or severe (B) mutations in ACE2 binding for healthy, older and immunosuppressed.** Predictions of the viral load, coagulation/microthrombi formation in the lungs, oxygen saturation (SpO2) and the concentration of the memory CD4+, CD8+ and B cells for breakthrough infections after BNT-162b2a vaccination and booster dose. Virus infection was assumed to take place 6 months after the booster dose. Normalized cell values are calculated by division with the initial value of the corresponding naïve cell type. The initial values of each naïve cell type are: immature dendritic cells 103 [cells], Naïve CD4+ T cells 103 [cells], Naïve CD8+ T cells 103 [cells], Naïve B cells 103 [cells]. The simulations were performed for mild mutations (10-fold change in parameter, A) and more severe mutations (100-fold changes in parameter, B).

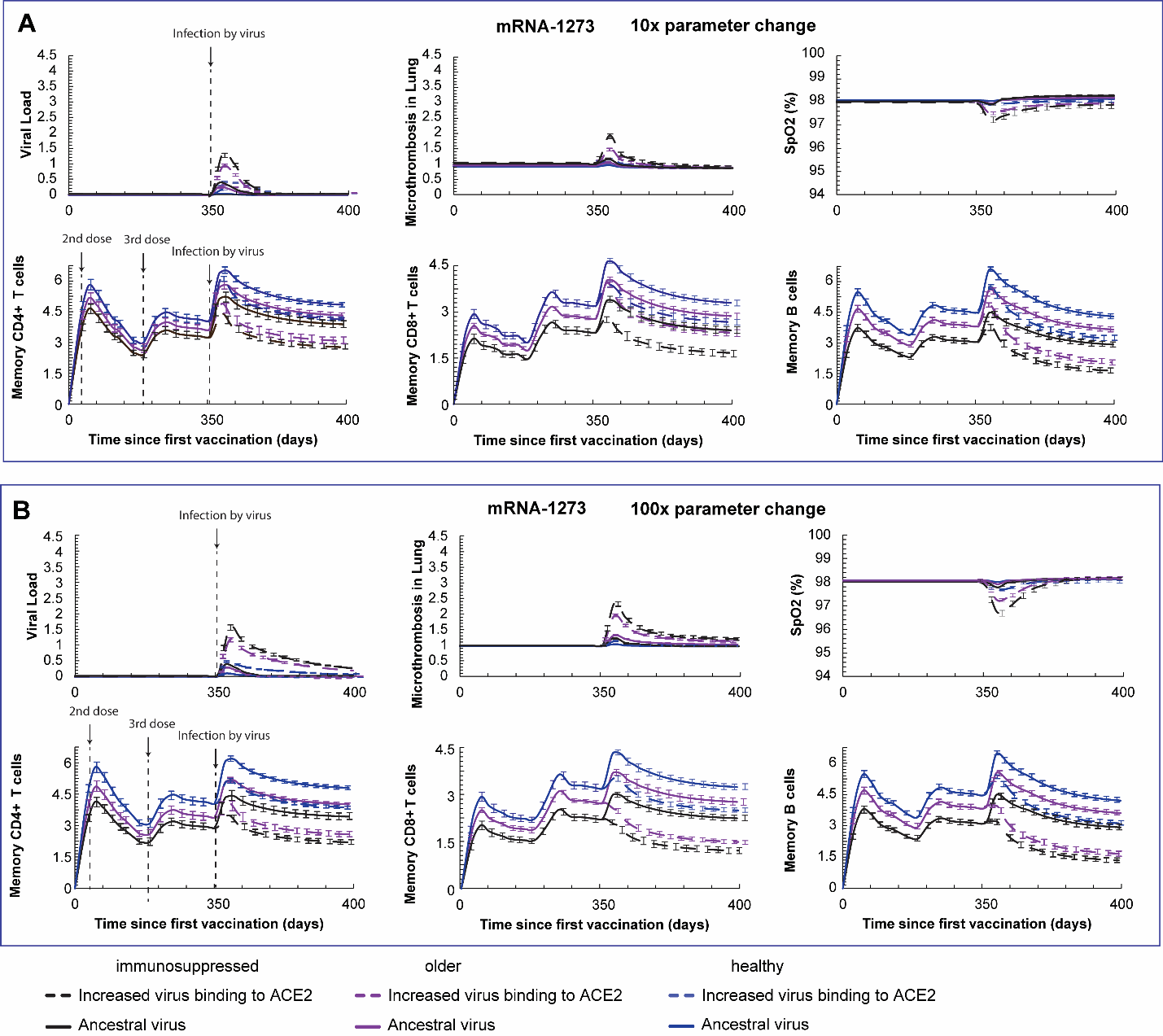

**Fig. S6. Vaccine (Ad26.COV2.S) induced protection against severe disease caused by viral variants with mild (A) or severe (B) mutations in ACE2 binding for healthy, older and immunosuppressed.** Predictions of the viral load, coagulation/microthrombi formation in the lungs, oxygen saturation (SpO2) and the concentration of the memory CD4+, CD8+ and B cells for breakthrough infections after BNT-162b2a vaccination and booster dose. Virus infection was assumed to take place 6 months after the booster dose. Normalized cell values are calculated by division with the initial value of the corresponding naïve cell type. The initial values of each naïve cell type are: immature dendritic cells 103 [cells], Naïve CD4+ T cells 103 [cells], Naïve CD8+ T cells 103 [cells], Naïve B cells 103 [cells]. The simulations were performed for mild mutations (10-fold change in parameter, A) and more severe mutations (100-fold changes in parameter, B).

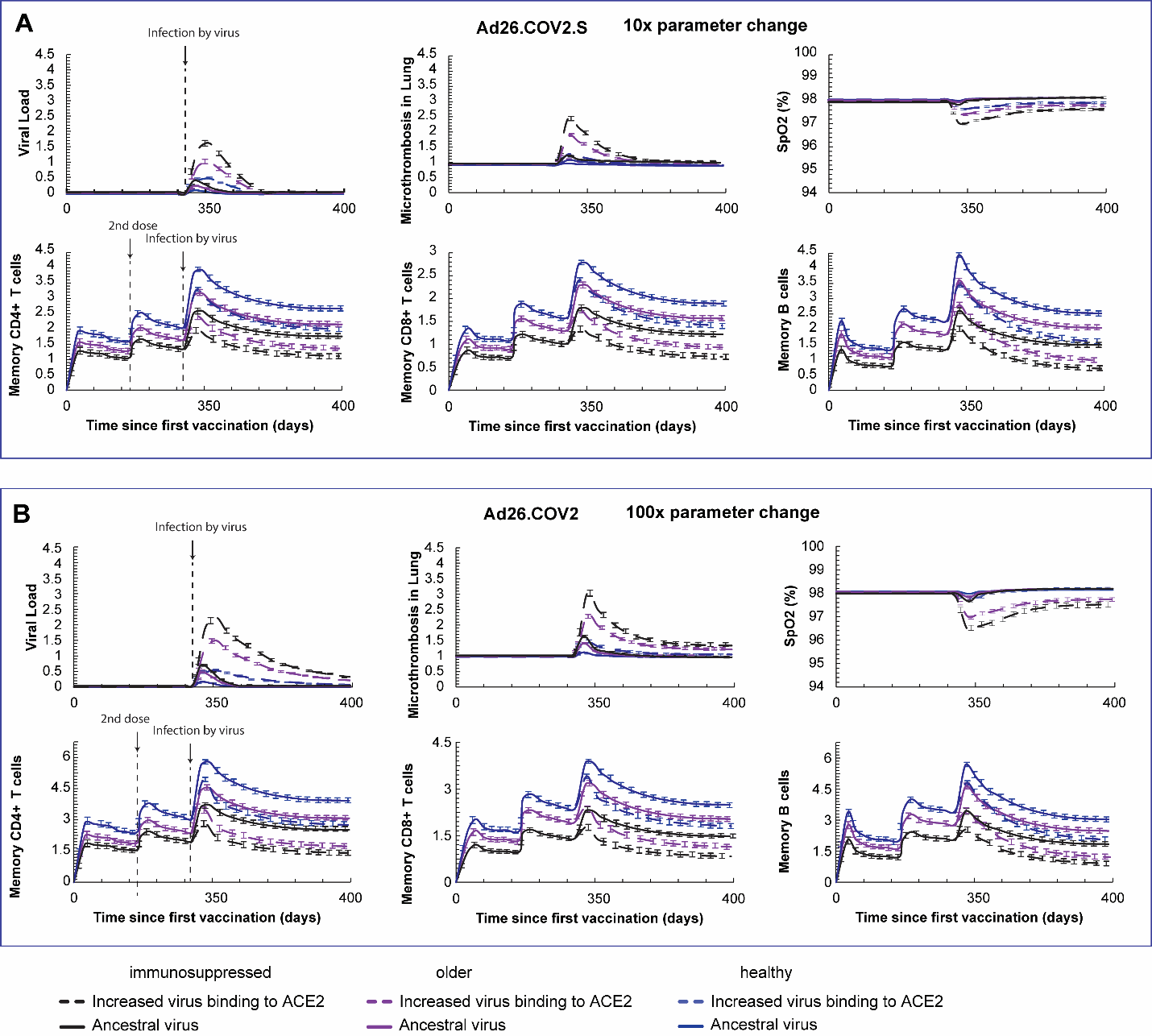

**Fig. S7**. **Vaccine (mRNA-1273) induced protection against severe disease caused by viral variants with mild (A) or severe (B) mutations in virus replication ability for healthy, older and immunosuppressed.** Predictions of the viral load, coagulation/microthrombi formation in the lungs, oxygen saturation (SpO2) and the concentration of the memory CD4^+^, CD8^+^ and B cells for breakthrough infections after BNT-162b2a vaccination and booster dose. Virus infection was assumed to take place 6 months after the booster dose. Normalized cell values are calculated by division with the initial value of the corresponding naïve cell type. The initial values of each naïve cell type are: immature dendritic cells 10^3^ [cells], Naïve CD4+ T cells 10^3^ [cells], Naïve CD8+ T cells 10^3^ [cells], Naïve B cells 10^3^ [cells]. The simulations were performed for mild mutations (10-fold change in parameter, A) and more severe mutations (100-fold change in parameter, B).

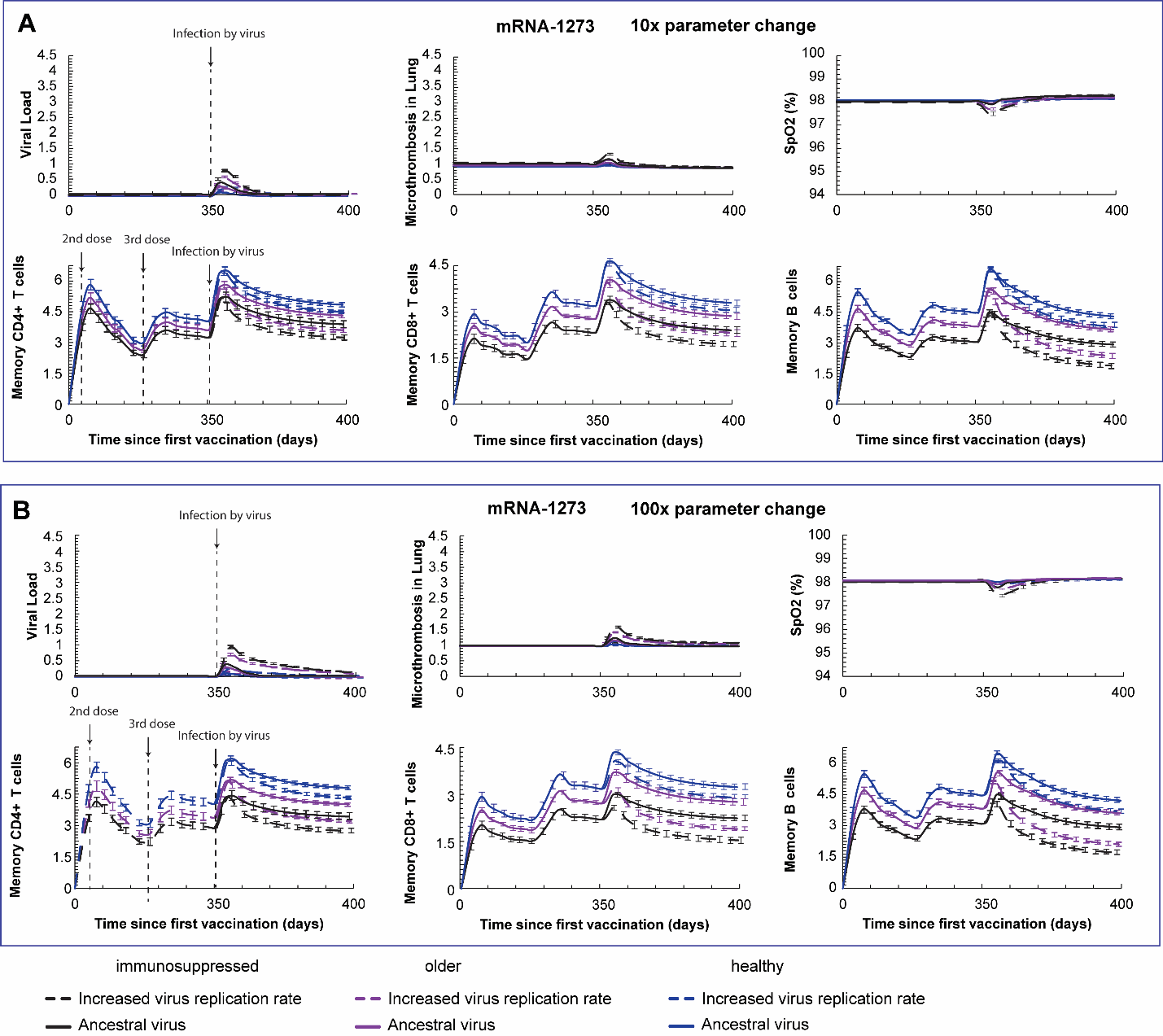

**Fig. S8**. **Vaccine (Ad26.COV2.S) induced protection against severe disease caused by viral variants with mild (A) or severe (B) mutations in virus replication ability for healthy, older and immunosuppressed.** Predictions of the viral load, coagulation/microthrombi formation in the lungs, oxygen saturation (SpO2) and the concentration of the memory CD4^+^, CD8^+^ and B cells for breakthrough infections after BNT-162b2a vaccination and booster dose. Virus infection was assumed to take place 6 months after the booster dose. Normalized cell values are calculated by division with the initial value of the corresponding naïve cell type. The initial values of each naïve cell type are: immature dendritic cells 10^3^ [cells], Naïve CD4+ T cells 10^3^ [cells], Naïve CD8+ T cells 10^3^ [cells], Naïve B cells 10^3^ [cells]. The simulations were performed for mild mutations (10-fold change in parameter, A) and more severe mutations (100-fold change in parameter, B).

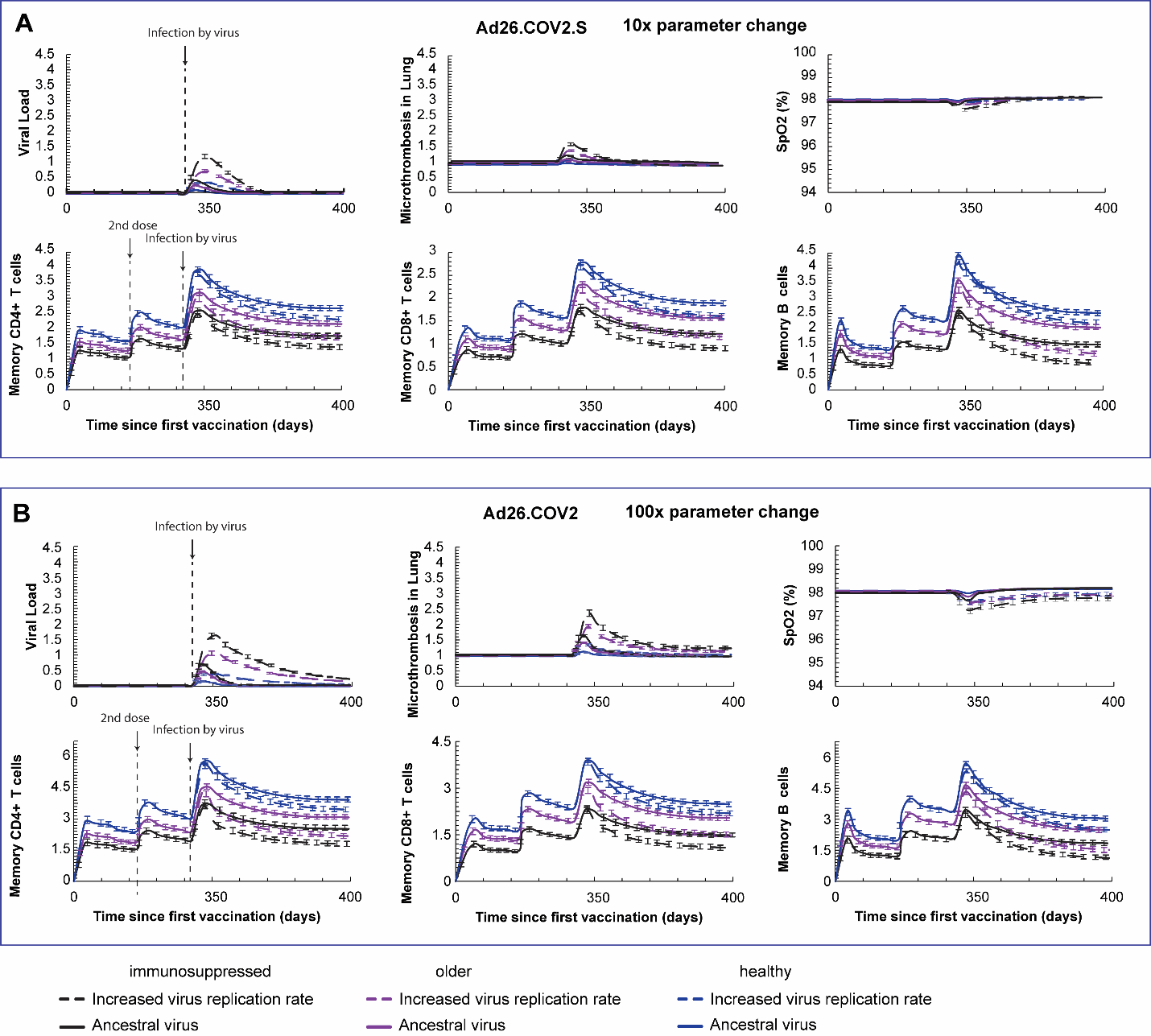

**Fig. S9**. Effect of timing of first booster on mRNA-1273 vaccine induced protection. Predictions of the viral load, coagulation/microthrombi formation in the lungs, oxygen saturation (SpO2) and the concentration of the memory CD4+, CD8+ and B cells for breakthrough infections after mRNA-1273 vaccination for intervals between the 2^nd^ and 3^rd^ doses of 3, 4, 5 and 6 months. Virus infection was assumed to take place 6 months after the booster dose. Normalized cell values are calculated by division with the initial value of the corresponding naïve cell type. Error bars represent the standard error for all values of model parameters considered in order to account for interpatient heterogeneity. The initial values of each naïve cell type are: immature dendritic cells 103 [cells], Naïve CD4+ T cells 103 [cells], Naïve CD8+ T cells 103 [cells], Naïve B cells 103 [cells].

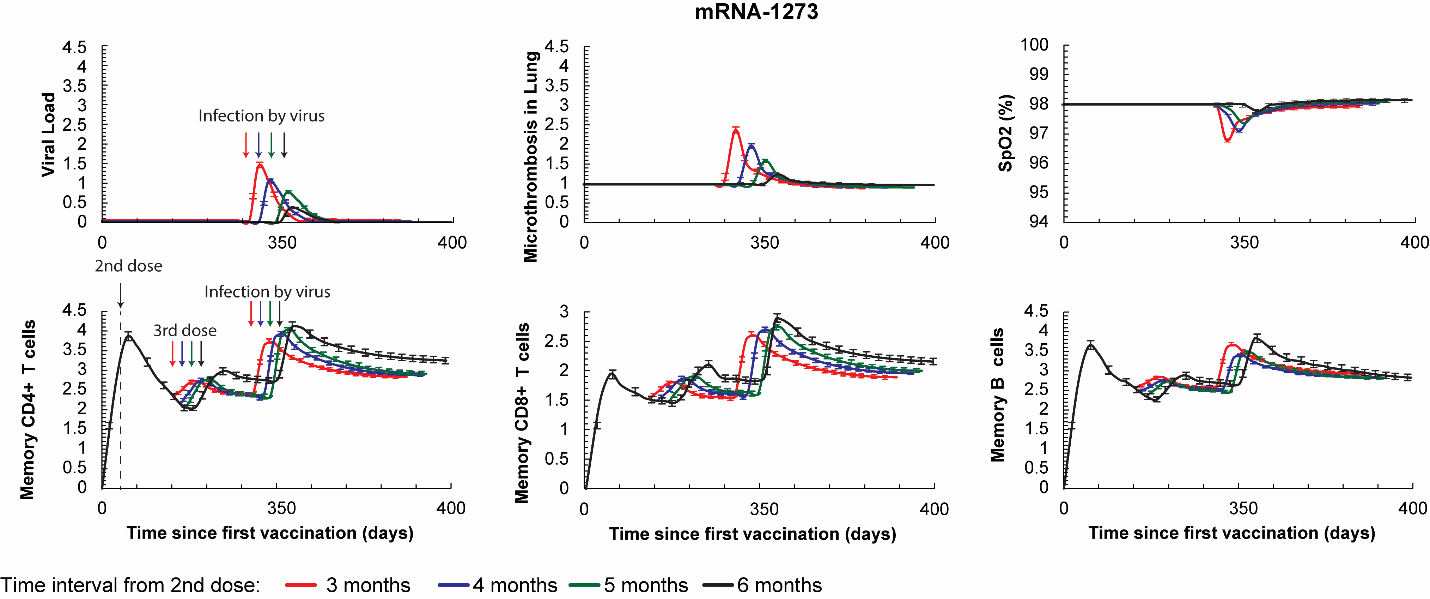

**Fig. S10**. Effect of timing of first booster dose on Ad26.COV2.S vaccine induced protection. Predictions of the viral load, coagulation/microthrombi formation in the lungs, oxygen saturation (SpO2) and the concentration of the memory CD4+, CD8+ and B cells for breakthrough infections after Ad26.COV2.S vaccination for intervals between the 1^st^ and 2^nd^ doses of 3, 4, 5 and 6 months. Virus infection was assumed to take place 6 months after the booster dose. Normalized cell values are calculated by division with the initial value of the corresponding naïve cell type. Error bars represent the standard error for all values of model parameters considered in order to account for interpatient heterogeneity. The initial values of each naïve cell type are: immature dendritic cells 10^3^ [cells], Naïve CD4+ T cells 10^3^ [cells], Naïve CD8+ T cells 10^3^ [cells], Naïve B cells 10^3^ [cells].

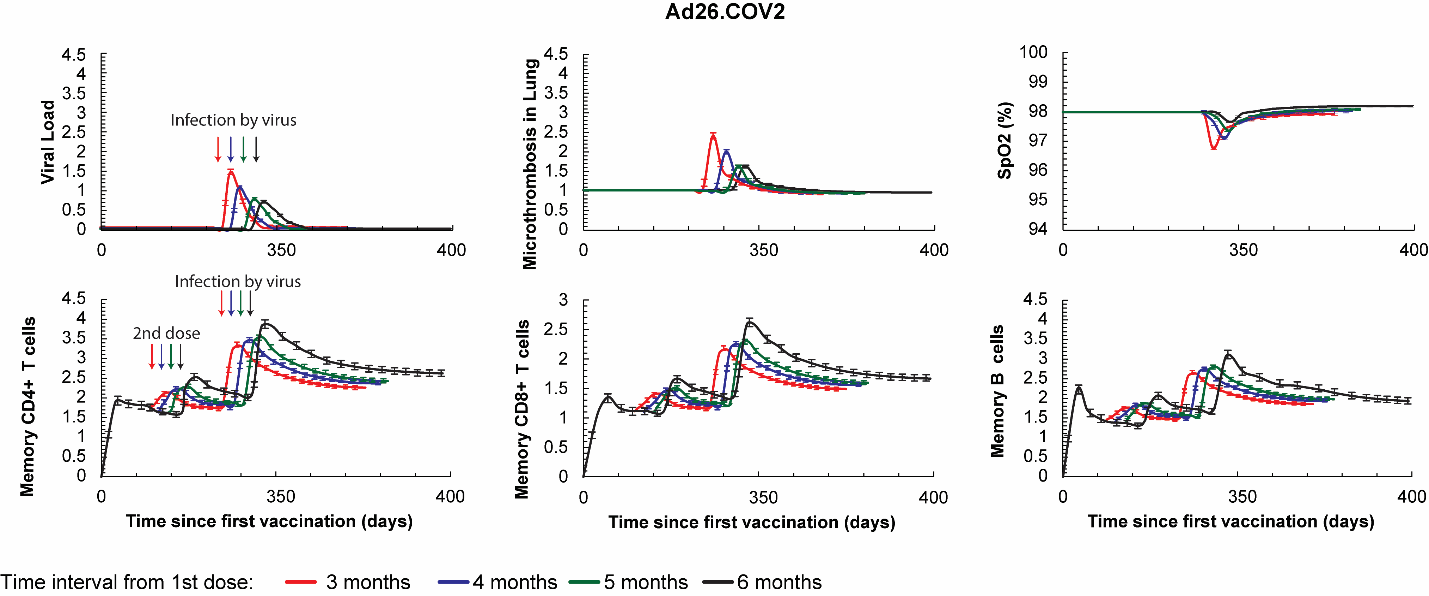

**Fig. S11**. Effect of timing of second booster on mRNA-1273 vaccine induced protection. Predictions of the viral load, coagulation/microthrombi formation in the lungs, oxygen saturation (SpO2) and the concentration of the memory CD4+, CD8+ and B cells for breakthrough infections after mRNA-1273 vaccination for intervals between the 3^rd^ and 4^th^ doses of 3, 4, 5 and 6 months. Virus infection was assumed to take place 6 months after the booster dose. Normalized cell values are calculated by division with the initial value of the corresponding naïve cell type. Error bars represent the standard error for all values of model parameters considered in order to account for interpatient heterogeneity. The initial values of each naïve cell type are: immature dendritic cells 10^3^ [cells], Naïve CD4+ T cells 10^3^ [cells], Naïve CD8+ T cells 10^3^ [cells], Naïve B cells 10^3^ [cells].

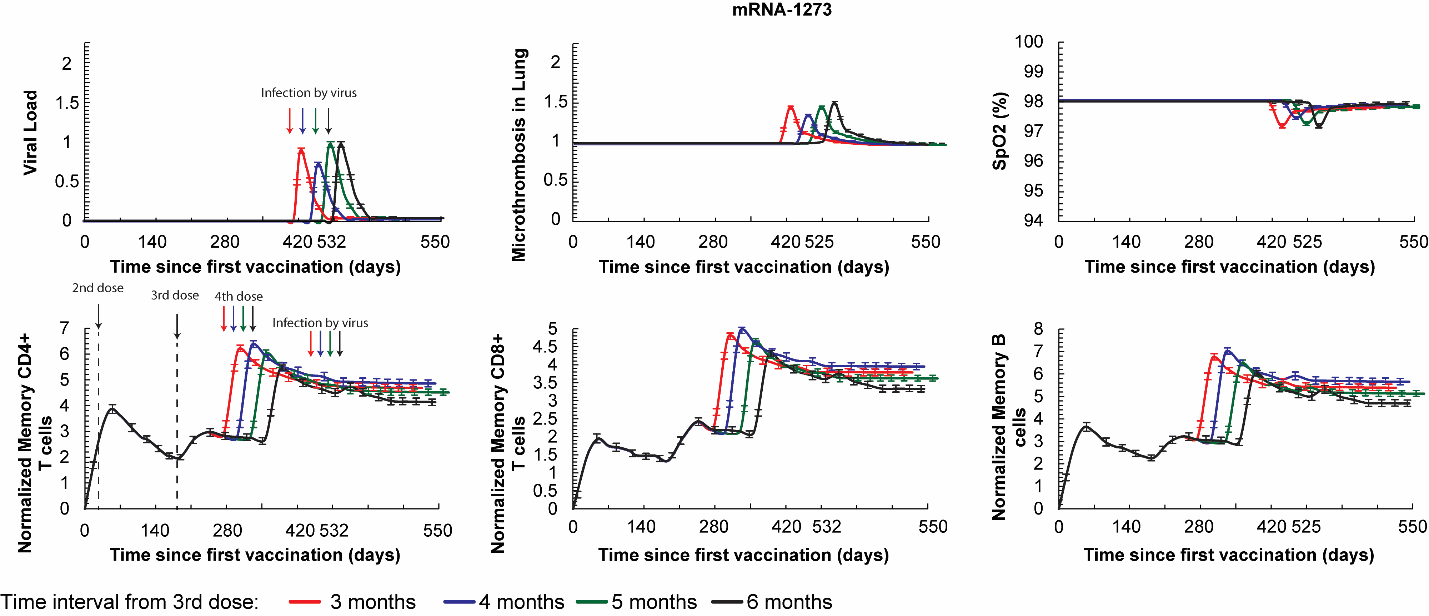

**Fig. S12**. Effect of timing of first booster on Ad26.COV2.S vaccine induced protection. Predictions of the viral load, coagulation/microthrombi formation in the lungs, oxygen saturation (SpO2) and the concentration of the memory CD4+, CD8+ and B cells for breakthrough infections after Ad26.COV2.S vaccination for intervals between the 2^nd^ and 3^rd^ doses of 3, 4, 5 and 6 months. Virus infection was assumed to take place 6 months after the booster dose. Normalized cell values are calculated by division with the initial value of the corresponding naïve cell type. Error bars represent the standard error for all values of model parameters considered in order to account for interpatient heterogeneity. The initial values of each naïve cell type are: immature dendritic cells 10^3^ [cells], Naïve CD4+ T cells 10^3^ [cells], Naïve CD8+ T cells 10^3^ [cells], Naïve B cells 10^3^ [cells].

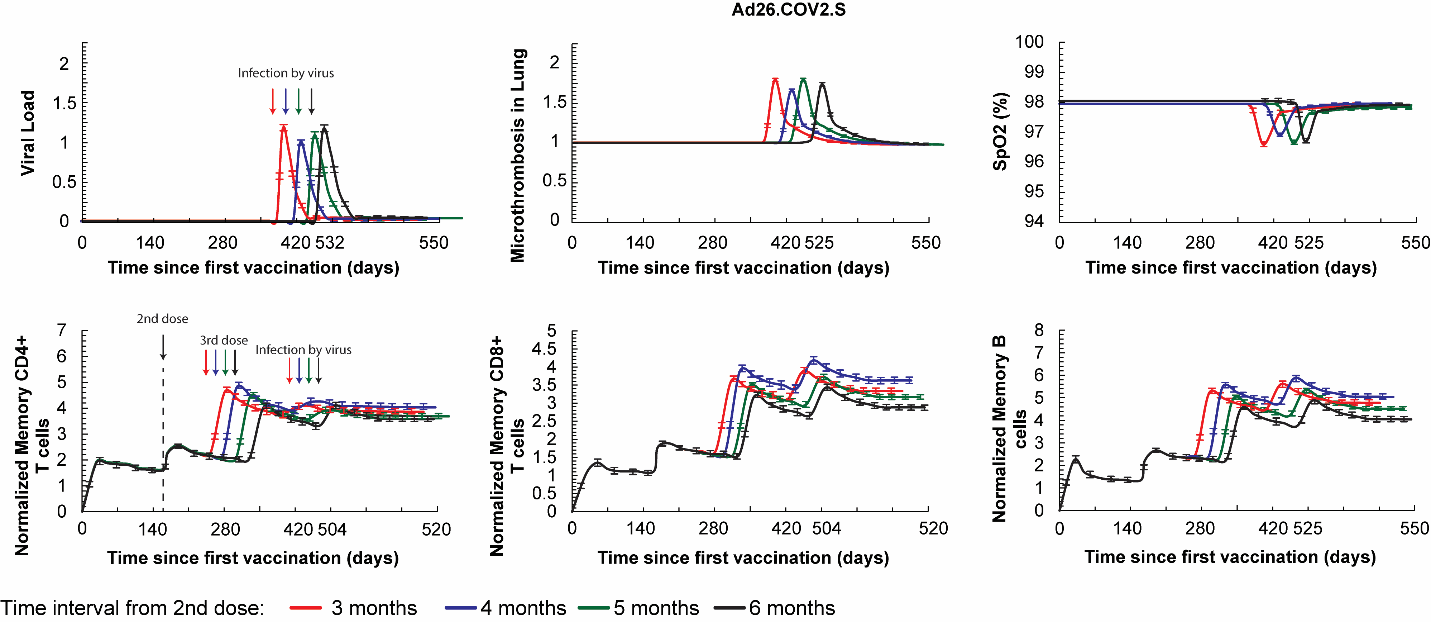

**Fig. S13.** Temporal variation of [PD1-PDL1] complex for varying the time to 1^st^ booster dose from 3 to 8 months for all vaccine types.

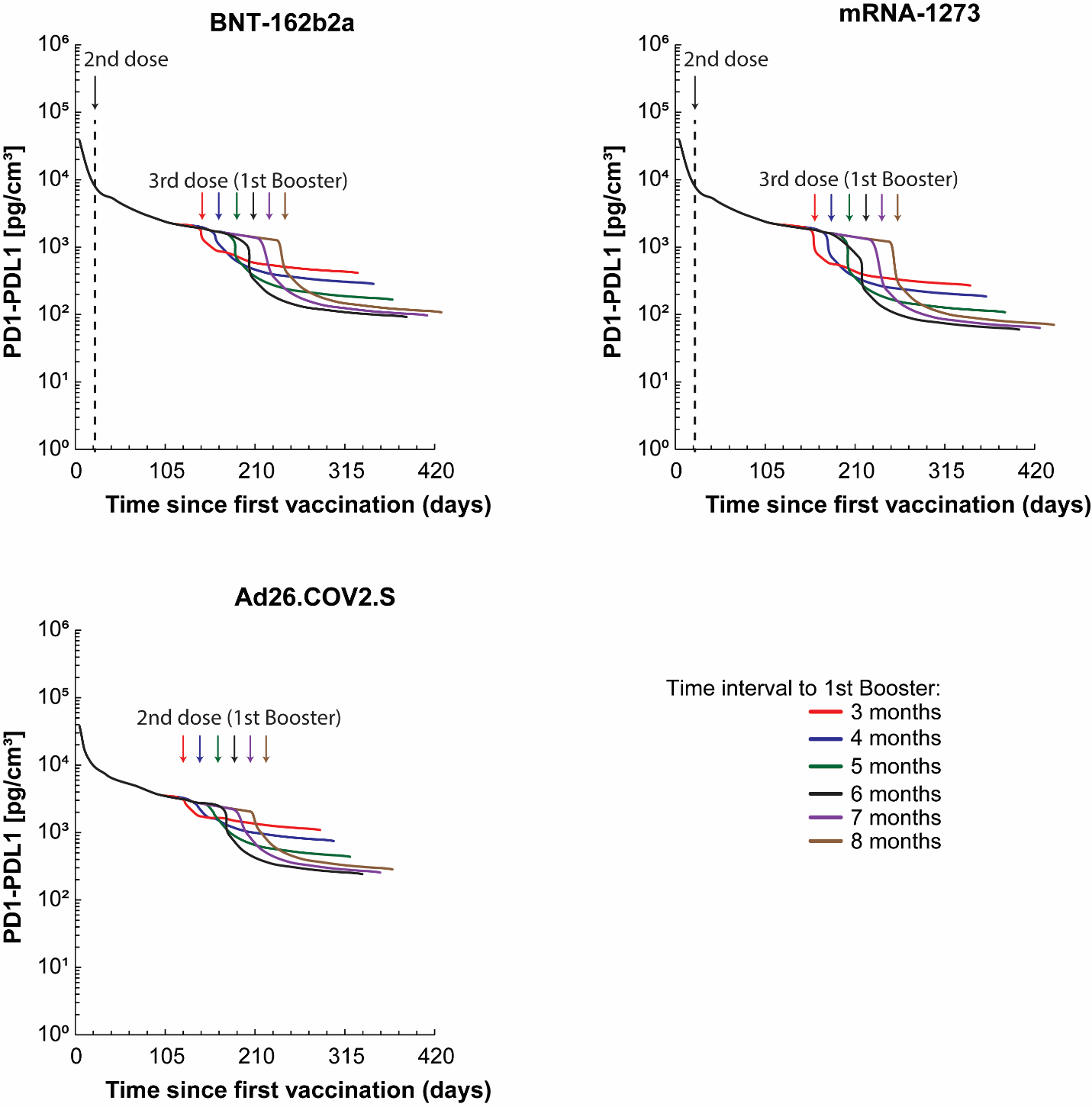

**Fig. S14.** Phase diagrams of memory CD4+, CD8+ and B cells caused by viral infection as a function of the time to 1st and 2nd booster vaccinations for all types of vaccines and for immunosuppressed individuals. Virus infection was assumed to take place 6 months following 2nd booster dose.

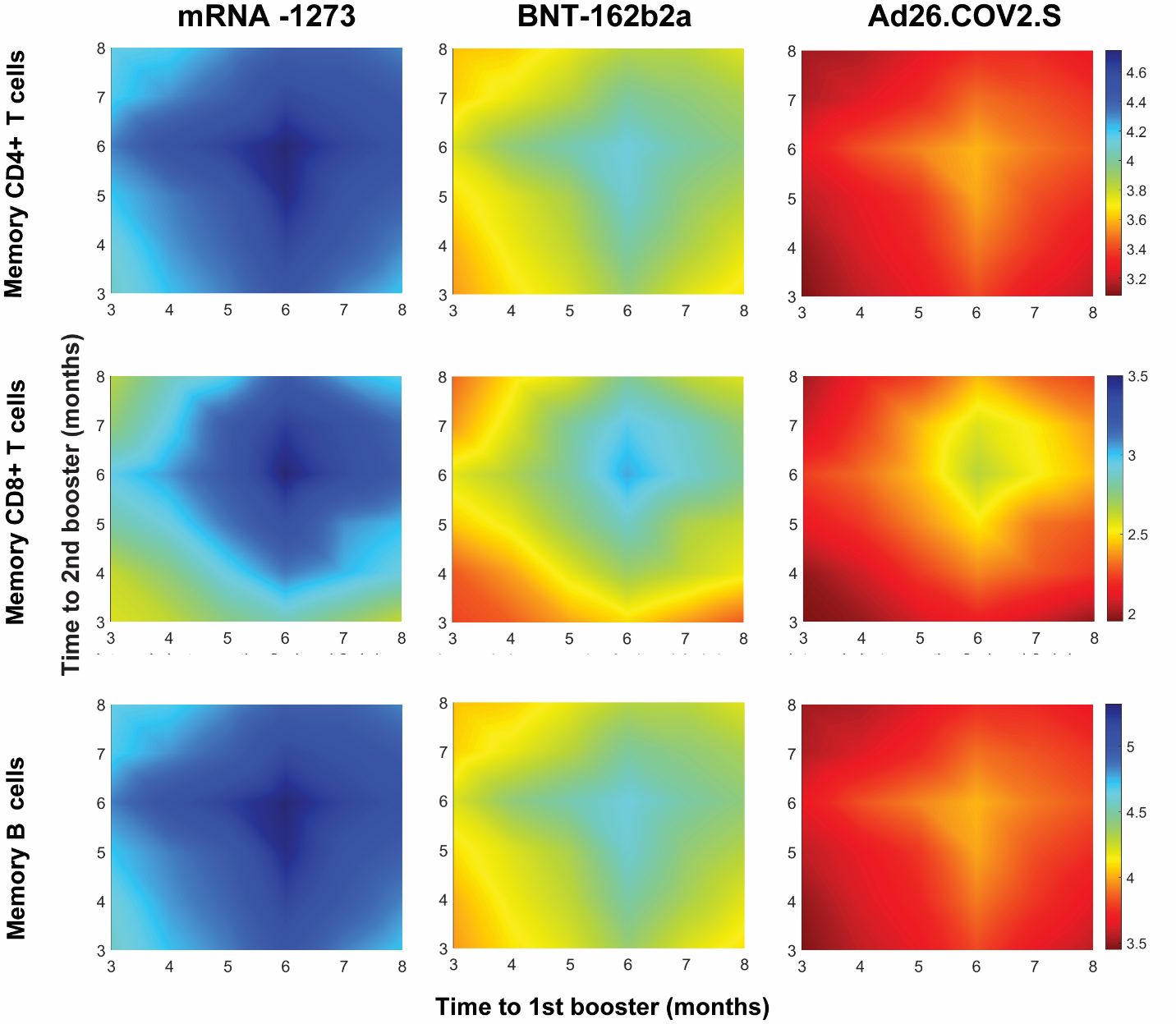

**Fig. S15.** Model predictions for viral infection of older people that have received a booster dose. For these simulations either the source term of the production of T cells (both CD4 and CD8) or the source term of the production of B cells was set to zero in order to investigate their relative contribution to protection.

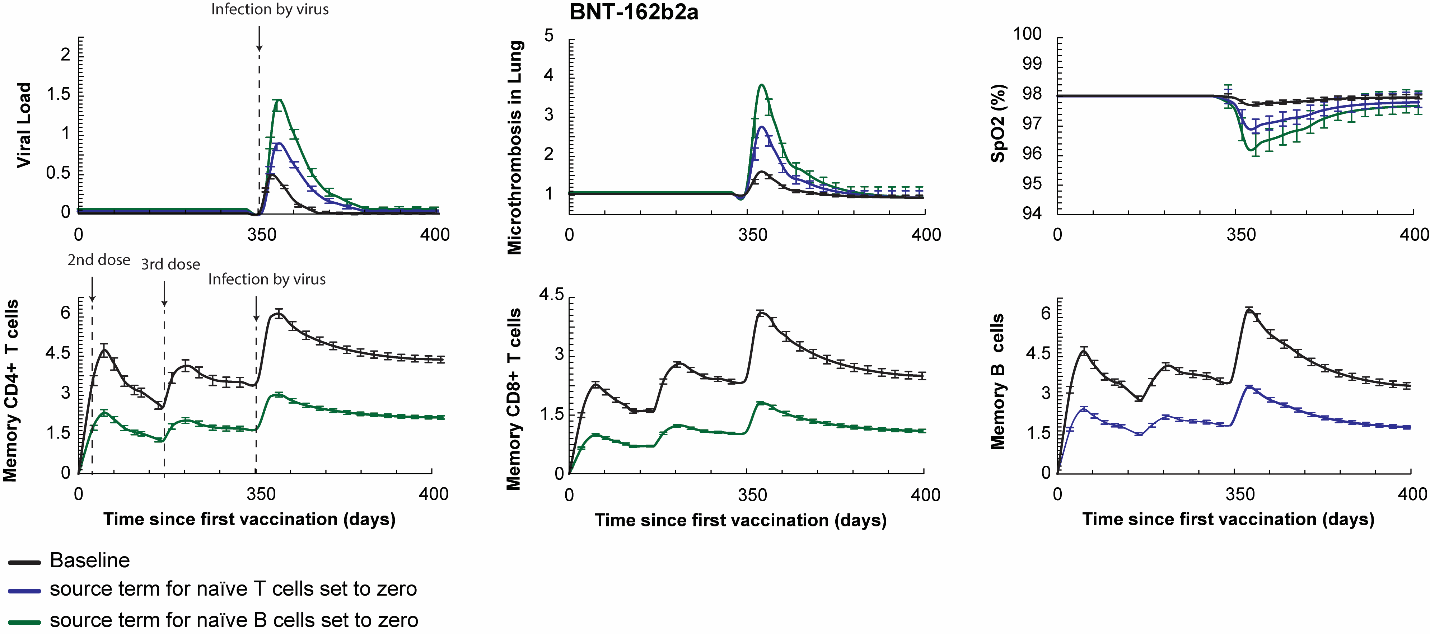

**Supplementary Table 1.** Values of model parameters related to vaccine activity and vaccination-induced immunity as well as changes in model parameters that account for immune-checkpoint inhibition, chemotherapy, B cell depletion and for the immunosuppressed patients (e.g., B cell depletion therapy, long term corticosteroids, TNF blockers). Baseline values were determined by validating the model to the clinical data and the range was taken to account for variability among individuals in the form of a sensitivity analysis. For the simulations, 100 equally spaced values for each parameter were considered to incorporate the variability among individuals and simulations were repeated for all possible combinations among parameter values. Results of model predictions for the baseline values are shown in the figures along with the results of individual variability as error bars.

| **Parameter** | **Description** | **BNT-162b2** | **mRNA-1273** | **Ad26.COV2.S** | **ICI** | **Chemotherapy** | **Immunosuppressed** |
| --- | --- | --- | --- | --- | --- | --- | --- |
| $\boldsymbol{K}_{\boldsymbol{cell}\mathbf{-}\boldsymbol{Vaccine}}^{\boldsymbol{on}}$ | Binding of the vaccine particles to healthy cells | Baseline:17[ml/h]  Range: 1.7-170 [ml/h] | Baseline: 17[ml/h] Range: 1.7-170 [ml/h] | Baseline: 15[ml/h] Range:1.5-150 [ml/h] |  |  |  |
| $\boldsymbol{K}_{\boldsymbol{cell}\mathbf{-}\boldsymbol{Vaccine}}^{\boldsymbol{off}}$ | unbinding of the vaccine particles to healthy cells | Baseline:6.22[m^3^/h]  Range: 0.622-62.2 [m^3^/h] | Baseline:5.22[m^3^/h]  Range: 0.522-52.2 [m^3^/h] | Baseline:5.22[m^3^/h]  Range: 0.522-52.2 [m^3^/h] |  |  |  |
| $\boldsymbol{K}_{\boldsymbol{dVa}}$ | Degradation rate of vaccine particles | Baseline:9.4e-4 [1/d]  Range: 9.4e-5 - 9.4e-3 [1/d] | Baseline:8.1e-4 [1/d]  Range: 8.1e-5-8.1e-3 [1/d] | Baseline: 9e-4 [1/d]  Range: 9e-5- 9 e-3 [1/d] |  |  |  |
| $\boldsymbol{K}_{\boldsymbol{intVa}}$ | internalization of the bound vaccine particles into the cells | Baseline: 4.78e-2[1/s]  Range: 4.78e-3 - 4.78e-1[1/s] | Baseline: 5.78e-2[1/s]  Range: 5.78e-3-5.78e-1[1/s] | Baseline: 5.78e-2[1/s]  Range: 5.78e-3-5.78e-1[1/s] |  |  |  |
| $\boldsymbol{K}_{\boldsymbol{dVab}}$ | degradation of the bound vaccine particles | Baseline: 4.5e-4 [1/d]  Range: 4.5e-5 - 4.5e-3 [1/d] | Baseline: 4e-4 [1/d]  Range: 4e-5 - 4e-3 [1/d] | Baseline: 5.5e-4 [1/d]  Range: 5.5e-5 - 5.5e-3 [1/d] |  |  |  |
| $\boldsymbol{K}_{\boldsymbol{Tran}}$ | DNA transcription to mRNA of the internalize vaccine particles | - | - | Baseline: 6.21e-4 [1/s]  Range: 6.21e-5 - 6.21e-3 [1/s] |  |  |  |
| $\boldsymbol{K}_{\boldsymbol{protein}}$ | production of viral protein or viral antigen from the translation of the mRNA | Baseline: 8.0e-5 [1/s]  Range: 8.0e-6 - 8.0e-4 [1/s] | Baseline: 8.52e-5 [1/s]  Range: 8.52e-6 - 8.52e-4 [1/s] | Baseline: 7.52e-5 [1/s]  Range: 7.52e-6 - 7.52e-4 [1/s] |  |  |  |
| $\boldsymbol{d}_{\boldsymbol{Vaint}}$ | constant degradation of internalized vaccine | Baseline: 3.8 e-4 [mole/m^3^/s]  Range: 3.8e-5 – 3.8e-3 [mole/m^3^/s] | Baseline: 3.4e-4 [mole/m^3^/s]  Range: 3.4e-5 - 3.4e-3 [mole/m^3^/s] | Baseline: 4.2e-4 [mole/m^3^/s]  Range: 4.2e-5 - 4.2e-3 [mole/m^3^/s] |  |  |  |
| $\boldsymbol{d}_{\boldsymbol{VaTran}}$ | constant degradation of translation DNA | Baseline: 2 e-4 [mole/m^3^/s]  Range: 2e-5 - 2e-3 [mole/m^3^/s] | Baseline: 1.5e-4 [mole/m^3^/s]  Range: 1.5e-5 - 1.5e-3 [mole/m^3^/s] | Baseline: 2.5e-4 [mole/m^3^/s]  Range: 2.5e-5 - 2.5e-3 [mole/m^3^/s] |  |  |  |
| $\boldsymbol{d}_{\boldsymbol{Vprotein}}$ | constant degradation of viral proteins | Baseline: 7e-5 [mole/m^3^/s]  Range: 7e-6 - 7e-4 [mole/m^3^/s] | Baseline: 6.5e-5 [mole/m^3^/s]  Range: 6.5e-6 - 6.5e-4 [mole/m^3^/s] | Baseline: 7.5e-5 [mole/m^3^/s]  Range: 7.5e-6 - 7.5e-4 [mole/m^3^/s] |  |  |  |
| $\boldsymbol{d}_{\boldsymbol{Q}}$ | Dissociation of PD-L1 from PD-1 |  |  |  | Baseline: 7.5e-5 [mole/m^3^/s]  Range: 1-100 [1/d] |  |  |
| **a_PL_** | Association of PD-1 with PD-L1 |  |  |  | Baseline: 7.5e-5 [mole/m^3^/s]  Range: 0.000258 -0.0258 [mm^3/g/s] |  |  |
| **g_A_** | Source of anti-PD1 |  |  |  | Baseline: 1x10^-8^ [g/cm^3/d]  Range: 1x10^-9^ - 1x10^-7^ [g/cm^3/d] |  |  |
| $\boldsymbol{d}_{\boldsymbol{i}\mathbf{-}\boldsymbol{Chemo}}$ | degradation of i cells from Chemotherapy |  |  |  |  | Baseline: 0.01 [1/d]  Range: 0.001 - 0.1 [1/d] |  |
| $\boldsymbol{S}_{\boldsymbol{TN}}\mathbf{,}\boldsymbol{S}_{\boldsymbol{ThN}}\mathbf{,}\boldsymbol{S}_{\boldsymbol{B}}$ | Source production of naïve CD8+, CD4+ and B cells |  |  |  |  |  | Baseline: 7.5 [1/d], 410^-4^ [1/d], 0.02 [1/d]  Range: 0.75 - 75 [1/d], 4x10-7 - 4x10-5 [1/d], 0.002 - 0.2 [1/d] |
| $\boldsymbol{h}_{\boldsymbol{TE}}$**,**$\boldsymbol{h}_{\boldsymbol{ThE}}\mathbf{,}\boldsymbol{h}_{\boldsymbol{BE}}$ | Conversion rate of Naïve CD8+, CD4+ and B cells to activated cells |  |  |  |  |  | Baseline: 0.00254 [1/h]  Range: 0.00000254 - 0.000254 [1/h] |

**Supplementary Table 2.** Values of model parameters related to mutations.

| **Parameter** | **Description** | **Baseline** | **Mild mutations** | **Severe mutations** |
| --- | --- | --- | --- | --- |
| $\boldsymbol{K}_{\boldsymbol{ACE}\boldsymbol{2-Virus}}^{\boldsymbol{on}}$ | virus binding to ACE2 | 17 [ml/h/nmol] | 170 [ml/h/nmol] | 1700 [ml/h/nmol] |
| $\boldsymbol{K}_{\boldsymbol{ACE}\boldsymbol{2-Virus}}^{\boldsymbol{off}}$ | virus detachment from ACE2 | 5.22[1/h] | 0.522[1/h] | 0.0522[1/h] |
| $\boldsymbol{K}_{\boldsymbol{a}}$ | virus exocytosis | 5.78x10^2^ [1/s] | 57.8x10^2^ [1/s] | 578x10^2^ [1/s] |
| $\boldsymbol{K}_{\boldsymbol{int}}$ | virus internalization | 5.78x10^-4^ [1/s] | 57.8x10^-4^ [1/s] | 578x10^-4^ [1/s] |
| $\boldsymbol{k}_{\boldsymbol{avp}}$ | virus replication rate | 0.5 | 5 | 50 |
| $\boldsymbol{k}_{\boldsymbol{v,A}}$ | antibody affinity to virus | 1x 10^-3^ [1/(mM.min)] | 1x 10^-4^ [1/(mM.min)] | 1x 10^-5^ [1/(mM.min)] |
| $\boldsymbol{\beta}_{\boldsymbol{DC}}$ | antigenicity | 1x10^-2^ [1/d/M] | 1x10^-3^ [1/d/M] | 1x10^-4^ [1/d/M] |

**Supplementary Table 3**: Values of Model parameters.

| Parameter | Description | Value [Units] | Reference |
| --- | --- | --- | --- |
| *K_AGT_* | Angiotensinogen production rate | 2.27x10^6^ [nmol/L/h] | Pilvankar et al. (1) |
| *c_Renin_* | Renin rate constant | 1.8x10^-14^ [1/s] | Pilvankar et al. (2) |
| *h_AGT_* | Angiotensinogen half-life | 10 [h] | Pilvankar et al. (1) |
| $\boldsymbol{h}_{\boldsymbol{Renin}}$ | Renin half-life | 0.25 [h] | Pilvankar et al. (1) |
| $\boldsymbol{K}_{\boldsymbol{f}}$ | Parameter for ANGII-renin feedback | 4.91x10^-5^ [1/h] | Pilvankar et al. (1) |
| *f* | Parameter for ANGII-renin feedback | 0.51 [nmol/ml] | Pilvankar et al. (1) |
| $\boldsymbol{K}_{\boldsymbol{Renin}}$ | Ang I production rate constant | 6.44x10^4^ [1/h] | Pilvankar et al. (1) |
| $\boldsymbol{K}_{\boldsymbol{ACE}}$ | Rate of conversion of ANGI->ANGII | 185.22[1/h] | Pilvankar et al. (1) |
| $\boldsymbol{K}_{\boldsymbol{NEP}}$ | Rate of conversion of ANGI->ANG(1-7) | 0.583 [1/h] | Pilvankar et al. (1) |
| $\boldsymbol{h}_{\boldsymbol{ANGI}}$ | Half-life of ANGI | 1.72x10^-4^ [h] | Pilvankar et al. (1) |
| $\boldsymbol{K}_{\boldsymbol{APA}}$ | Rate of conversion of ANGII->ANGIII | 43.6 [1/h] | Pilvankar et al. (1) |
| $\boldsymbol{h}_{\boldsymbol{ANGII}}$ | Half-life of ANGII | 5x10^-3^ [h] | Pilvankar et al. (1) |
| $\boldsymbol{h}_{\boldsymbol{AT}\boldsymbol{1}\boldsymbol{R-ANGII}}$ | Half-life of AT1R bound to ANGII | 1.5 [min] | Pilvankar et al. (2) |
| $\boldsymbol{h}_{\boldsymbol{AT}\boldsymbol{2}\boldsymbol{R-ANGII}}$ | Half-life of AT2R bound to ANGII | 1.5 [min] | Pilvankar et al. (2) |
| $\boldsymbol{h}_{\boldsymbol{ANG}_{\boldsymbol{(1-7)}}}$ | Half-life of ANG(1-7) | 30 [min] | Pilvankar et al. (2) |
| $\boldsymbol{h}_{\boldsymbol{MAsR-}\boldsymbol{ANG}_{\boldsymbol{(1-7)}}}$ | Half-life of Ang(1-7) bound to MAsR | 1.5 [min] | Estimate – same as *h _(AT1R-ANGII)_* |
| $\boldsymbol{h}_{\boldsymbol{ANG}_{\boldsymbol{(1-9)}}}$ | Half-life of ANG(1-9) | 24 [min] | Pilvankar et al. (2) |
| $\boldsymbol{K}_{\boldsymbol{APM}}$ | Rate of conversion of ANGIII->ANGIV | 43.6 [1/h] | Estimate – same as K_APA_ |
| $\boldsymbol{h}_{\boldsymbol{ANGIII}}$ | Half-life of ANGIII | 30 [s] | Pilvankar et al. (2) |
| $\boldsymbol{h}_{\boldsymbol{ANGIV}}$ | Half-life of ANGIV | 0.5 [min] | Lo A. et al. (3) |
| $\boldsymbol{h}_{\boldsymbol{AT}\boldsymbol{4}\boldsymbol{R-ANGIV}}$ | Half-life for ANGIV bound to AT4R | 1.5 [min] | Estimate – same as *h _(AT1R-ANGII)_* |
| $\boldsymbol{D}$ | Diffusion of the virus in the lung | 5x10^-3^ [cm^2^/s] | Estimate – Mok W. et al. (4) |
| $\boldsymbol{K}_{\boldsymbol{d}}$ | Inactivating rate of the virus | 4.8x10^-5^ [1/s] | Mok W. et al. (4) |
| $\boldsymbol{K}_{\boldsymbol{a}}$ | Rate of virus replication and release from the cell | 5.78x10^2^ [1/s] | Estimate - Mok W. et al. (4) |
| *K_IF_* | Strength of virus replication inhibition by Interferon | 0.0025[pg/ml] | Estimate |
| $\boldsymbol{K}_{\boldsymbol{int}}$ | Rate of bound virus internalization | 5.78x10^-4^ [1/s] | Mok W. et al. (4) |
| $\boldsymbol{K}_{\boldsymbol{AT}\boldsymbol{1}\boldsymbol{R}}$ | Rate of production of pro-inflammatory cytokines | 4.2 x10^2^ [pg/h/fmol] | Estimate |
| $\boldsymbol{S}_{\boldsymbol{n}}$ | Cytokine production by innate immune cells and infected cells | 2.1x10^-2^ [pg/h] | Smith A.M. et al (5) |
| $\boldsymbol{S}_{\boldsymbol{c}}$ | Cytokine production by the internalized virus | 2.9 x10^-2^ [pg/ml/h] | Smith A.M. et al (5) |
| $\boldsymbol{d}_{\boldsymbol{c}}$ | Degradation rate of pro-inflammatory cytokines | 8.3x10^-1^ [1/h] | Smith A.M. et al (5) |
| $\boldsymbol{K}_{\boldsymbol{g}}$ | Production of anti-inflammatory cytokines by macrophages and neutrophils | 2.1x10^-2^ [pg/h] | Dunster J.L. et al. (6) |
| $\boldsymbol{\varphi}_{\boldsymbol{a}}$ | Production of anti-inflammatory cytokines by macrophages interaction neutrophils | 2.1x10^-6^ [ml] | Dunster J.L. et al. (6) |
| $\boldsymbol{K}_{\boldsymbol{Ang}\boldsymbol{1-7}}$ | Production rate of anti-inflammatory cytokines by ANG(1-7) bound to MAs receptor | 4.2x10^2^ [pg/h/fmol] | Estimate |
| $\boldsymbol{\gamma}_{\boldsymbol{a}}$ | Degradation rate of anti-inflammatory cytokines | 3 [1/day] | Dunster J.L. et al. (6) |
| 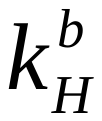 | Rate of conversion of healthy to infected epithelial cells | 1.36x10^5^ [1/M/s] | Su Z. and Wu Y. (7) |
| 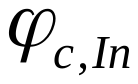 | Production of infected epithelial cells by neutrophils and pro/anti-inflammatory cytokines | 2.3x10^-9^ [ml/h] | Estimate |
| $\boldsymbol{R}_{\boldsymbol{H}}$ | Rate of production of healthy epithelial cells | 2.75x10^-3^ [1/h] | Mahasa KJ et al.(8) |
| $\boldsymbol{\chi}_{\boldsymbol{n}}$ | Production rate of neutrophils by pro-inflammatory cytokines | 2.1x10^-2^ [1/h] | Estimate- Dunster J.L. et al. (6) |
| $\boldsymbol{\gamma}_{\boldsymbol{n}}$ | Rate of production of neutrophil NETs | 6.3x10^-4^ [1/h] | Dunster J.L. et al. (6) |
| $\boldsymbol{\gamma}_{\boldsymbol{NETS}}$ | Degradation rate of NETs | 6.3x10^-7^[1/h] | Estimate |
| $\boldsymbol{\chi}_{\boldsymbol{m}}$ | Production rate of macrophages by pro-inflammatory cytokines | 0.02 [1/pg/h] | Estimate- Dunster J.L. et al. (6) |
| $\boldsymbol{\gamma}_{\boldsymbol{Ma}}$ | Degradation rate of macrophages | 6.3x10^-4^ [1/h] | Dunster J.L. et al. (6) |
| $\boldsymbol{K}_{\boldsymbol{ACE}\boldsymbol{2-ANGI}}^{\boldsymbol{on}}$ | Rate of ANGI binding to ACE2 receptors | 25 [ml/h/nmol] | Estimate – Mok W. et al. (4) |
| $\boldsymbol{K}_{\boldsymbol{ACE}\boldsymbol{2-ANGI}}^{\boldsymbol{off}}$ | Rate of ANGI detachment from ACE2 receptors | 5.22[1/h] | Estimate – Mok W. et al. (4) |
| $\boldsymbol{K}_{\boldsymbol{AT}\boldsymbol{1}}^{\boldsymbol{on}}$ | Rate of ANGII binding to AT1R receptors | 25 [ml/h/nmol] | Estimate – Mok W. et al. (4) |
| $\boldsymbol{K}_{\boldsymbol{AT}\boldsymbol{1}}^{\boldsymbol{off}}$ | Rate of ANGII detachment from AT1R receptors | 5.22[1/h] | Estimate – Mok W. et al. (4) |
| $\boldsymbol{K}_{\boldsymbol{AT}\boldsymbol{2}}^{\boldsymbol{on}}$ | Rate of ANGII binding to AT2R receptors | 25 [ml/h/nmol] | Estimate – Mok W. et al. (4) |
| $\boldsymbol{K}_{\boldsymbol{AT}\boldsymbol{2}}^{\boldsymbol{off}}$ | Rate of ANGII detachment from AT2R receptors | 5.22[1/h] | Estimate – Mok W. et al. (4) |
| $\boldsymbol{K}_{\boldsymbol{ACE}\boldsymbol{2-ANGII}}^{\boldsymbol{on}}$ | Rate of ANGII binding to ACE2 receptors | 25 [ml/h/nmol] | Estimate – Mok W. et al. (4) |
| $\boldsymbol{K}_{\boldsymbol{ACE}\boldsymbol{2-ANGII}}^{\boldsymbol{off}}$ | Rate of ANGII detachment from ACE2 receptors | 15.66[1/h] | Estimate – Mok W. et al. (4) |
| $\boldsymbol{K}_{\boldsymbol{ANG}\left( \boldsymbol{1-9} \right)}$ | Production of ANG(1-7) from ANG(1-9) | 5.22[1/h] | Estimate – Mok W. et al. (4) |
| $\boldsymbol{K}_{\boldsymbol{MAs}}^{\boldsymbol{on}}$ | Rate of ANG(1-7) binding to MAs receptors | 25 [ml/h/nmol] | Estimate – Mok W. et al. (9) |
| $\boldsymbol{K}_{\boldsymbol{MAs}}^{\boldsymbol{off}}$ | Rate of ANG(1-7) detachment from MAS receptors | 5.22[1/h] | Estimate – Mok W. et al. (4) |
| $\boldsymbol{K}_{\boldsymbol{AT}\boldsymbol{4}}^{\boldsymbol{on}}$ | Rate of ANGIV binding to AT4R receptors | 25 [ml/h/nmol] | Estimate – Mok W. et al. (4) |
| $\boldsymbol{K}_{\boldsymbol{AT}\boldsymbol{4}}^{\boldsymbol{off}}$ | Rate of ANGIV detachment from AT4R receptors | 5.22[1/h] | Estimate – Mok W. et al. (4) |
| $\boldsymbol{K}_{\boldsymbol{IL}\boldsymbol{6}}$ | Production rate of IL6 by activated macrophages | 0.5 [ml/h] | Estimate |
| $\boldsymbol{\gamma}_{\boldsymbol{IL}\boldsymbol{6}}$ | Degradation rate of IL6 | 6.3 x10^-4^ [1/h] | Estimate |
| $\boldsymbol{K}_{\boldsymbol{IL}\boldsymbol{6}}^{\boldsymbol{on}}$ | Rate of IL6 binding to IL6 receptors | 15 [ml/h/nmol] | Estimate – Mok W. et al. (4) |
| $\boldsymbol{K}_{\boldsymbol{IL}\boldsymbol{6}}^{\boldsymbol{off}}$ | Rate of IL6 detachment from IL6 receptors | 5.22[1/h] | Estimate – Mok W. et al. (4) |
| $\boldsymbol{h}_{\boldsymbol{IL}\boldsymbol{6}\boldsymbol{R}}$ | The half-life for IL6 receptor | 1.5 [min] | Estimate – same as $h_{AT1R-ANGII}$ |
| $\boldsymbol{S}_{\boldsymbol{sIL}\boldsymbol{6}\boldsymbol{R}}$ | Source production of sIL6R | 0.57[fmol/ml/h] | Estimate – same as $s_{Renin}$ |
| $\boldsymbol{K}_{\boldsymbol{sIL}\boldsymbol{6}\boldsymbol{R}}^{\boldsymbol{on}}$ | Rate of IL6 binding to soluble IL6 receptors | 17 [ml/h/nmol] | Estimate – Mok W. et al. (4) |
| $\boldsymbol{S}_{\boldsymbol{VEGF}}$ | Source production of VEGF | 0.235[fmol/ml/h] | Estimate – same as $\frac{s_{Renin}}{2}$ |
| $\boldsymbol{K}_{\boldsymbol{VEGF}}$ | Production rate of VEGF by IL6 bound on the soluble IL6 receptor | 5.22[1/h] | Estimate – Mok W. et al. (4) |
| $\boldsymbol{K}_{\boldsymbol{AT}\boldsymbol{1}\boldsymbol{R-MAsR}}$ | Degradation rate of pro-inflammatory cytokines by Mas receptor | 5.22[1/h] | Estimate – Mok W. et al. (4) |
| $\boldsymbol{K}_{\boldsymbol{AT}\boldsymbol{1}\boldsymbol{R-AT}\boldsymbol{2}\boldsymbol{R}}$ | Degradation rate of pro-inflammatory cytokines by AT2 receptor | 5.22[1/h] | Estimate – Mok W. et al. (4) |
| $\boldsymbol{S}_{\boldsymbol{AT}\boldsymbol{1}\boldsymbol{R}}$ | Source term for AT1R | 0.57[fmol/ml/h] | Estimate – same as $s_{Renin}$ |
| $\boldsymbol{S}_{\boldsymbol{AT}\boldsymbol{2}\boldsymbol{R}}$ | Source production of AT2R | 0.57[fmol/ml/h] | Estimate – same as $s_{Renin}$ |
| $\boldsymbol{S}_{\boldsymbol{MAsR}}$ | Source production of MAsR | 0.57[fmol/ml/h] | Estimate – same as $s_{Renin}$ |
| $\boldsymbol{S}_{\boldsymbol{AT}\boldsymbol{4}\boldsymbol{R}}$ | Source production of AT4R | 0.57[fmol/ml/h] | Estimate – same as $s_{Renin}$ |
| $\boldsymbol{S}_{\boldsymbol{IL}\boldsymbol{6}\boldsymbol{R}}$ | Source production of IL6R receptor | 0.57[fmol/ml/h] | Estimate – same as $s_{Renin}$ |
| $\boldsymbol{K}_{\boldsymbol{sIL}\boldsymbol{6}\boldsymbol{R}}$ | Conversion rate of IL6 receptor to soluble IL6 receptor | 5.22[1/h] | Estimate – Mok W. et al. (4) |
| $\boldsymbol{K}_{\boldsymbol{sIL}\boldsymbol{6}\boldsymbol{R}}^{\boldsymbol{off}}$ | Rate of IL6 detachment from soluble IL6 receptors | 2.22[1/h] | Estimate – Mok W. et al. (4) |
| $\boldsymbol{S}_{\boldsymbol{sACE}\boldsymbol{2}}$ | Source production of sACE2 | 0.57[fmol/ml/h] | Estimate – Mok W. et al. (4) |
| $\boldsymbol{K}_{\boldsymbol{sACE}\boldsymbol{2}}$ | Rate of soluble ACE2 receptor binding to the virus | 17 [ml/h/nmol] | Estimate – Mok W. et al. (4) |
| $\boldsymbol{K}_{\boldsymbol{Adam}\boldsymbol{17}}$ | Production rate of soluble ACE2 receptor by ACE2 receptor interaction through Adam17 | 5.22[1/h] | Estimate – Mok W. et al. (4) |
| $\boldsymbol{K}_{\boldsymbol{ACE}\boldsymbol{2-Virus}}^{\boldsymbol{on}}$ | Rate of virus binding to ACE2 receptors | 17 [ml/h/nmol] | Estimate – Mok W. et al. (4) |
| $\boldsymbol{K}_{\boldsymbol{ACE}\boldsymbol{2-Virus}}^{\boldsymbol{off}}$ | Rate of virus detachment from ACE2 receptors | 5.22[1/h] | Estimate – Mok W. et al. (4) |
| $\boldsymbol{\chi}_{\boldsymbol{N-IL}\boldsymbol{6}}$ | Production of neutrophils by IL6-R activation | 5.26[1/fmol/h] | Estimate- Dunster J.L. et al. (6) |
| $\boldsymbol{\chi}_{\boldsymbol{Ma-IL}\boldsymbol{6}}$ | Production of macrophages by IL6-R activation | 5.26[1/fmol/h] | Estimate- Dunster J.L. et al. (6) |
| $\boldsymbol{K}_{\boldsymbol{ec}}$ | Proliferation of endothelial cells | 5.22x10^-7^ [1/h] | Estimate |
| $\boldsymbol{K}_{\boldsymbol{ec}}^{\boldsymbol{b}}$ | Endothelial cell infection rate | 70 [ml/h/nmol] | Estimate |
| $\boldsymbol{S}_{\boldsymbol{v}}^{\boldsymbol{0}}$ | Vascular density of the normal lung | 70 [1/cm] | Mpekris F. et al (10) |
| $\boldsymbol{K}_{\boldsymbol{ACE}\boldsymbol{2}}$ | Production of ANG(1-7) by ANGII bound to ACE2 | 65.22[1/h] | Pilvankar et al. (1) |
| $\boldsymbol{S}_{\boldsymbol{ACE}\boldsymbol{2}}$ | Production rate of ACE2 by healthy endothelial and epithelial cells | 0.57[fmol/ml/h] | Estimate – same as $s_{Renin}$ |
| $\boldsymbol{V}_{\boldsymbol{O}\boldsymbol{2}}^{\boldsymbol{max}}$ | oxygen uptake rate | 3700 [mlO_2_/min] | Weibel E.R. et al (11) |
| $\boldsymbol{P}_{\boldsymbol{A}}$ | Partial pressure of oxygen in alveolar air | 100 [mmHg] | T.K. Roy, T.W. Secomb (12) |
| $\boldsymbol{K}_{\boldsymbol{02}}^{\boldsymbol{0}}$ | Krogh permeability coefficient KO2 | 3.3x10^-8^ [cm^2^/min/mmHg] | Weibel E.R. et al (11) |
| $\boldsymbol{S(A)}$ | Alveolar gas exchange areas | 130 [m^2^] | Weibel E.R. et al (11) |
| $\boldsymbol{S(c)}$ | Capillary gas exchange areas | 115 [m^2^] | Weibel E.R. et al (11) |
| $\boldsymbol{\tau}_{\boldsymbol{hb}}$ | Harmonic mean thickness of the air–blood barrier | 1 [μm] | Weibel E.R. et al (11) |
| $\boldsymbol{\theta}_{\boldsymbol{O}\boldsymbol{2}}$ | Oxygen unloading conductance of blood | 1.8[mlO_2_/ml/min/mmHg] | Roy T.K., Secomb T.W. (12) |
| $\boldsymbol{V(c)}$ | Lung blood volume | 194 [ml] | Roy T.K., Secomb T.W. (12) |
| $\boldsymbol{n}$ | Hill coefficient | 2.7 | Roy T.K., Secomb T.W. (12) |
| $\boldsymbol{P}_{\boldsymbol{50}}$ | Oxygen tension when the binding sites are 50 percent saturated. | 26.3 [mmHg] | Roy T.K., Secomb T.W. (12) |
| $\boldsymbol{K}_{\boldsymbol{in}}$ | Rate of release of replicated virus | 0.4 x10^-7^ [1/h] | Mahasa KJ et al.(8) |
| $\boldsymbol{d}_{\boldsymbol{AT}\boldsymbol{1}\boldsymbol{R}}$ | Degradation rate of AT1R | 6.3x10^-4^[1/h] | Estimate- Dunster J.L. et al. (6) |
| $\boldsymbol{d}_{\boldsymbol{AT}\boldsymbol{2}\boldsymbol{R}}$ | Degradation rate of AT2R | 6.3x10^-4^[1/h] | Estimate- Dunster J.L. et al. (6) |
| $\boldsymbol{d}_{\boldsymbol{MAsR}}$ | Degradation rate of MAsR | 6.3x10^-4^[1/h] | Estimate- Dunster J.L. et al. (6) |
| $\boldsymbol{d}_{\boldsymbol{AT}\boldsymbol{4}\boldsymbol{R}}$ | Degradation rate of AT4R | 6.3x10^-4^[1/h] | Estimate- Dunster J.L. et al. (6) |
| $\boldsymbol{d}_{\boldsymbol{IL}\boldsymbol{6}\boldsymbol{R}}$ | Degradation rate of IL6R | 6.3x10^-4^[1/h] | Estimate- Dunster J.L. et al. (6) |
| $\boldsymbol{d}_{\boldsymbol{sACE}\boldsymbol{2}}$ | Degradation rate of soluble ACE2 receptor | 6.3x10^-4^[1/h] | Estimate- Dunster J.L. et al. (6) |
| $\boldsymbol{K}_{\boldsymbol{IL}\boldsymbol{6-TN}}$ | Production rate of IL6 by Naïve T cells | 0.02554[mol/pg/s] | Estimate - Lai, X., & Friedman A. (13) |
| $\boldsymbol{K}_{\boldsymbol{IL}\boldsymbol{6-In}}$ | Production rate of IL6 by infected epithelial cells | 0.02554[mol/pg/s] | Estimate - Lai, X., & Friedman A. (13) |
| $\boldsymbol{K}_{\boldsymbol{IL}\boldsymbol{6-TE}}$ | Production rate of IL6 by Activated T cells | 0.02554[mol/pg/s] | Estimate - Lai, X., & Friedman A. (13) |
| $\boldsymbol{K}_{\boldsymbol{IL}\boldsymbol{6-iEC}}$ | Production rate of IL6 by infected endothelial cells | 0.02554[mol/pg/s] | Estimate - Lai, X., & Friedman A. (13) |
| $\boldsymbol{\gamma}_{\boldsymbol{VEGF}}$ | Production rate of VEGF by hypoxia | 0.0152[mol/ml/h] | Estimate |
| $\boldsymbol{K}_{\boldsymbol{AT}\boldsymbol{1}\boldsymbol{R}}$ | Production of pro-inflammatory cytokines by AT1R activation | 4.2x10^2^ [pg/h/fmol] | Estimate |
| $\boldsymbol{K}_{\boldsymbol{c-IL}\boldsymbol{6}}$ | Production of pro-inflammatory cytokines by IL6-R activation | 0.03 [ml/h/fmol] | Estimate |
| $\boldsymbol{K}_{\boldsymbol{c-EC}}$ | Production of pro-inflammatory cytokines by healthy endothelial cells | 1 [pg] | Estimate |
| $\boldsymbol{K}_{\boldsymbol{c-H}}$ | Production of pro-inflammatory cytokines by healthy epithelial cells | 1 [pg] | Estimate |
| $\boldsymbol{K}_{\boldsymbol{HIn}}\boldsymbol{=}\boldsymbol{K}_{\boldsymbol{HIEC}}$ | proportion coefficients for production of infected epithelial and endothelial cells by neutrophils and pro/anti-inflammatory cytokines | 0.02554 [ml/pg] | Estimate |
| $\boldsymbol{h}_{\boldsymbol{T}}\boldsymbol{=}\boldsymbol{h}_{\boldsymbol{TE}}$=$\boldsymbol{h}_{\boldsymbol{ThE}}\mathbf{=}\boldsymbol{h}_{\boldsymbol{BE}}$ | Conversion rate of Naïve T and B cells to activated cells | 0.00254 [1/h] | Estimate - Lai, X., & Friedman A. (13) |
| $\boldsymbol{As}$ | Antigen strength | 1 | Estimate - Lai, X., & Friedman A. (13) |
| $\boldsymbol{K}_{\boldsymbol{T}}$ | Constant for blocking PD-1 inhibition | 1.365x10^-18^ [g/cm^3^] | Estimate - Lai, X., & Friedman A. (13) |
| $\boldsymbol{\varepsilon}$ | Degradation of activated T cells by PD1 bound to PDL1 | 0.01575 [cm^3^/g/s] | Estimate - Lai, X., & Friedman A. (13) |
| $\boldsymbol{d}_{\boldsymbol{H}}$ | Degradation rate PDL1 by healthy epithelial cells | 0.00215 [1/s] | Estimate - Lai, X., & Friedman A. (13) |
| $\boldsymbol{d}_{\boldsymbol{EC}}$ | Degradation rate PDL1 by endothelial healthy cells | 0.00215 [1/s] | Estimate - Lai, X., & Friedman A. (13) |
| $\boldsymbol{d}_{\boldsymbol{In}}$ | Degradation rate PDL1 by epithelial infected Cells | 0.00215 [1/s] | Estimate - Lai, X., & Friedman A. (13) |
| $\boldsymbol{d}_{\boldsymbol{TE}}$ | Degradation rate PD1 by Activated T cells | 0.00215x10^-3^ [1/s] | Estimate - Lai, X., & Friedman A. (13) |
| $\boldsymbol{d}_{\boldsymbol{TN}}$ | Degradation rate PD1 by Naïve T cells | 0.00215 [1/s] | Estimate - Lai, X., & Friedman A. (13) |
| $\boldsymbol{d}_{\boldsymbol{N}}$ | Degradation rate PD1 by neutrophils | 0.00215 [1/s] | Estimate - Lai, X., & Friedman A. (13) |
| $\boldsymbol{d}_{\boldsymbol{Ma}}$ | Degradation rate PD1 by macrophages | 0.00215 [1/s] | Estimate - Lai, X., & Friedman A. (13) |
| a_PL_ | Association of PD-1 with PD-L1 | 0.258[mm^3/g/s] | Estimate - Lai, X., & Friedman A. (13) |
| $\boldsymbol{d}_{\boldsymbol{Q}}$ | Dissociation rate of PD-L1 from PD-1 | 0.1 [1/d] | Estimate - Lai, X., & Friedman A. (13) |
| γ_A_ | Source of anti-PD1 | 1x10^-10^[g/cm^3/d] | Estimate - Lai, X., & Friedman A. (13) |
| µ_PD1_ | Efficiency of PD1 blocking by ICI | 0.00215x10^-3^[1/s] | Estimate - Lai, X., & Friedman A. (13) |
| $\boldsymbol{d}_{\boldsymbol{A}}$ | Degradation rate of anti-PD1 | 0.0462 [1/d] | Estimate - Lai, X., & Friedman A. (13) |
| $\boldsymbol{\lambda}_{\boldsymbol{H}}$ | Production rate of PDL1 by healthy epithelial cells | 0.154 [1/s] | Estimate - Lai, X., & Friedman A. (13) |
| $\boldsymbol{\lambda}_{\boldsymbol{EC}}$ | Production rate of PDL1 by healthy endothelial cells | 0.154 [1/s] | Estimate - Lai, X., & Friedman A. (13) |
| $\boldsymbol{\lambda}_{\boldsymbol{In}}$ | Production rate of PDL1 by infected endothelial cells | 0.154 [1/s] | Estimate - Lai, X., & Friedman A. (13) |
| $\boldsymbol{\lambda}_{\boldsymbol{TE}}$ | Production rate of PD1 by activated T cells | 0.154 [1/s] | Estimate - Lai, X., & Friedman A. (13) |
| $\boldsymbol{\lambda}_{\boldsymbol{TN}}$ | Production rate of PD1 by Naïve T cells | 0.154 [1/s] | Estimate - Lai, X., & Friedman A. (13) |
| $\boldsymbol{\lambda}_{\boldsymbol{N}}$ | Production rate of PD1 by neutrophils | 0.154 [1/s] | Estimate - Lai, X., & Friedman A. (13) |
| $\boldsymbol{\lambda}_{\boldsymbol{Ma}}$ | Production rate of PD1 by macrophages | 0.154 [1/s] | Estimate - Lai, X., & Friedman A. (13) |
| $\boldsymbol{h}_{\boldsymbol{PDL}\boldsymbol{1}}$ | Production rate of the PD1 ligand by Effector (Activated) T | 0.154 [1/s] | Estimate - Lai, X., & Friedman A. (13) |
| $\boldsymbol{\mu}_{\boldsymbol{PD}\boldsymbol{1}}$ | Degradation rate of PD1 by anti-PD1 | 0.00215 x10^-3^ [1/s] | Estimate - Lai, X., & Friedman A. (13) |
| $\boldsymbol{a}_{\boldsymbol{PL}}$ | rate of PD1 binding to PDL1 | 0.258 [mm^3^/g/s] | Estimate - Lai, X., & Friedman A. (13) |
| $\boldsymbol{\gamma}_{\boldsymbol{A}}$ | Source term of anti-PD1 | 1x10^-10^ [g/cm^3^/d] | Estimate - Lai, X., & Friedman A. (13) |
| $\boldsymbol{\mu}_{\boldsymbol{A}}$ | Degradation rate of anti-PD1 by PD1 | 6.87x10^6^ [cm^3^/g/d] | Estimate - Lai, X., & Friedman A. (13) |
| 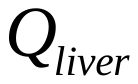 | Blood flow rate of liver (hepatic portal vein from G.I. and spleen, and hepatic artery) | 800 [ml/min] | (14) |
| 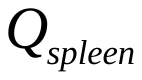 | Blood flow rate of spleen | 138 [ml/min] | (14) |
| 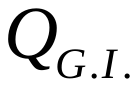 | Blood flow rate of G.I. | 468 [ml/min] | (14) |
| 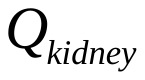 | Blood flow rate of kidney | 630 [ml/min] | (14) |
| 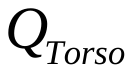 | Blood flow rate of Torso | 220 [ml/min] | Estimate - (14) |
| 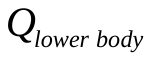 | Blood flow rate of lower body | 413 [ml/min] | Estimate - (14) |
| 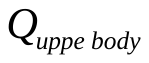 | Blood flow rate of upper body | 138 [ml/min] | Estimate - (14) |
| 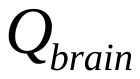 | Blood flow rate of brain | 300 [ml/min] | Estimate - (14) |
| 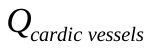 | Blood flow rate of cardiac vessels | 120 [ml/min] | Estimate - (14) |
| 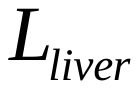 | Lymphatic flow rate of liver | 8.7x10^-2^ [ml/min] | (14) |
| 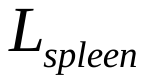 | Lymphatic flow rate of spleen | 8.7x10^-4^ [ml/min] | (14) |
| 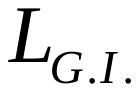 | Lymphatic flow rate of G.I. | 3.0 x 10^-1^ [ml/min] | (14) |
| 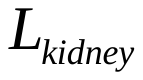 | Lymphatic flow rate of kidney | 1x 10^-3^ [ml/min] | Estimate - (14) |
| 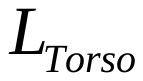 | Lymphatic flow rate of torso | 4.3x 10^-3^ [ml/min] | Estimate - (14) |
| 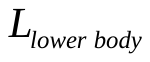 | Lymphatic flow rate of lower body | 1x 10^-3^ [ml/min] | Estimate - (14) |
| 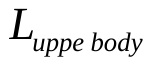 | Lymphatic flow rate of upper body | 2.6 x 10^-2^ [ml/min] | Estimate - (14) |
| 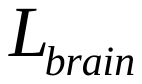 | Lymphatic flow rate of brain | 1x 10^-3^ [ml/min] | Estimate - (14) |
| 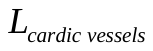 | Lymphatic flow rate of cardiac vessels | 4.3x 10^-3^ [ml/min] | Estimate - (14) |
| 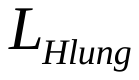 | Lymphatic flow rate of normal part of lung | 4.3x 10^-2^ [ml/min] | (14) |
| 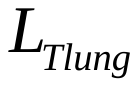 | Lymphatic flow rate of tumor part of lung | 2x 10^-2^ [ml/min] | Estimate - (14) |
| 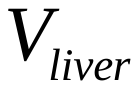 | Averaged vascular volume of liver | 180.9 [ml] | (14) |
| 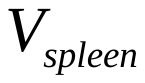 | Averaged vascular volume of spleen | 17 [ml] | (14) |
| 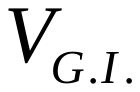 | Averaged vascular volume of GI system | 43 [ml] | (14) |
| 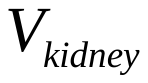 | Averaged vascular volume of kidney | 28.4 [ml] | (14) |
| 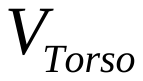 | Averaged vascular volume of torso | 462 [ml] | Estimate |
| 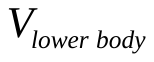 | Averaged vascular volume of lower body | 700 [ml] | Estimate |
| 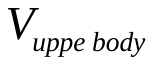 | Averaged vascular volume of upper body | 150 [ml] | Estimate |
| 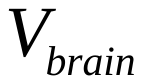 | Averaged vascular volume of brain | 150 [ml] | Estimate |
| 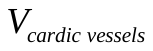 | Averaged vascular volume of cardiac vessels | 100 [ml] | Estimate |
| 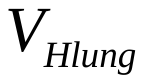 | Averaged vascular volume of normal part of lung | 99.9 [ml] | (14) |
| 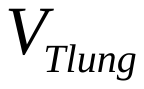 | Averaged vascular volume of normal part of lung | 50 [ml] | Estimate |
| 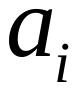 | Attachment rate of virus to ACE2 | 1x 10^-3^ [1/(mM.min)] | Estimate |
| 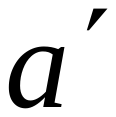 | Attachment rate of virus to sACE2 | 8.5 [ml/h/nmol] | Estimate |
| 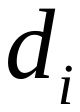 | Detachment rate of virus from endothelium | 6.7x 10^-4^ [1/min] | Estimate |
| 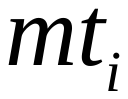 | Maximum production of micro-thrombus (ACE2) | 7x 10^-3^ [mM/min] | Estimate |
| 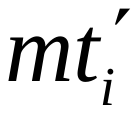 | Maximum production of micro-thrombus (IL6) | 7x 10^-3^ [mM/min] | Estimate |
| *mt_i_’’* | Maximum production of micro-thrombus (cytokines) | 7x 10^-3^ [mM/min] | Estimate |
| 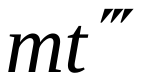 | Maximum production of micro-thrombus (NET) | 7x 10^-3^ [mM/min] | Estimate |
| 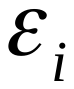 | Dissolution of micro-thrombi due to thrombolysis | 7x 10^-5^ [1/min] | Estimate |
| 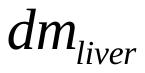 | Attachment rate of micro-thrombus to vessels in liver | 6.9x 10^-6^ [1/min] | Estimate |
| 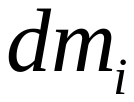 | Attachment rate of micro-thrombus to vessels in other organs | 6.9x 10^-6^ [1/min] | Estimate |
| 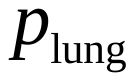 | Proliferation rate of internalized virus in lung | 7x 10^-3^ [1/min] | Estimate |
| 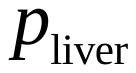 | Proliferation rate of internalized virus in liver | 7x 10^-3^ [1/min] | Estimate |
| 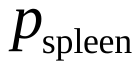 | Proliferation rate of internalized virus in spleen | 0.7x 10^-3^ [1/min] | Estimate |
| 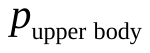 | Proliferation rate of internalized virus in upper body | 7.6x 10^-3^ [1/min] | Estimate |
| 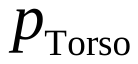 | Proliferation rate of internalized virus in torso | 14x 10^-3^ [1/min] | Estimate |
| 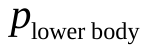 | Proliferation rate of internalized virus in lower body | 6.3x 10^-3^ [1/min] | Estimate |
| 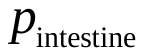 | Proliferation rate of internalized virus in intestine | 4.9x 10^-3^ [1/min] | Estimate |
| 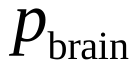 | Proliferation rate of internalized virus in brain | 9.7x 10^-3^ [1/min] | Estimate |
| 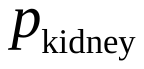 | Proliferation rate of internalized virus in kidney | 7.6x 10^-3^ [1/min] | Estimate |
| 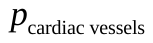 | Proliferation rate of internalized virus in cardiac vessels | 12.5x 10^-3^ [1/min] | Estimate |
| 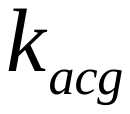 | Micro-thrombus-inhibiting coefficient of anti-coagulation drugs for virus-ACE-2-induced coagulation | 0.5 | Estimate |
| 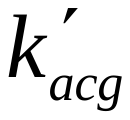 | Micro-thrombus-inhibiting coefficient of anti-coagulation drugs for IL6-induced coagulation | 0.5 | Estimate |
| 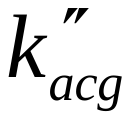 | Micro-thrombus-inhibiting coefficient of anti-coagulation drugs for cytokine storm-induced coagulation | 0.5 | Estimate |
| 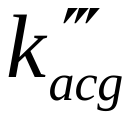 | Micro-thrombus-inhibiting coefficient of anti-coagulation drugs for NET-induced coagulation | 0.5 | Estimate |
| 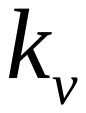 | Degradation rate of free virus by Antibody | 4x10^-6^ [1/d/nmol] | Estimate – Lee et al (15) |
| 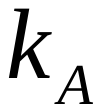 | Clearance rate of antibody by free viruses | 0.04 [1/d/nmol] | Lee et al (15) |
| 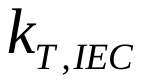 | Production rate of the micro-thrombus by infected endothelial cells | 1.4x 10^-3^ [mM/min] | Estimate |
| 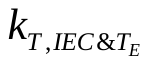 | Production rate of the micro-thrombus by infected endothelial cells and effector CD8+ T cells | 1.4x 10^-3^ [mM/min] | Estimate |
| 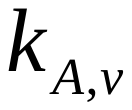 | Clearance rate of antibody by free viruses | 1x 10^-3^ [1/(mM.min)] | Estimate |
| 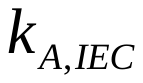 | Degradation rate of antibody by infected endothelial cells | 2.1x10^-10^ [ml/h/nmol] | Estimate |
| 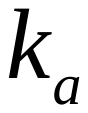 | Natural decay rate of antibody | 4x10^-7^ [1/d ] | Estimate |
| 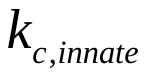 | Production rate of the cytokine by innate immune cells | 2.3x10^-9^ [ml/h/nmol] | Estimate |
| 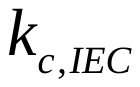 | Production rate of the cytokine by infected endothelial cells | 2.3x10^-9^ [ml/h/nmol] | Estimate |
| 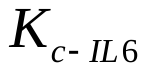 | Production rate of the cytokine by IL6 bound to sIL6R | 0.03 [ml/h/fmol] | Estimate |
| 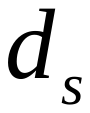 | Degradation rate of pro-inflammatory cytokines | 8.3x10^-1^ [1/h] | Smith A.M. et al (5) |
| 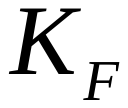 | Killing rate of infected cells by NK cells | 4.2 [1/d] | A. Haghnegahdar et all (16) |
| 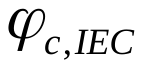 | proportion coefficient for production of infected endothelial cells by neutrophils and pro/anti-inflammatory cytokines | 2.1x10^-2^ [pg/h] | Dunster J.L. et al. (6) |
| 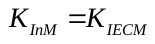 | Killing rate of infected cells by Memory cells | 4.2 [1/d] | Estimate |
| 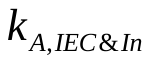 | Degradation rate of antibody by infected endothelial and epithelial cells | 2.1x10^-10^ [ml/h] | Estimate |
| 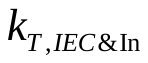 | Production rate of the micro-thrombus by infected endothelial and epithelial cells | 1.4x 10^-3^ [mM/min] | Estimate |
| 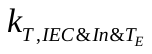 | Production rate of the micro-thrombus by infected endothelial and epithelial cells and effector CD8+ T cells | 1.4x 10^-3^ [mM/min] | Estimate |
| 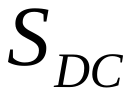 | source term of dendritic cells | 1 [1/d] | Lee et al (15) |
| 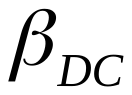 | Conversion rate of immature dendritic cells to APC | 1x10^-2^ [1/d/M] | Lee et al (15) |
| 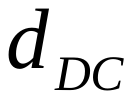 | Death rate of dendritic cells | 1x10^-3^ [1/d] | Lee et al (15) |
| 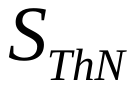 | source term of Naïve CD4+ T cells | 4x10^-4^ [1/d] | Lee et al (15) |
| 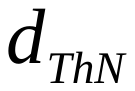 | Death rate of Naïve CD4+ T cells | 0.75 [1/d] | Lee et al (15) |
| $\boldsymbol{\pi}_{\boldsymbol{ThM}}$ | conversion rate of effector CD4+ T cells to memory CD4+ T cells | - | Estimate |
| 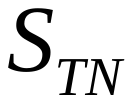 | source term of Naïve CD8+ T cells | 750 [1/d] | Lee et al (15) |
| 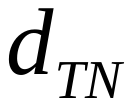 | Death rate of Naïve CD8+ T cells | 0.75 [1/d] | Lee et al (15) |
| $\boldsymbol{\pi}_{\boldsymbol{TM}}$ | conversion rate of effector CD8+ T cells to memory CD8+ T cells | - | Estimate |
| 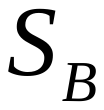 | source term of Naïve B cells | 2 [1/d] | Lee et al (15) |
| 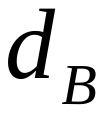 | Death rate of Naïve B cells | 0.002 [1/d] | Lee et al (15) |
| $\boldsymbol{\pi}_{\boldsymbol{s}}$ | differentiation rate of activated B cells into short-lived antibody-secreting plasma cells. | 1e-3 [1/d] | Lee et al (15) |
| $\boldsymbol{\pi}_{\boldsymbol{L}}$ | differentiation rate of activated B cells into long-lived antibody-secreting plasma cells. | 8e-9 [1/d] | Lee et al (15) |
| $\boldsymbol{\rho}_{\boldsymbol{B}}$ | the proliferation rate of activated B cells | 2.6 [1/d] | Lee et al (15) |
| $\boldsymbol{\delta}_{\boldsymbol{L}}$ | death rate of long-lived antibody-secreting plasma cell | 3e-2 [1/d] | Lee et al (15) |
| $\boldsymbol{\delta}_{\boldsymbol{s}}$ | death rate of short-lived antibody-secreting plasma cell | 0.1 [1/d] | Lee et al (15) |
| $\boldsymbol{\delta}_{\boldsymbol{ThM}}$ | death rate of memory CD4+ cells | 3e-4 [1/d] | Estimate |
| $\boldsymbol{\delta}_{\boldsymbol{TM}}$ | death rate of memory CD8+ cells | 3e-4 [1/d] | Estimate |
| $\boldsymbol{S}^{\boldsymbol{Va}}$ | Source term for Free vaccine | 1[M/d] | Estimate |
| $\boldsymbol{K}_{\boldsymbol{cell}\mathbf{-}\boldsymbol{Vaccine}}^{\boldsymbol{on}}$ | Binding of the vaccine particles to Healthy cells | 17 [ml/h] | Estimate |
| $\boldsymbol{K}_{\boldsymbol{cell}\mathbf{-}\boldsymbol{Vaccine}}^{\boldsymbol{off}}$ | unbinding of the vaccine particles to Healthy cells | 5.22[m^3^/h] | Estimate |
| $\boldsymbol{K}_{\boldsymbol{dVa}}$ | Degradation rate of vaccine particles | 8.1e-4 [1/d] | Estimate |
| $\boldsymbol{K}_{\boldsymbol{intVa}}$ | internalization of the bound vaccine particles into the cells | 5.78e-2[1/s] | Estimate |
| $\boldsymbol{K}_{\boldsymbol{dVab}}$ | degradation of the bound vaccine particles | 4e-4 [1/d] | Estimate |
| $\boldsymbol{K}_{\boldsymbol{Tran}}$ | DNA transcription to mRNA of the internalize vaccine particles | 6.21 e-4 [1/s] | Estimate |
| $\boldsymbol{K}_{\boldsymbol{protein}}$ | production of viral protein or viral antigen from the translation of the mRNA | 8.52 e-5 [1/s] | Estimate |
| $\boldsymbol{d}_{\boldsymbol{Vaint}}$ | constant degradation of internalized vaccine | 3.4 e-4 [mole/m^3^/s] | Estimate |
| $\boldsymbol{d}_{\boldsymbol{VaTran}}$ | constant degradation of translation DNA | 1.5 e-4 [mole/m^3^/s] | Estimate |
| $\boldsymbol{d}_{\boldsymbol{Vprotein}}$ | constant degradation of viral proteins | 6.5 e-5 [mole/m^3^/s] | Estimate |

**Description of mathematical model**

We developed a mechanistic model of COVID-19 immunity and infection based on previous research (17) and used this to explore how variation in viral, host and vaccine characteristics may affect COVID-19 outcomes. The model incorporates the infection of lung epithelium by SARS-CoV-2, the response of innate and adaptive immune cells to infection, the production of pro-and anti-inflammatory cytokines, the activation of the coagulation cascade, as well as the proliferation of cancer cells. The model further accounts for the interactions between the virus, the immune cells and the tumor cells as well as for vaccination-induced immunity (**Fig 1**). The basic components of the model are:

**Virus infection**: SARS-CoV-2 enters the cell by docking to ACE2, a key component of the Renin-angiotensin system (RAS). ACE2 can be membrane-bound or soluble, and it regulates inflammation by converting Ang II to Ang 1-7 and Ang I to Ang 1-9 (17). Intracellular virus initiates inflammatory pathways through toll-like receptors and NFκB, which produces interferons and other inflammatory cytokines. The viral antigens, along with inflammatory cytokines, facilitate activation of naïve T cells, creating virus-specific T effector cells. T cell activation is controlled by viral antigen strength and the status of immune checkpoint inhibition (specifically PD-L1 / PD-1). In the presence of inflammatory cytokines and virus, neutrophils can produce neutrophil extracellular traps (NETs).

**Vaccination-induced immunity.** The model considers separately the mechanisms of mRNA and vector vaccines. The vaccines, as particles, either lipid nanoparticles in the case of mRNA vaccines or viral-vector in the case of vector vaccines, enter host cells and either induce DNA transcription to mRNA (vector vaccine) and then translation into viral antigen or result directly in translation of viral antigens (mRNA vaccine). Subsequently, vaccine-induced peptides exit the cells and interact with dendritic cells to produce antigen presenting cells. These subsequently activate T cells and B cells to create CD4^+^ and CD8^+^ effector and memory T cells as well as short-lived and long-lived plasma (antibody-secreting) B cells.

**Coagulation cascade**. The virus can infect endothelial cells and disseminate via the blood stream, with the possibility of systemic infection and thrombosis. Infection of endothelial cells, combined with high levels of inflammatory cytokines in the plasma, can result in thrombosis. Damage to virally infected endothelial cells and the production of NETs can exacerbate the thrombosis, and microthrombi can enter the blood stream to accumulate in other organs, including the brain, heart and lung. We use a simplified model of the coagulation pathways, assuming that formation of microthrombi is proportional to the number of infected endothelial cells, the presence of neutrophil NETs, and the level of inflammatory cytokines. Transport of oxygen from the alveolar space to the blood vessels in the lung is calculated using a modified diffusion model, which accounts for damage-induced thickening of the alveolar membrane.

**Pharmacokinetic-pharmacodynamic (PK/PD) model.** The PK/PD model has been formulated to incorporate major organs: lung, heart, liver, brain, spleen, gastro-intestinal, upper body, lower body, torso, cardiac vessels. The tumor is also incorporated in the PK/PD model as a separate organ. The PK/PD model allows for transport of viral particles, antibodies, cytokines and micro-thrombi among these compartments (Supplementary Fig 1).

**Tumor progression.** Cancer cell proliferation depends on the oxygen levels in the tissue, and their death rate depends on the interaction of cancer cells with immune cells (effector CD8^+^ T cells, natural killer cells, type 1 macrophages and neutrophils) as well as on the effect of cancer therapy (18, 19). We assume that cancer cells are not infected by the virus.

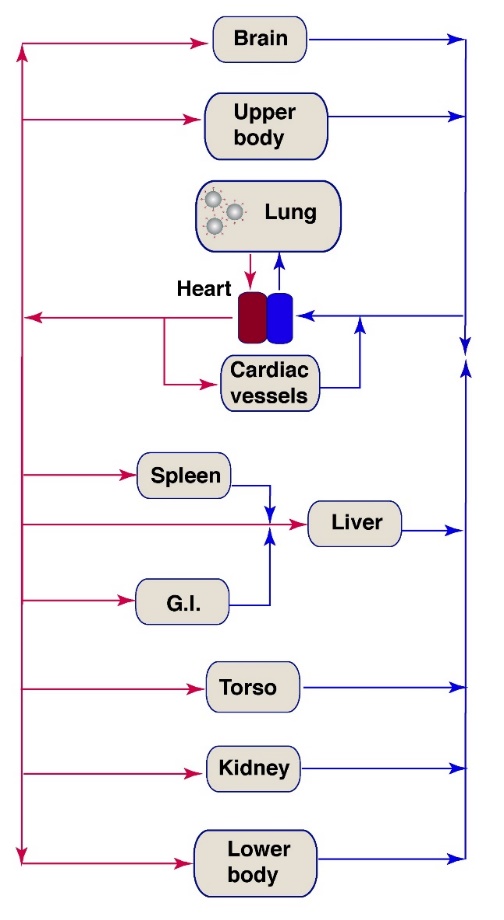

**Fig S1**. **PK/PD model of COVID-19 infection and thrombosis simulates events that happen throughout the body when a patient contracts COVID-19.** Virus exits the lung via the systemic circulation, and can infect endothelial cells (ECs) in various organs, modeled here as well-mixed compartments. Upon infection, viral replication and cell death cause vessel damage and local inflammation that induce thrombosis. Each tissue can then become a source of micro-thrombi, which enter the systemic circulation and can accumulate in the microvessels of the heart, lung and brain, inducing ischemic events.

**Equations of the Mathematical Modeling Framework**

1. Pharmacokinetic-pharmacodynamic (PK/PD) model

The PK/PD model has been formulated to incorporate major organs: lung, heart, liver, brain, spleen, gastro-intestinal, upper body, lower body, torso, cardiac vessels and the tumor . The PK/PD model allows for transport of viral particles, antibodies, cytokines and micro-thrombi among these compartments.

1.1 Heart compartment

In this model, heart circulates blood through all the compartments. The heart compartment includes arterial and venous blood flows for free viruses, micro-thrombosis, cytokines and antibody. aHeart and vHeart refer to the left and right heart ventricles, respectively.

1.1.1 Free virus recirculation in arterial and venous blood flows into left and right heart ventricles,
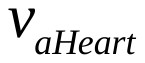
and
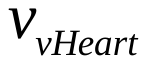
,

| 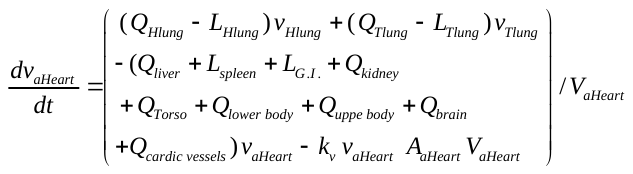 | (1a) |
| --- | --- |

where the first group of terms represents the recirculation in arterial blood flow free viruses and the last term describes the effect of the antibodies on the virus reproduction

| 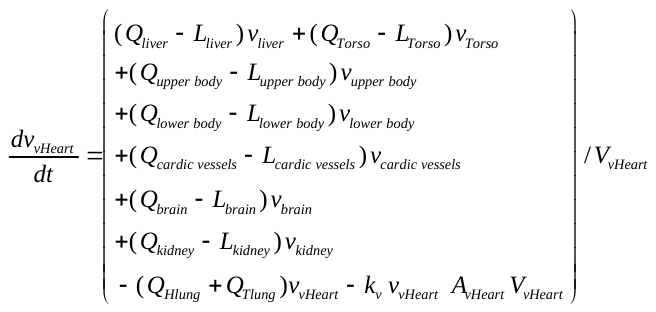 | (1b) |
| --- | --- |

where the first group of terms represents the recirculation in venous blood flow for free viruses and the last term describes the effect of the antibodies on the virus reproduction

1.1.2 Arterial and venous recirculation of micro-thrombi,
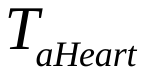
 and
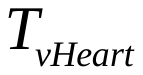
,

| 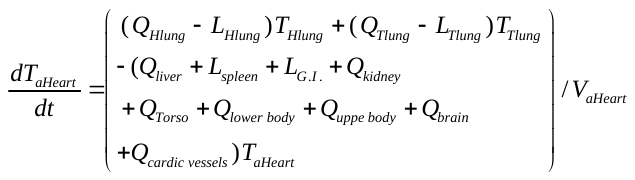 | (2a) |
| --- | --- |

where the terms represent the recirculation in arterial blood flow for micro-thrombosis

| 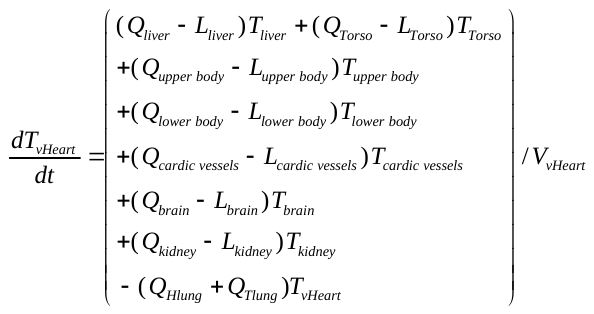 | (2b) |
| --- | --- |
| where the terms represent the recirculation in venous blood flow for micro-thrombosis |  |

1.1.3 Arterial and venous recirculation of antibody,
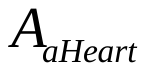
 and
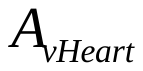
,

| 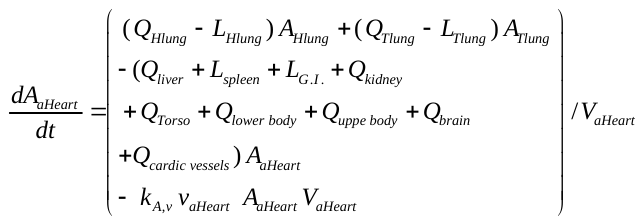 | (3a) |
| --- | --- |

where the first group of terms represents the recirculation in arterial blood flow for antibody and the last term describes the effect of the antibodies on the virus reproduction

| 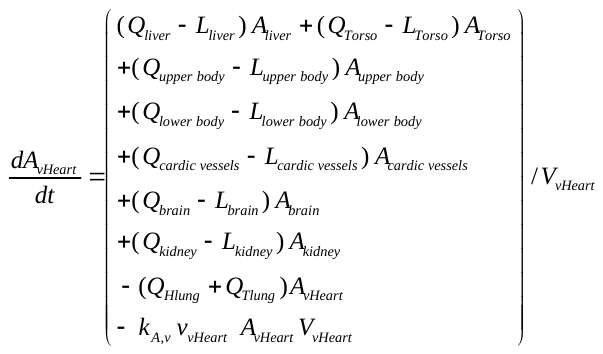 | (3b) |
| --- | --- |
| where the first group of terms represents the recirculation in venous blood flow for antibody and the last term describes the effect of the antibodies on the virus reproduction |  |

1.1.4 Arterial and venous recirculation of cytokines,
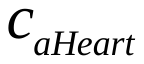
 and
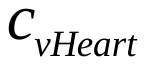
,

| 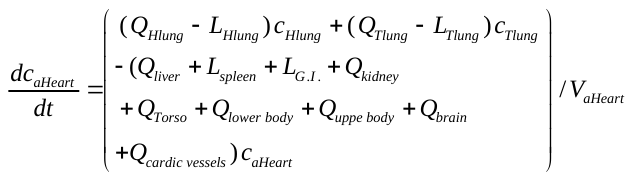 | (4a) |
| --- | --- |

where the first group of terms represents the recirculation in arterial blood flow for cytokines

| 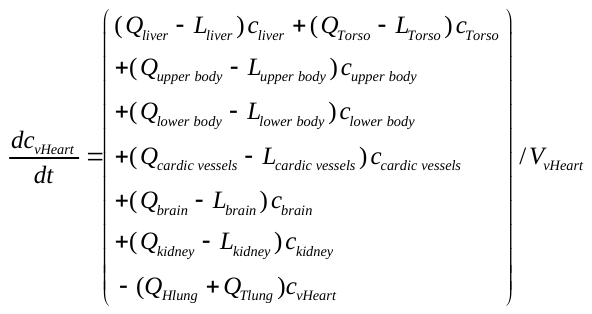 | (4b) |
| --- | --- |

where the first group of terms represents the recirculation in venous blood flow for cytokines

1.2 Liver

The liver compartment includes the hepatic portal vein from G.I. and spleen, and the hepatic artery to transport free viruses, antibody, and micro-thrombosis.

1.2.1 Free virus in liver vessels: hepatic portal vein from G.I. and spleen, and hepatic artery

| 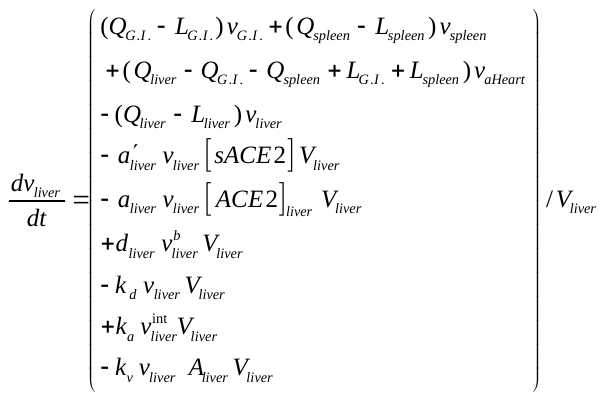 | (6a) |
| --- | --- |

where the first group of terms represents the recirculation in liver,
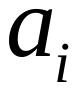
and are respectively the attachment rates of free virus to ACE2 and sACE2, *d* is detachment rate of bound virus from ACE2, is inactivating rate of the virus is the rate of virus release from the cell and the last term describes the effect of the antibodies on the virus reproduction

1.2.2 Virus bound to liver vessel wall

|  | (6b) |
| --- | --- |

where are the attachment rates of free virus to ACE2, *d* is the detachment rate of bound virus from ACE2, is the inactivating rate of the virus and  is the rate of bound virus internalization

1.2.3 Virus internalized into liver vessel endothelium

|  | (6c) |
| --- | --- |

where the first term describes replication, the second the internalization of the bound virus and the third term describes the viral particles that exit the cell.

1.2.4 Micro-thrombus formation and transport in liver

|  | (6d) |
| --- | --- |

where the first group of terms represents the recirculation in liver, , , and are the micro-thrombus-inhibiting coefficients of anti-virus drugs, anti-coagulation drugs for virus-ACE-2-induced coagulation, anti-coagulation drugs for IL6-induced coagulation,anti-coagulation drugs for cytokine storm-induced coagulation, is the attachment rate of micro-thrombus to vessels. is the dissolution of micro-thrombi due to thrombolysis.

1.2.5 Micro-thrombus accumulation in liver

|  | (6e) |
| --- | --- |

is the attachment rate of micro-thrombus to vessels and is the dissolution of micro-thrombi due to thrombolysis.

1.2.6 ACE2 density of liver

|  | (6f) |
| --- | --- |

where the first term is the detachment rate of bound virus from ACE2 and the second term is the binding of the virus on the ACE2 receptor.

1.2.7 Antibody transport in liver

|  | (6g) |
| --- | --- |

where the first group of terms represents the recirculation in liver for antibody and the following terms describes the effect of the antibodies on the virus reproduction and the infected endothelial cell interaction and the last term describes the degradation rate of antibodies

1.2.8 Cytokine transport in liver

|  | (6h) |
| --- | --- |

where the first group of terms represents the recirculation in liver for antibodies and the following terms describes the production of cytokines by the natural killer, antigen-presenting, effector CD4+ T cells, neutrophils, macrophages, infected endothelial cells, infected epithelial cells, the degradation rate of cytokines by Mas receptor, AT2 receptor and regulatory T cells, respectively, and the last term describes the degradation rate of cytokines

1.2.9 Healthy endothelial cells of liver vessels

|  | (6i) |
| --- | --- |

Where the first term describes the overall (“effective”) proliferation/death of endothelial cells and the second term is the conversion of healthy endothelial cells to infected endothelial cells by the internalized virus, and the last term is the conversion of healthy endothelial cells to infected endothelial cells by neutrophils and cytokines

1.2.10 Infected endothelial cells of liver vessels

|  | (6j) |
| --- | --- |

Where the first term is the conversion of healthy endothelial cells to infected endothelial cells by the internalized virus, and the second term is the conversion of healthy endothelial cells to infected endothelial cells by neutrophils and cytokines, and the last terms describe the overall (“effective”) death of infected endothelial cells.

1.3 For the i^th^ organ/tissue, where i = spleen, G.I., upper body, lower body, torso and cardiac vessels

All these compartments have upstream blood flow coming from the left ventricle of heart and downstream blood flow going back to the right ventricle of heart to allow both free viruses and micro-thrombosis to circulate in the whole body.

1.3.1 Free virus in the vessels of i^th^ organ

|  | (7a) |
| --- | --- |

where the first group of terms represents the recirculation in i^th^ organ, and are respectively the attachment rates of free virus to ACE2 and sACE2, *d* is the detachment rate of bound virus from ACE2, is the inactivating rate of the virus is the rate of virus released from the cell and the last term describes the effect of the antibodies on the virus reproduction

1.3.2 Bound virus on vessel wall of i^th^ organ

|  | (7b) |
| --- | --- |

where are the attachment rates of free virus to ACE2, *d* is the detachment rate of bound virus from ACE2, is the inactivating rate of the virus and is the rate of bound virus internalization.

1.3.3 Internalized virus in endothelium of i^th^ organ

|  | (7c) |
| --- | --- |

where the first term describes replication, the second the internalization of the bound virus and the third term describes the viral particles that exit the cell.

1.3.4 Micro-thrombus formation and transport in i^th^ organ

|  | (7d) |
| --- | --- |

where the first group of terms represents the recirculation in i^th^ organ , , , and are the micro-thrombus-inhibiting coefficients of anti-virus drugs, anti-coagulation drugs for virus-ACE-2-induced coagulation, anti-coagulation drugs for IL6-induced coagulation,anti-coagulation drugs for cytokine storm-induced coagulation, is the attachment rate of micro-thrombus to vessels. is the dissolution of micro-thrombi due to thrombolysis.

1.3.5 Micro-thrombus accumulation in i^th^ organ

|  | (7e) |
| --- | --- |

is the attachment rate of micro-thrombus to vessels. is the dissolution of micro-thrombi due to thrombolysis.

1.3.6 ACE2 density of i^th^ organ

|  | (7f) |
| --- | --- |

where the first term is the detachment rate of bound virus from ACE2 and the second term is the binding of the virus on the ACE2 receptor.

1.3.7 Antibody transport in i^th^ organ

|  | (7g) |
| --- | --- |

where the first group of terms represents the recirculation in ith organ for antibody and the following terms describe the effect of the antibodies on the virus reproduction and the infected endothelial cell interaction and the last term describes the degradation rate of antibodies

1.3.8 cytokine transport in i^th^ organ

|  | (7g) |
| --- | --- |

where the first group of terms represents the recirculation in ith organ for antibodies and the following terms describe the production of cytokines by the natural killer, antigen-presenting, effector CD4+ T cells, neutrophils, macrophages, infected endothelial cells, infected epithelial cells, the degradation rate of cytokines by Mas receptor, AT2 receptor and regulatory T cells, respectively, and the last term describes the degradation rate of cytokines

1.3.9 Healthy endothelial cells of liver vessels

|  | (7i) |
| --- | --- |

Where the first term describes the overall (“effective”) proliferation/death of endothelial cells and the second term is the conversion of healthy endothelial cells to infected endothelial cells by internalized virus, and the last term is the conversion of healthy endothelial cells to infected endothelial cells by neutrophils and cytokines.

1.3.10 Infected endothelial cells of liver vessels

|  | (7j) |
| --- | --- |

Where the first term is the conversion of healthy endothelial cells to infected endothelial cells by internalized virus, and the second term is the conversion of healthy endothelial cells to infected endothelial cells by neutrophils and cytokines, and the last terms describe the overall (“effective”) death of infected endothelial cells.

1.4 Lung

The lung compartment has upstream blood flow coming from heart and downstream blood flow going back to heart to transport both free viruses and micro-thrombi. In this study, the lung compartment is divided into two different parts: normal healthy lung (*Hlung*), tumor part of lung (*Tlung*). Free, bound and internalized virus concentrations in the lung are calculated using the microscale model of the lung, Eqs. 33-35. ACE2 concentration of the lung is calculated by Eq. 32 of the microscale model of lung. The microthrombus dynamics are:

Normal part of Lung :

1.1.1 Micro-thrombus formation and transport in healthy regions of lung

|  | (8a) |
| --- | --- |

where the first group of terms represents the recirculation in healthy regions of lung , , , and are the micro-thrombus-inhibiting coefficients of anti-virus drugs, anti-coagulation drugs for virus-ACE-2-induced coagulation, anti-coagulation drugs for IL6-induced coagulation,anti-coagulation drugs for cytokine storm-induced coagulation and NETs, is the dissolution of micro-thrombi due to thrombolysis, is the attachment rate of micro-thrombus to vessels.

1.1.2 Micro-thrombus accumulation in lung

|  | (8b) |
| --- | --- |

is the attachment rate of micro-thrombus to vessels. is the dissolution of micro-thrombi due to thrombolysis.

1.1.3 Antibody transport in lung

|  | (8c) |
| --- | --- |

where the first group of terms represents the recirculation in healthy regions of lung for antibodies, the following terms describe the antibodies production by long and short-lived plasma respectively, the effect of the antibodies on the virus reproduction and the infected endothelial and epithelial cell interaction and the last term describes the degradation rate of antibodies

1.1.4 cytokine transport in lung

|  | (8d) |
| --- | --- |

where the first group of terms represents the recirculation in healthy regions of lung for antibodies and the following terms describe the production of cytokines by the natural killer, antigen-presenting, effector CD4+ T cells, neutrophils, macrophages, infected endothelial cells, infected epithelial cells, the degradation rate of cytokines by MAs receptor, AT2 receptor and regulatory T cells, respectively, and the last term describes the degradation rate of cytokines

1.1.5 Healthy endothelial cells of lung vessels

|  | (8e) |
| --- | --- |

Where the first term describes the overall (“effective”) proliferation/death of endothelial cells and the second term is the conversion of healthy endothelial cells to infected endothelial cells by internalized virus, and the last term is the conversion of healthy endothelial cells to infected endothelial cells by neutrophils and cytokines.

1.1.6 Infected endothelial cells of lung vessels

|  | (8f) |
| --- | --- |

Where the first term is the conversion of healthy endothelial cells to infected endothelial cells by internalized virus, and the second term is the conversion of healthy endothelial cells to infected endothelial cells by neutrophils and cytokines, and the last terms describe the overall (“effective”) death of infected endothelial cells.

1.1.7 Healthy epithelial cells of lung vessels

|  | (8g) |
| --- | --- |

Where the first term describes the overall (“effective”) proliferation/death of epithelial cells and the second term is the conversion of healthy epithelial cells to infected epithelial cells by internalized virus, and the last term is the conversion of healthy epithelial cells to infected endothelial cells by neutrophils and cytokines.

1.1.8 Infected epithelial cells of lung vessels

|  | (8h) |
| --- | --- |

Where the first term is the conversion of healthy endothelial cells to infected epithelial cells by internalized virus, and the second term is the conversion of healthy epithelial cells to infected endothelial cells by neutrophils and cytokines, and the last terms describe the overall (“effective”) death of infected epithelial cells.

Tumor part of lung:

1.1.9 Micro-thrombus formation and transport in lung tumor

|  | (8a) |
| --- | --- |

where the first group of terms represent the recirculation in lung tumor , , , and are the micro-thrombus-inhibiting coefficients of anti-virus drugs, anti-coagulation drugs for virus-ACE-2-induced coagulation, anti-coagulation drugs for IL6-induced coagulation,anti-coagulation drugs for cytokine storm-induced coagulation and NETs, is the dissolution of micro-thrombi due to thrombolysis, is the attachment rate of micro-thrombus to vessels.

1.1.10 Micro-thrombus accumulation in lung tumor

|  | (8b) |
| --- | --- |

is the attachment rate of micro-thrombus to vessels. is the dissolution of micro-thrombi due to thrombolysis.

1.1.11 Antibody transport in lung tumor

|  | (8c) |
| --- | --- |

where the first group of terms represents the recirculation in lung tumor for antibodies, the following terms describe the antibodies production by long and short-lived plasma respectively, the effect of the antibodies on the virus reproduction and the infected endothelial and epithelial cell interaction and the last term describes the degradation rate of antibodies.

1.1.12 cytokine transport in the tumor part of lung

|  | (8d) |
| --- | --- |

where the first group of terms represents the recirculation in lung tumor for antibodies and the following terms describe the production of cytokines by the natural killer, antigen-presenting, effector CD4+ T cells, neutrophils, macrophages, infected endothelial cells, infected epithelial cells, the degradation rate of cytokines by MAs receptor, AT2 receptor and regulatory T cells, respectively, and the last term describes the degradation rate of cytokines.

1.1.13 Healthy endothelial cells of the vessels of lung tumor

|  | (8e) |
| --- | --- |

Where the first term describes the overall (“effective”) proliferation/death of endothelial cells and the second term is the conversion of healthy endothelial cells to infected endothelial cells by internalized virus, and the last term is the conversion of healthy endothelial cells to infected endothelial cells by neutrophils and cytokines.

1.1.14 Infected endothelial cells of the vessels of lung tumor

|  | (8f) |
| --- | --- |

Where the first term is the conversion of healthy endothelial cells to infected endothelial cells by internalized virus, and the second term is the conversion of healthy endothelial cells to infected endothelial cells by neutrophils and cytokines, and the last terms describe the overall (“effective”) death of infected endothelial cells.

1.1.15 Healthy epithelial cells of the vessels of lung tumor

|  | (8g) |
| --- | --- |

Where the first term describes the overall (“effective”) proliferation/death of epithelial cells and the second term is the conversion of healthy epithelial cells to infected epithelial cells by internalized virus, and the last term is the conversion of healthy epithelial cells to infected endothelial cells by neutrophils and cytokines.

1.1.16 Infected epithelial cells of the vessels of lung tumor

|  | (8h) |
| --- | --- |

Where the first term is the conversion of healthy endothelial cells to infected epithelial cells by internalized virus, and the second term is the conversion of healthy epithelial cells to infected endothelial cells by neutrophils and cytokines, and the last terms describe the overall (“effective”) death of infected epithelial cells.

1.2 Mass conservation

|  | (9) |
| --- | --- |

**Simplifying Assumptions:**

**T_E_,** Ma, N, a (Anti-inflammatory cytokines), , IL6, IF, T^M^ are uniform.

Antibodies are released in lung and then circulate. There is a production term just for lung. But we could assume uniform short- and Long-lived B cells and add production terms to each compartment.

2. Microscale Lung
   2.1 Equations describing the Renin-Angiotensin system

The reaction of angiotensinogen (AGT) is governed by Eq. 7,(1)

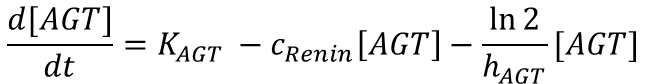
(10)

The rate of change of angiotensinogen depends on its production (K_AGT_); production of ANG I catalyzed by renin is assumed to follow first order kinetics, and thus PRA = c_Renin_[AGT], degradation of AGT is considered to exhibit first-order kinetics in terms of its half-life h_AGT_.

The mass balance for Renin is (1)

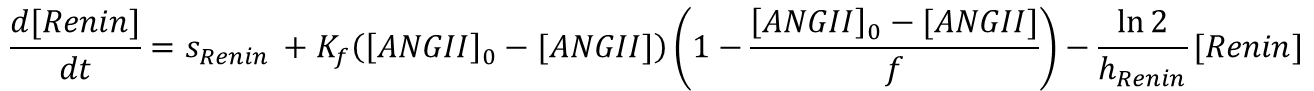
(11)

The first term, s_Renin_ , accounts for a constant source of renin from the kidney; the second term is the influence of ANG II negative feedback on renin production, [ANG II]_0_ is the initial concentration of ANG II, k_f_ and f are parameters for the feedback, and h_Renin_ is the half-life for the degradation of renin.

$$s_{Renin}=\frac{\ln2}{h_{Renin}}{[Renin]}_{0}$$

(12)

The renin source term is computed at steady state, where [Renin]_0_ is the initial concentration of renin.

The mass balance for ANG I is(1)

$\frac{d[ANGI]}{dt}=c_{Renin}\left[ AGT \right]+K_{Renin}\left( {\left[ Renin \right]-\left[ Renin \right]}_{0} \right)-K_{ACE}\left[ ANGI \right]-K_{NEP}ANGI-K_{ACE2-ANGI}^{on}\left[ ANGI \right]\left[ ACE2 \right]+K_{ACE2-ANGI}^{off}\left[ ACE2-ANGI \right]-\frac{\ln2}{h_{ANGI}}[ANGI]$	(13)

where the first term represents the glucose-dependent renin-catalyzed contribution to the production of ANG I from AGT, the second term represents the change to ANG I synthesis from AGT due to the feedback of ANG II on renin with rate constant K_Renin_, the third term is the ACE-catalyzed conversion of ANG I to ANG II and has a glucose-dependent rate parameter K_ACE_, the fourth term is the consumption of ANG I to form ANG-(1-7) with the glucose-independent rate parameters K_NEP_, the following two terms describe the binding/unbinding of ANG I on the ACE 2 receptors, and h_ANG I_ is the half-life for the degradation of ANG I.

The mass balance for the free ANG II is(1)

$$\frac{d[ANGII]}{dt}=K_{ACE}\left[ ANGI \right]-K_{AT1}^{on}\left[ ANGII \right]\left[ AT1R \right]+K_{AT1}^{off}\left[ AT1R-ANGII \right]-K_{AT2}^{on}\left[ ANGII \right]\left[ AT2R \right]+K_{AT2}^{off}\left[ AT2R-ANGII \right]-K_{APA}\left[ ANGII \right]-K_{ACE2-ANGII}^{on}\left[ ANGII \right]\left[ ACE2 \right]+ K_{ACE2-ANGII}^{off}[ACE2-ANGII]-\frac{\ln2}{h_{ANGII}}[ANGII]$$

(14)

where the first term is the production of ANG II in the presence of ACE, the following two terms

The mass balance for ANGII bound to AT1 receptor is

(15)

$$\frac{d[AT1R-ANGII]}{dt}=K_{AT1}^{on}\left[ ANGII \right]\left[ AT1R \right]-K_{AT1}^{off}[AT1R-ANGII]-\frac{\ln2}{h_{AT1R-ANGII}}[AT1R-ANGII]$$

where the first two terms describe the binding/unbinding of ANG II on the AT1 receptors and hAT1R-ANGII is the degradation half-life.

The mass balance for ANGII bound to AT2 receptor is(2)

(16)

$$\frac{d[AT2R-ANGII]}{dt}=K_{AT2}^{on}\left[ ANGII \right]\left[ AT2R \right]-K_{AT2}^{off}[AT2R-ANGII]-\frac{\ln2}{h_{AT2R-ANGII}}[AT2R-ANGII]$$

where the first two terms describe the binding/unbinding of ANG II on the AT2 receptor and has a degradation half-life h_AT2R-ANGII_.

The mass balance for ANG(1-7) is(2)

$$\frac{d[{ANG}_{(1-7)}]}{dt}=K_{NEP}\left[ ANGI \right]+K_{ACE2} \left[ ACE2-ANGII \right] {+K}_{ANG\left( 1-9 \right)} \left[ ANG\left( 1-9 \right) \right]-K_{MAs}^{on}\left[ ANG\left( 1-7 \right) \right]\left[ MAsR \right]+K_{MAs}^{off}[MAsR-ANG(1-7)]-\frac{\ln2}{h_{{ANG}_{(1-7)}}}[{ANG}_{(1-7)}]$$

(17)

where the first three terms are the production of ANG(1-7) by ANGI, ANGII bound to ACE2, and by ANG(1-9), the following two terms describe the binding/unbinding of ANG 1-7 to the MAs receptors and h_ANG 1-7_ is the half-life for degradation of ANG 1-7.

The mass balance for ANG(1-7) bound to Mas receptor is

(18)

$$\frac{d[MAsR-ANG(1-7)]}{dt}=K_{MAs}^{on}\left[ ANG(1-7) \right]\left[ MAsR \right]-K_{MAs}^{off}[MAsR-ANG(1-7)]-\frac{\ln2}{h_{MAsR-ANG\left( 1-7 \right)}}[MAsR-ANG(1-7)]$$

where the first two terms describe the binding/unbinding of ANG 1-7 on the MAs receptor and has a degradation half-life of h_MasR-ANG1-7_.

The mass balance of ANG (1-9) is(2)

(19)

$$\frac{d[ANG(1-9)]}{dt}=K_{ACE2}\left[ ACE2-ANGI \right]-\frac{\ln2}{h_{ANG\left( 1-9 \right)}}[ANG(1-9)]$$

where the first term is the production of ANG(1-9) by ANGI bound to the ACE2 and h_ANG 1-9_ is the half-life for degradation of ANG 1-9.

The mass balance term of ANGIII is(2)

(20)

$$\frac{d[ANGIII]}{dt}=K_{APA} \left[ ANGII \right] -K_{APM} [ANGIII]-\frac{\ln2}{h_{ANGIII}}[ANGIII]$$

where the first term is the production of ANGIII by ANG II, the second term describes the production of ANGIV by ANGIII and h_ANG III_ is the half-life for degradation of ANG III.

The mass balance for ANGIV is(3)

(21)

$$\frac{d[ANGIV]}{dt}=K_{APM} \left[ ANGIII \right] -K_{AT4}^{on}\left[ ANGIV \right]\left[ AT4R \right]+K_{AT4}^{off}[AT4R-ANGIV]-\frac{\ln2}{h_{ANGIV}}[ANGIV]$$

where the first term is the production of ANG IV by ANG III, the following two terms describe the binding/unbinding of ANG IV on the AT4 receptor and h_ANGIV_ the half-life for degradation of ANG IV.

The mass balance for ANGIV bound to AT4 receptor is

(22)

$$\frac{d[AT4R-ANGIV]}{dt}=K_{AT4}^{on}\left[ ANGIV \right]\left[ AT4R \right]-K_{AT4}^{off}[AT4R-ANGIV]-\frac{\ln2}{h_{AT4R-ANGIV}}[AT4R-ANGIV]$$

where the first two terms describe the binding/unbinding of ANG IV on the AT4 receptors and has a degradation half-life h_AT4R-ANGIV_.

The mass balance for ANGI bound to ACE2 is

(23)

$$\frac{d\left[ ACE2-ANGI \right]}{dt}=K_{ACE2-ANGI}^{on}\left[ ANGI \right]\left[ ACE2 \right]-K_{ACE2-ANGI}^{off}\left[ ACE2-ANGI \right]-K_{ACE2}\left[ ACE2-ANGI \right]$$

where the first two terms describe the binding/unbinding of ANGI on the ACE2 and the last term describes the degradation of ANGI bound to ACE2.

Mass balance for ANGII bound to ACE2:

(24)

${\frac{d\left[ ACE2-ANGII \right]}{dt}=K}_{ACE2-ANGII}^{on}\left[ ANGII \right]\left[ ACE2 \right]- K_{ACE2-ANGII}^{off}\left[ ACE2-ANGII \right]-K_{ACE2} \left[ ACE2-ANGII \right]$

where the first two terms describe the binding/unbinding of ANGII on the ACE2 and the last term describes the degradation of the ANGII bound to ACE2.

The mass balance for AT1 receptor is

(25)

$$\frac{d[AT1R]}{dt}=S_{AT1R}-K_{AT1}^{on}\left[ ANGII \right]\left[ AT1R \right]+K_{AT1}^{off}\left[ AT1R-ANGII \right]-d_{AT1R}[AT1R]$$

where the first term represents the source term for AT1 receptor, the following two terms describe the binding/unbinding of ANG II on the AT1 receptor, and the last term describes the degradation of the AT1 receptor.

The mass balance for AT2 receptor is

(26)

$$\frac{d[AT2R]}{dt}=S_{AT2R}-K_{AT2}^{on}\left[ ANGII \right]\left[ AT2R \right]+K_{AT2}^{off}[AT2R-ANGII]-d_{AT2R}[AT2R]$$

where the first term represents the source term for AT2 receptor, the following two terms describe the binding/unbinding of ANG II on the AT2 receptors, and the last term describes the degradation of the AT2 receptor.

The mass balance for MAs receptor is

(27)

$$\frac{d[MAsR]}{dt}=S_{MAsR}-K_{MAs}^{on}\left[ ANG\left( 1-7 \right) \right]\left[ MAsR \right]+K_{MAs}^{off}[MAsR-ANG(1-7)]-d_{MAsR}[MAsR]$$

where the first term represents the source term for MAs receptor, the following two terms describe the binding/unbinding of ANG 1-7 on the MAs receptors, and the last term describes the degradation of the MAs receptor.

The mass balance for AT4 receptor is

(28)

$$\frac{d[AT4R]}{dt}=S_{AT4R}-K_{AT4}^{on}\left[ ANGIV \right]\left[ AT4R \right]+K_{AT4}^{off}[AT4R-ANGIV]-d_{AT4R}[AT4R]$$

where the first term represents the source term for AT4 receptor, the following two terms describe the binding/unbinding of ANG IV on the AT4 receptors, and the last term describes the degradation of the AT4 receptor.

2.2 Mass Balance Equations for IL6, IL6 receptor, soluble IL6R, binding of IL6 to IL6R and sIL6R and production of VEGF

The reaction rate of the IL-6 reads

$$\frac{d\left[ IL6 \right]}{dt}=K_{IL6-Nk}Nk+K_{IL6-DC}{DC}^{*}+K_{IL6-TE}\left( {Th}^{E}+T^{E} \right)+K_{IL6-M1}M_{1}-\gamma_{IL6}\left[ IL6 \right]$$

$$-K_{IL6}^{on}\left[ IL6 \right]\left[ IL6R \right]+K_{IL6}^{off}\left[ IL6R-IL6 \right]{-K_{sIL6R}^{on}\left[ sIL6R \right]\left[ IL6 \right]+K}_{sIL6R}^{off}\left[ sIL6R-IL6 \right]-K_{sIL6R}^{on}\left[ sIL6R \right]\left[ IL6 \right]$$

(29)

where the first four terms are the production of IL-6 by the natural killer, activated dendritic, and effector T cells, and the type 1 macrophages, respectively. The fifth term is the degradation of the IL-6. The rest terms describe the binding/unbinding of IL-6 to the IL-6 receptor and soluble IL-6 receptor.

The reaction rate of the IL-6 receptor reads

$$\frac{d[IL6R]}{dt}=S_{IL6R}-K_{IL6}^{on}\left[ IL6 \right]\left[ IL6R \right]+K_{IL6}^{off}\left[ IL6R-IL6 \right]-K_{sIL6R}\left[ IL6R \right]-d_{IL6R}[IL6R]$$

(30)

where the first term represents the source term for IL-6 receptor, the following two terms describe the binding/unbinding of IL-6 to the IL-6 receptors and the last terms are the transformation to soluble IL-6 receptors and the degradation of IL-6 receptors respectively.

The reaction rate of the soluble IL-6 receptor reads

$$\frac{d[sIL6R]}{dt}=K_{sIL6R}\left[ IL6R \right]+K_{sIL6R}^{off}[sIL6R-IL6]-K_{sIL6R}^{on}\left[ sIL6R \right]\left[ IL6 \right]$$

(31)

where the first term is the transformation of IL-6 receptor to the soluble IL-6 receptor and the last terms describe the binding/unbinding of IL6 to the soluble IL-6 receptors.

The reaction rate of the IL-6 - IL-6 receptor complex reads

$$\frac{d[IL6R-IL6]}{dt}=K_{IL6}^{on}\left[ IL6 \right]\left[ IL6R \right]-K_{IL6}^{off}[IL6R-IL6]-\frac{\ln2}{h_{IL6R}}[IL6R-IL6]$$

(32)

where the first two terms describe the binding/unbinding of IL-6 to IL-6 receptor and the last term is the degradation of IL-6-IL-6 receptor complex (with a half-life $h_{IL6R}$)

The reaction rate of the IL-6-soluble receptor - IL-6 complex reads

$$\frac{d\left[ sIL6R-IL6 \right]}{dt}=K_{sIL6R}^{on}\left[ sIL6R \right][IL6]{-K}_{sIL6R}^{off}\left[ sIL6R-IL6 \right]$$

(33)

where the two terms describe the binding/unbinding of IL-6 to the soluble IL-6 receptor.

The mass balance for VEGF

$$\frac{d\left[ VEGF \right]}{dt}=\nabla\cdot\left( D_{VEGF}\nabla\left[ VEGF \right] \right)+K_{T}G_{a}\left( c_{ox} \right)T+K_{T_{reg}}G_{a}\left( c_{ox} \right)T_{reg}+K_{VEGF}\left[ sIL6R-IL6 \right]-d_{VEGF}\left[ VEGF \right]+\gamma_{VEGF}\left( 100-SPO2 \right)$$

VEGF is assumed to be produced by cancer cells and *T_regs_* and its production is enhanced under hypoxic conditions as described by the oxygen tension term *G_a._* The following term describes the production of VEGF by IL-6 bound on the soluble IL-6 receptor, the following term is the degradation rate of the VEGF and the last term describes the production of VEGF by hypoxia

(34)

| *G_a_(*$\hat{c_{ox}}$*)=* | *3*$\hat{c_{ox}}$ *for 0<*$\hat{c_{ox}}$*<0.5 (hypoxia)*  *2 -*$\hat{c_{ox}}$ *for 0.5<*$\hat{c_{ox}}$*<1 (normoxia)*  $\hat{c_{ox}}$ *for 1<*$\hat{c_{ox}}$ *(hyperoxia)* |
| --- | --- |

2.3 Model equations for virus infection and immune cells activation

The mass balance for soluble ACE2 receptor

(35)

$$\frac{d[sACE2]}{dt}=- K_{sACE2}\left[ sACE2 \right]\left[ V \right]-\sum_{i} K_{sACE2}\left[ sACE2 \right]\left[ v_{i} \right]+K_{Adam17}[ACE2]-d_{sACE2}\left[ sACE2 \right]$$

where the first term is the binding of soluble ACE2 receptor to the virus, the third term is internalization everywhere, and the following term is the production of soluble ACE2 receptor by ACE2 receptor interaction through Adam17 and the last term is the degradation rate of the soluble ACE2 receptor.

The mass balance of ACE2 receptor is

(36)

$\frac{d[ACE2]}{dt}=S_{ACE2}(\left[ EC \right]+H)-K_{ACE2-Virus}^{on}\left[ v \right]\left[ ACE2 \right]+K_{ACE2-Virus}^{off}\left[ v_{b} \right]-K_{Adam17}[ACE2]-K_{ACE2-ANGI}^{on}\left[ ANGI \right]\left[ ACE2 \right]+K_{ACE2-ANGI}^{off}\left[ ACE2-ANGI \right]-K_{ACE2-ANGII}^{on}\left[ ANGII \right]\left[ ACE2 \right]+ K_{ACE2-ANGII}^{off}[ACE2-ANGII]$

where the first term describes the production of ACE2 receptor by endothelial and epithelial cells, the second and third term describes the interaction with the virus, the fourth term describes the production of soluble ACE2 and the rest terms describe the interaction of ACE2 with ANGI and ANGII.

The virus can be in three states, the free virus that diffuses in the lung tissue, the bound virus on the epithelial cells of the lungs and the internalized virus. The equations for the three states of the virus are:

Free virus in normal Lung

$$\frac{dv_{Hlung}}{dt}=\frac{Q_{Hlung}v_{Hlung}}{V_{Hlung}}-\frac{\left( Q_{Hlung}-L_{Hlung} \right)v_{Hlung}}{V_{Hlung}}-K_{sACE2}\left[ v_{Hlung} \right]\left[ sACE2 \right]-K_{ACE2-Virus}^{on}\left[ v_{Hlung} \right]\left[ ACE2 \right]+K_{ACE2-Virus}^{off}\left[ v_{b} \right]-K_{d}v_{Hlung}+K_{a}{\left( 1-K_{IF} \frac{IF}{K_{IF}+IF} \right) v}_{int}-K_{v}v_{Hlung}A_{Hlung}$$

(37)

Where the terms respectively describe convectional transport of free virus in lung, the binding of the virus to soluble ACE2, the binding of the virus to ACE2, the detachment of bound virus from ACE2, the inactivating of the virus, and the source of new viral particles from the release of the infected cells following cell death.

The effect of the antibodies on the virus reproduction is represented by the term $K_{v}vA$ (20)(3).

Free virus in Tumor part of lung

$$\frac{dv_{Tlung}}{dt}=\frac{Q_{Tlung}v_{Tlung}}{V_{Tlung}}-\frac{\left( Q_{Tlung}-L_{Tlung} \right) v_{Tlung}}{V_{Tlung}}-K_{sACE2}\left[ v_{Tlung} \right]\left[ sACE2 \right]-K_{ACE2-Virus}^{on}\left[ v_{Tlung} \right]\left[ ACE2 \right]+K_{ACE2-Virus}^{off}\left[ v_{b} \right]-K_{d}v_{Tlung}+K_{a}{\left( 1-K_{IF} \frac{IF}{K_{IF}+IF} \right) v}_{int}-K_{v}v_{Tlung}A_{Tlung}$$

(38)

Where the terms respectively describe convectional transport of free virus in tumor, the binding of the virus to soluble ACE2, the binding of the virus to ACE2, the detachment of bound virus from ACE2, the inactivating of the virus, and the source of new viral particles from the release of the infected cells following cell death.

The effect of the antibodies on the virus reproduction is represented by the term $K_{v}vA$ (20)(3).

Bound virus in normal lung

(39)

$$\frac{dv_{Hlung}^{b}}{dt}=K_{ACE2-Virus}^{on}\left[ v_{Hlung} \right]\left[ ACE2 \right]-K_{ACE2-Virus}^{off}\left[ v_{Hlung}^{b} \right]-K_{d} v_{Hlung}^{b}-Kintv_{Hlung}^{b}$$

where the first two terms describe the binding/unbinding of the virus on the ACE2 receptor, the third term is the degradation of the bound virus, the last term is the internalization of the bound virus into the cells.

Internalized virus in normal lung

$$\frac{dv_{Hlung}^{int}}{dt}={P_{lung}{k_{avp} v_{Hlung}^{int}}+K}_{int}v_{Hlung}^{b}-K_{a} \left( 1-K_{IF} \frac{IF}{K_{IF}+IF} \right)v_{Hlung}^{int}$$

(40)

where the first term describes replication, the second the internalization of the bound virus and the third term describes the viral particles that exit the cell.

Bound virus in tumor part of lung

(39)

$$\frac{dv_{Tlung}^{b}}{dt}=K_{ACE2-Virus}^{on}\left[ v_{Tlung} \right]\left[ ACE2 \right]-K_{ACE2-Virus}^{off}\left[ v_{Tlung}^{b} \right]-K_{d}v_{Tlung}^{b}-Kint v_{Tlung}^{b}$$

where the first two terms describe the binding/unbinding of the virus on the ACE2 receptor, the third term is the degradation of the bound virus, the last term is the internalization of the bound virus into the cells.

Internalized virus in tumor part of lung

$$\frac{dv_{Tlung}^{int}}{dt}={P_{lung}{k_{avp} v_{Tlung}^{int}}+K}_{int}v_{Tlung}^{b}-K_{a} \left( 1-K_{IF} \frac{IF}{K_{IF}+IF} \right)v_{Tlung}^{int}$$

(40)

where the first term describes replication of virus in tumor region of lung, the second term is the internalization of the bound virus and the third term describes the viral particles that exit the tumor cell.

2.4 Interferon gamma ($\boldsymbol{IF}\boldsymbol{N}_{\boldsymbol{\gamma}}$)

The reaction rate of the interferon gamma reads

$$\frac{d{IFN}_{\gamma}}{dt}=k_{IFN_{\gamma} NK}NK+k_{IFN_{\gamma} {Th}^{E}}{Th}^{E}+k_{IFN_{\gamma} T^{E}}T^{E}+a_{IF} \left( IEC+In \right)+k_{IFN_{\gamma}}- \varepsilon_{IF} {IFN}_{\gamma}$$

(41)

where the first five terms describe the production rate of the interferon gamma by the natural killer, effector CD4^+^ T, effector CD8^+^ T, infected endothelial, infected epithelial, and other cells, respectively. The last term describes the degradation rate of interferon gamma.

2.5 Anti-inflammatory cytokines

The reaction rate of the anti-inflammatory cytokines reads

$$\frac{da}{dt}=k_{a-N}N*\left( M_{a}+DC \right)+K_{Ang1-7}\left[ MAsR-{ANG}_{\left( 1-7 \right)} \right]+K_{ant-T_{reg}}T_{reg}+K_{ant-Ma}M_{a}-\gamma_{a}a$$

(42)

where the first term describes the production of anti-inflammatory cytokines by type 2 macrophages and dendritic cells after phagocytosis of apoptotic neutrophils. The other three terms are the production by ANG(1-7)-MasR complex, regulatory T cells and type 2 macrophages respectively. The last term is the degradation rate of anti-inflammatory cytokines.

2.6 Reactions of PDL-1, PD-1, and anti-PD-1

The reaction of the PDL-1 reads

$$\frac{d[PDL1]}{dt}=\left[ \left( \lambda_{H}-d_{H} \right)H+\left( \lambda_{EC}-d_{EC} \right)\left[ EC \right]+\left( \lambda_{T}-d_{T} \right)\hat{T} \right]\frac{\left[ PDL1 \right]}{H+\left[ EC \right]+\hat{T}}+h_{PDL1}T_{E}+h_{M2}M_{a}$$

(43)

where the first term describes the production(*λ*_i_)/degradation(*d*_i_) of the PD1 ligand by healthy epithelial, healthy endothelial cells and tumor cells, the last two terms describe the production of the PD1 ligand by Effector (Activated) T cells and type 2 macrophages.

The reaction of the PD-1 reads

$$\frac{d\left[ PD1 \right]}{dt}=\left[ \left( \lambda_{TE}-d_{TE} \right)T_{E}+\left( \lambda_{TN}-d_{TN} \right)T_{N}+\left( \lambda_{N}-d_{N} \right)N+\left( \lambda_{NK}-d_{NK} \right)NK+\left( \lambda_{M2}-d_{M2} \right)M_{a} \right]\frac{\left[ PD1 \right]}{T_{E}+T_{N}+N+NK+M_{a}}-\mu_{PD1-aPD1}\left[ PD1 \right]\left[ anti-PD1 \right]$$

(44)

where the first describes the production(*λ*_i_)/degradation(*d*_i_) of the PD1 by Effector (Activated) T cells, Naïve T cells, neutrophils, natural killer cells and macrophages. The last term describes the binding of the anti-PD1 to PD1.

The reaction of the PD-1 – PDL-1 complex reads

$$\frac{d\left[ PD1-PDL1 \right]}{dt}=a_{PL}\left[ PD1 \right]\left[ PDL1 \right]-d_{Q}\left[ PD1-PDL1 \right]$$

(45)

where the first term describes the binding of PD1 to the PD1 ligand and the last term describes its degradation rate.

The reaction of the anti-PD-1 reads

$$\frac{d\left[ anti-PD1 \right]}{dt}=\gamma_{A}-\mu_{PD1-aPD1}\left[ PD1 \right]\left[ anti-PD1 \right]-d_{A}\left[ anti-PD1 \right]$$

(46)

where the first term represents the source term of the anti-PD1, the second term describes the production of the anti-PD1 - PD1 complex and the last term describes its degradation rate.

2.7 Cells of immune system
    2.7.1 Neutrophils

The reaction rate of the neutrophils reads

$$\frac{dN}{dt}=\frac{\chi_{N} c}{1+a}+\chi_{N-IL6}\left[ IL6R-IL6 \right]-\gamma_{n}N*(1+\beta_{n}\frac{v}{v+1})$$

(47)

Where the production rate of the neutrophils depends on cytokines (pro/anti-inflammatory) and IL-6 - IL-6R complex, and the last term describes their death rate.

2.7.2 Neutrophils Extracellular Traps (NETs)

(48)

$$\frac{d[NETs]}{dt}=\gamma nN*\left( 1+\beta_{n}\frac{v}{v+1} \right)-\gamma_{NETs}[NETs]$$

where the first term describes the production Neutrophil extracellular traps and the last term is their degradation rate.

2.7.3 Immature dendritic cells (DC)

The reaction rate of the immature dendritic cells reads

$$\frac{dDC}{dt}=S_{DC}-d_{DC}DC-\left[ \beta_{D}DC \left( v+V_{proteins} \right)+\frac{\chi_{DC} c}{1+a} DC \overset{̑}{T} \right]- k_{Treg-DC} T_{reg} DC$$

(49)

where the first term describes the source term of DC and the second term is the death rate of DC, the rest term describes the conversion of dendritic cells to antigen presenting cells and the last term describes the degradation rate of DC by regulatory T cells.

2.7.4 Antigen presenting cells APCs (*DC^*^*)

The reaction rate of the APCs reads

$$\frac{d{DC}^{*}}{dt}=\left[ \beta_{D}DC \left( v+V_{proteins} \right)+\frac{\chi_{DC} c}{1+a} DC \overset{̑}{T} \right]-k_{Treg-DC^{*}}T_{reg}DC^{*}-\delta_{DC*}{DC}^{*}$$

(50)

where the first two terms describe the conversion rates of dendritic cells to APCs, , the rest term describes the degradation rate of APCs by regulatory T cells and the last term describes the death rate of APCs.

2.7.5 Naïve CD4^+^ T cells (*Th^N^*)

The reaction rate of the Naive CD4^+^ T cells reads

$$\frac{d{Th}^{N}}{dt}=S_{ThN}-h_{Th}{DC}^{*}{Th}^{N}\left[ \left( \frac{V_{proteins}}{1+V_{proteins}} \right)+\left( \frac{As}{1+As} \right) \right]\left( \frac{c}{1+c} \right)\left( \frac{{IFN}_{\gamma}}{K_{IF}+{IFN}_{\gamma}} \right) \frac{K_{T}}{K_{T}+\left[ PD1-PDL1 \right]}-d_{TN}{Th}^{N}-\frac{h_{Treg}DC {Th}^{N}a}{\left( K_{Th^{N} DC}+DC \right)\left( K_{DC Th^{N}}+{Th}^{N} \right)\left( K_{T_{reg} a}+a \right)}$$

(51)

where the first term describes the source term of Naïve CD4^+^ T cells, the second term describes the activation rate of naive CD4^+^ T cells, the following term describes the death rate of naïve CD4^+^ T cells and the last term describes its transformation to regulatory T cells.

2.7.6 Effector CD4^+^ T cells (*Th^E^*)

The reaction rate of the effector CD4^+^ T cells reads

$$\frac{d{Th}^{E}}{dt}=h_{Th} {DC}^{*}{Th}^{N}\left[ \left( \frac{V_{proteins}}{1+V_{proteins}} \right)+\left( \frac{As}{1+As} \right) \right]\left( \frac{c}{1+c} \right)\left( \frac{{IFN}_{\gamma}}{K_{IF}+{IF}_{\gamma}} \right) \frac{K_{T}}{K_{T}+\left[ PD1-PDL1 \right]}+\rho_{N}{Th}^{E}\left( \frac{As}{1+As} \right)\left( \frac{c}{1+c} \right)\left( \frac{{IFN}_{\gamma}}{K_{IF}+{IFN}_{\gamma}} \right) \frac{K_{T}}{K_{T}+\left[ PD1-PDL1 \right]}-\pi_{ThM}{Th}^{E}-\varepsilon{Th}^{E}\frac{\left[ PD1-PDL1 \right]}{AS}-\frac{k_{Treg_{1}}{Th}^{E}T_{reg}}{(1+{Th}^{E})(1+T_{reg})}$$

(52)

where the first term describes the activation rate of naïve CD4^+^ T cells, the second term describes the proliferation rate of effector CD4^+^ T cells, the third term describes the conversion rate of effector CD4^+^ T cells to memory CD4^+^ T cells and the last two terms describe their death rates which depend on the PD1 – PDL1 complex and the regulatory T cells.

2.7.7 Regulatory T cells (*Treg*)

The reaction rate of the regulatory T cells reads

$$\frac{dT_{reg}}{dt}=S_{Treg}+\rho_{Treg} T_{reg} \left( \frac{a}{1+c} \right)-\pi_{Treg}T_{reg}$$

(53)

where the first term describes their production rate from the thymus. The next term is their proliferation rate due to cytokines (cytokines and anti -inflammatory). The last term describes their death rate.

2.7.8 Naïve CD8^+^ T cells (*T^N^*)

The reaction rate of the naïve CD8^+^ T cells reads

$$\frac{dT^{N}}{dt}=S_{TN}-h_{T}{DC}^{*}T^{N}\left[ \left( \frac{V_{proteins}}{1+V_{proteins}} \right)+\left( \frac{As}{1+As} \right) \right]\left( \frac{c}{1+c} \right)\left( \frac{{IFN}_{\gamma}}{K_{IF}+{IFN}_{\gamma}} \right) \frac{K_{T}}{K_{T}+\left[ PD1-PDL1 \right]}-d_{TN}T^{N}$$

(54)

where the first term describes the source term of Naïve CD8^+^ T cells, the second term describes the activation rate of naïve CD8^+^ T cells, and the last term describes the death rate of naïve CD8^+^ T cells.

2.7.9 Effector CD8^+^ T cells (*T^E^*)

The reaction rate of the effector CD8+ T cells reads

$$\frac{dT^{E}}{dt}=h_{T}{DC}^{*}T^{N}\left[ \left( \frac{V_{proteins}}{1+V_{proteins}} \right)+\left( \frac{As}{1+As} \right) \right]\left( \frac{c}{1+c} \right)\left( \frac{{IFN}_{\gamma}}{K_{IF}+{IFN}_{\gamma}} \right) \frac{K_{T}}{K_{T}+\left[ PD1-PDL1 \right]}+\rho_{T}T^{E}\left( \frac{As}{1+As} \right)\left( \frac{c}{1+c} \right)\left( \frac{{IFN}_{\gamma}}{K_{IF}+{IFN}_{\gamma}} \right) \frac{K_{T}}{K_{T}+\left[ PD1-PDL1 \right]}-\pi_{TM}T^{E}-\varepsilon T^{E}\frac{\left[ PD1-PDL1 \right]}{AS}-\frac{k_{Treg_{2}}T^{E}T_{reg}}{\left( 1+T^{E} \right)\left( 1+T_{reg} \right)}-d_{TE}T^{E}$$

(55)

where the first term describes the activation rate of the naïve CD8^+^ T cells, the second term describes the proliferation rate of the effector CD8^+^ T cells, the third term describes the conversion rate of effector CD8^+^ T cells to memory CD8^+^ T cells and the last terms describe their death rates which depend on the PD1 – PDL1 complex, the regulatory T cells, and the tumor cells.

2.7.10 Macrophages

$$\frac{dMa}{dt}=\chi mc+\chi_{Ma-IL6}[IL6R-IL6]]-\frac{\chi_{DC} c}{1+a}M_{a}\overset{̑}{T}-\gamma aM_{a}$$

(56)

Macrophages are recruited by cytokines and IL-6 which is bound to IL-6R, die with a rate constant γa.

2.7.11 Natural killer cells

The reaction rate of the natural killer cells reads

$$\frac{dNK}{dt}=\frac{\chi_{NK} c}{1+a}-\gamma_{NK}NK$$

(58)

The first term is the production of Natural killer cells depending on the cytokines (pro/anti-inflammatory). The last term is their death rate.

2.7.12 Naïve Β cells (*Β^N^*) (From Ref. (21))

(59)

$$\frac{dB^{N}}{dt}=S_{B}-h_{T}B^{N}{DC}^{*}{Th}^{E}\left[ \left( \frac{V_{proteins}}{1+V_{proteins}} \right)+\left( \frac{As}{1+As} \right) \right]\left( \frac{c}{1+c} \right)\left( \frac{IF}{K_{IF}+IF} \right) \frac{K_{T}}{K_{T}+\left[ PD1-PDL1 \right]} -d_{B}B^{N}$$

where the first term describes the source term of Naïve B cells, the second term describes the activation rate of naıve B cells, and the last term describes the death rate of naıve B cells

2.7.13 Activated Β cells (*Β^Α^*) (From Ref. (21))

$$\frac{dB^{A}}{dt}=h_{T}B^{N}{DC}^{*}{Th}^{E}\left[ \left( \frac{V_{proteins}}{1+V_{proteins}} \right)+\left( \frac{As}{1+As} \right) \right]\left( \frac{c}{1+c} \right)\left( \frac{IF}{K_{IF}+IF} \right) \frac{K_{T}}{K_{T}+\left[ PD1-PDL1 \right]}+\rho_{B}{Th}^{E}B^{A}-\delta_{BA}B^{A}-\pi_{s}B^{A}-\pi_{L}({Th}^{E}+{Th}^{M})B^{A}$$

(60)

where the first term describes the activation rate of naıve B cells, the second term describes the proliferation rate of activated B cells, the following term describes the clearance rate of activated B cells, the next term describes the differentiation rate of activated B cells into short-lived antibody-secreting plasma cells, and the last term describes the differentiation rate of activated B cells into long-lived antibody-secreting plasma cells.

2.7.14 Long-lived plasma (antibody-secreting) B cells (*P^L^*) From Ref. (21))

(61)

$$\frac{dP^{L}}{dt}=\pi_{L}({Th}^{E}+{Th}^{M})B^{A}-\delta_{L}P^{L}$$

where the first term describes the differentiation rate of activated B cells into long-lived antibody-secreting plasma cells by cell-to-cell interactions between effector and memory CD4+ cells (Th^E^ & Th^M^) and activated B cells (B^A^), and the second term describes the death rate of long-lived antibody-secreting plasma cell

2.7.15 Short-lived plasma (antibody-secreting) B cells (*P^S^*) (From Ref. (21))

(62)

$$\frac{dP^{S}}{dt}=\pi_{S}B^{A}-\delta_{S}P^{S}$$

where the first term describes the differentiation rate of activated B cells into short-lived antibody-secreting plasma cells, and the last term describes the death rate of short-lived antibody-secreting plasma cells

2.7.16 Memory CD4+ T cells (Th^M^)

(63)

$$\frac{d{Th}^{M}}{dt}=\pi_{ThM}{Th}^{E}-\delta_{ThM}{Th}^{M}$$

where the first term describes the conversion rate of effector CD4+ cells to memory CD4+-cell, and the last term describes the clearance rate of memory CD4+ cells

2.7.17 Memory CD8+ T cells (T^M^)

(64)

$$\frac{dT^{M}}{dt}=\pi_{TM}T^{E}-\delta_{TM}T^{M}$$

where the first term describes the conversion rate of effector CD8+ cells to memory CD8+-cell, and the last term describes the clearance rate of memory CD8+ cells

3. Cancer Cells

Cancer cell proliferation depends on the oxygen levels in the tissue, and their death rate depends on the interaction of cancer cells with immune cells (effector CD8^+^ T cells, natural killer cells, type 1 macrophages and neutrophils) as well as on the effect of cancer therapy (18, 19). We assume that cancer cells are not infected by the virus.

The mass balance of the tumor cell density is given by a convection-diffusion-reaction equation and it reads

$$\frac{\boldsymbol{\partial}\overset{̑}{\boldsymbol{T}}}{\boldsymbol{\partial t}}\boldsymbol{-}\boldsymbol{\nabla}\boldsymbol{\cdot}\left( \mathbf{D}_{\boldsymbol{Tc}}\boldsymbol{\nabla}\overset{̑}{\boldsymbol{T}} \right)\boldsymbol{=}\frac{\boldsymbol{\lambda}_{\boldsymbol{c}}\boldsymbol{c}_{\boldsymbol{ox}}}{\boldsymbol{k}_{\boldsymbol{c}}\boldsymbol{+}\boldsymbol{c}_{\boldsymbol{ox}}}\overset{̑}{\boldsymbol{T}}\boldsymbol{-}\left( \boldsymbol{n}_{\boldsymbol{T}} \boldsymbol{T}^{\boldsymbol{E}}\overset{̑}{\boldsymbol{T}}\boldsymbol{+}\boldsymbol{n}_{\boldsymbol{T}} \boldsymbol{Nk}\overset{̑}{\boldsymbol{T}}\boldsymbol{+}\boldsymbol{n}_{\boldsymbol{M}\boldsymbol{1}} \boldsymbol{M}_{\boldsymbol{1}}\overset{̑}{\boldsymbol{T}}\boldsymbol{+}\boldsymbol{n}_{\boldsymbol{T}} \boldsymbol{N}\overset{̑}{\boldsymbol{T}} \right) \frac{\overset{̑}{\boldsymbol{T}}}{\left[ \boldsymbol{PD}\boldsymbol{1-PDL}\boldsymbol{1} \right]}\boldsymbol{-}\boldsymbol{d}_{\boldsymbol{Tc}}\overset{̑}{\boldsymbol{T}}$$

(65)

Where the reaction term is given by the proliferation rate which depends on the concentration of the oxygen and by the death rate which depends on the tumor cell density and the immune cells (effector CD8+, natural killer, type 1 macrophages and neutrophils).

4. Vascular density

(66)

$$S_{v}=\frac{[EC]}{{[EC}_{o}]}S_{v}^{0}$$

where $S_{v}^{0}$ is the vascular density of the normal lung and ${[EC}_{o}]$ is the initial endothelial cell population.

5. Oxygen consumption rate(11)

(67)

$V_{O2}=D_{LO2}(P_{A}-P_{b})$

where $D_{LO2}$ is the lung diffusing capacity, $P_{A}$ is the partial pressure of oxygen (*P_O2_*) in alveolar air and $P_{b}$ is the mean *P_O2_* in pulmonary capillary.

The oxygen diffusion can be represented as two components in series, one associated with the alveolar membrane $D_{MO2}$ and one associated with erythrocytes $D_{eO2}$

(68)

$$\left( D_{LO2} \right)^{-1}=\left( D_{MO2} \right)^{-1}+\left( D_{eO2} \right)^{-1}$$

The membrane component is estimated as

(69)

$$D_{MO2}=K_{O2}\frac{\left( \frac{1}{2} \right)[S\left( A \right)+S\left( c \right)]}{\tau_{hb}}$$

where $K_{O2}$ is the Krogh diffusion constant, $S\left( A \right)$ is the average of the alveolar surface area, $S\left( c \right)$ is the capillary surface area and $\tau_{hb}$ is the harmonic mean of the distance between the alveolar surface and the erythrocyte surface.

(70)

$$K_{O2}=\varphi_{KO2}\frac{[EC]}{{[EC}_{o}]}K_{02}^{0}$$

The erythrocyte component is calculated from

(71)

$$D_{eO2}=\theta_{O2}V(c)$$

where $V(c)$ is the pulmonary capillary blood volume and $\theta_{O2}$ is the oxygen unloading conductance of blood.

The partial pressure of oxygen (*P_O2_*) in pulmonary capillary calculated from

$$P_{b}=P_{A}-\frac{V_{o2}^{max}}{D_{LO2}}$$

The oxyhemoglobin saturation according to the Hill equation

(72)

(73)

$$S\left( P_{b} \right)=\frac{\left( \frac{P_{b}}{P_{50}} \right)^{n}}{1+\left( \frac{P_{b}}{P_{50}} \right)^{n}}$$

5.1 Functional vascular density

To quantify the vascular density we assume that it is affected by the decrease in the vessel diameter (d/d_o_) caused by increased number of cancer cells (22) and elevation of solid stress (10) and by the permeability of the tumor vessel wall (23).

The functional vascular density will be given from:

(84)

whereas $S_{V}^{0}$ will depend on vessel wall pore size and $\rho_{v}^{EC}$the density of endothelial cells which is given below. Vessel wall pore size depend on IFNγ concentration as described below (page 7).

5.2 Oxygen Concentration

The rate of change of oxygen in tissues depends on its transport through convection and diffusion, minus the amount of oxygen consumed by cells, plus the amount that enters the tissue from the blood vessels (24, 25), i.e.,

(85)

,

where *c_ox_* is the oxygen concentration, *D_ox_* is the diffusion coefficient of oxygen in the interstitial space, *A_ox_* and *k_ox_* are oxygen uptake parameters, *P_er_* is the vascular permeability of oxygen that describes diffusion across the tumor vessel wall and *C_iox_* is the oxygen concentration in the vessels.

6. Vaccination-induced Immunity (26-29)

The model considers separately the mechanisms of mRNA and vector vaccines. The vaccines , as particles, either lipid nanoparticles in the case of mRNA vaccines or viral-vector in the case of vector vaccines, enter host cells and either induce DNA transcription to mRNA (vector vaccine) and then translation into viral antigen or result directly in translation of viral antigens (mRNA vaccine). Subsequently, vaccine-induced peptides exit the cells and interact with dendritic cells to produce antigen presenting cells. These subsequently activate T cells and B cells to create CD4+ and CD8+ effector and memory T cells as well as short-lived and long-lived plasma (antibody-secreting) B cells.

6.1.1 Free vaccine [adenovirus (DNA) or lipid coat (mRNA)]

$$\frac{dVa}{dt}=S^{Va}+\frac{Q_{lung}Va}{V_{lung}}-\frac{\left( Q_{lung}-L_{lung} \right)Va}{V_{lung}}-K_{cell-Vaccine}^{on}VaH+K_{cell-Vaccine}^{off}VaH-K_{dVa}Va$$

(74)

where the terms respectively describe convectional transport of free vaccine particles (adenovirus or lipid coat) in lung, the binding of the vaccine particles to cells (Healthy and Antigen presenting cells-APCs) membranes, the detachment of bound vaccine particles from cells membranes and the inactivation of the vaccine particles.

6.1.2 Bound vaccine [adenovirus (DNA) or lipid coat (mRNA)]

(75)

$$\frac{d{Va}_{b}}{dt}=K_{cell-Vaccine}^{on}VaH-K_{cell-Vaccine}^{off}VaH-Kint{Va}_{b}-K_{dVab}{Va}_{b}$$

where the terms respectively describe the binding/unbinding of the vaccine particles on the cells membrane, the internalization of the bound vaccine particles into the cells and the last term is the degradation of the bound vaccine particles.

6.1.3 Internalized vaccine [adenovirus (DNA)]

$$\frac{d{Va}_{int}}{dt}=K_{intVa}{Va}_{b}-d_{Vaint}$$

(76)

where the first term describes the internalization of the bound vaccine particles and the second term describes the constant deradation of the internalized vaccine particles.

6.1.4 DNA Transcription [adenovirus (DNA)]

(77)

$$\frac{d{Va}_{Tran}}{dt}=K_{Tran}{Va}_{int}-d_{VaTran}$$

where the first term describes the DNA transcription of the internalized vaccine particles and the second term describes the constant degradation of the translation of the DNA.

6.1.5 Production Viral proteins [adenovirus (DNA)]

(78)

$$\frac{dV_{proteins}}{dt}=K_{protein}{Va}_{Tran}-d_{Vprotein}$$

where the first term describes the production of viral protein or viral antigen from the translation of the mRNA and the last describes the constant degradation of viral proteins.

6.1.6 Internalized vaccine [lipid coat (mRNA)]

$$\frac{d{Va}_{int}}{dt}=K_{intVa}{Va}_{b}-d_{Vaint}$$

(79)

where the first term describes the internalization of the bound vaccine particles, the second term describes the constant degradation of the internalized vaccine particles.

6.1.7 Production Viral proteins [lipid coat (mRNA)]

(80)

$$\frac{dV_{proteins}}{dt}=K_{protein}{Va}_{int}-d_{Vprotein}$$

where the first term describes the production of viral protein or viral antigen from the translation of the mRNA and the last describes the constant degradation of viral proteins.

Table S1. Model Parameters related to vaccines

| Parameter | Description |
| --- | --- |
| $\boldsymbol{S}^{\boldsymbol{Va}}$ | Source term for Free vaccine |
| $\boldsymbol{K}_{\boldsymbol{cell}\mathbf{-}\boldsymbol{Vaccine}}^{\boldsymbol{on}}$ | Binding of the vaccine particles to Healthy cells |
| $\boldsymbol{K}_{\boldsymbol{APCs}\mathbf{-}\boldsymbol{Vaccine}}^{\boldsymbol{on}}$ | Binding of the vaccine particles to APCs cells |
| $\boldsymbol{K}_{\boldsymbol{cell}\mathbf{-}\boldsymbol{Vaccine}}^{\boldsymbol{off}}$ | unbinding of the vaccine particles to Healthy cells |
| $\boldsymbol{K}_{\boldsymbol{APCs}\mathbf{-}\boldsymbol{Vaccine}}^{\boldsymbol{off}}$ | unbinding of the vaccine particles to APCs cells |
| $\boldsymbol{K}_{\boldsymbol{d}}$ | Degradation rate of vaccine particles |
| $\boldsymbol{K}_{\boldsymbol{int}}$ | internalization of the bound vaccine particles into the cells |
| $\boldsymbol{K}_{\boldsymbol{dVab}}$ | degradation of the bound vaccine particles |
| $\boldsymbol{K}_{\boldsymbol{Tran}}$ | DNA transcription of the internalize vaccine particles |
| $\boldsymbol{K}_{\boldsymbol{protein}}$ | production of viral protein or viral antigen from the translation of the mRNA |
| $\boldsymbol{d}_{\boldsymbol{Vaint}}$ | constant of degradation of internalized vaccine |
| $\boldsymbol{d}_{\boldsymbol{VaTran}}$ | constant of degradation of translation DNA |
| $\boldsymbol{d}_{\boldsymbol{Vprotein}}$ | constant degradation of viral proteins |
